# Estimating the historical impact of outbreak response immunization programs across 210 outbreaks in LMICs

**DOI:** 10.1101/2024.06.02.24308241

**Authors:** D. Delport, A.M. Muellenmeister, G. MacKechnie, S. Vaccher, T. Mengistu, D. Hogan, R.G. Abeysuriya, N. Scott

**Affiliations:** Burnet Institute, Melbourne, VIC 3004, Australia; Department of Epidemiology and Preventive Medicine, Monash University, Melbourne, VIC 3004, Australia; School of Population and Global Health, The University of Melbourne, Parkville, VIC 3010, Australia; Gavi, The Vaccine Alliance, 1218 Geneva, Switzerland

## Abstract

**Background:** Outbreaks of vaccine-preventable diseases continue to occur in low- and middle-income countries (LMICs), requiring outbreak response immunization (ORI) programs for containment. To inform future investment decisions, this study aimed to estimate the cases, deaths, disability-adjusted life years (DALYs), and societal economic costs averted by past ORI programs. Outbreaks of measles, Ebola, yellow fever, cholera, and meningococcal meningitis in LMICs between 2000-2023 were considered.

**Methods:** 210 outbreaks (51 measles, 40 cholera, 88 yellow fever, 24 meningitis, 7 Ebola) were identified with sufficient data for analysis. Agent-based models were calibrated for each disease such that after controlling for baseline vaccine coverage, ORI initiation time, speed of vaccine delivery, environmental variables, or endemic prevalence of the disease, observed outbreaks were within the distribution of simulated outbreaks. A status-quo and no ORI scenario were compared for each outbreak.

**Findings:** Across 210 outbreaks, ORI programs are estimated to have averted 5·81M [95% uncertainty interval 5·75M–5·87M] cases (4·01M measles, 283K cholera, 1·50M yellow fever, 21·3K meningitis, 820 Ebola), 327K [317K–338K] deaths (20.0K measles, 5215 cholera, 300K yellow fever, 1599 meningitis, 381 Ebola), 14·6M [14·1M–15·1M] DALYs (1·27M measles, 220K cholera, 13·0M yellow fever, 113K meningitis, 16·6K Ebola), and US$31·7B [29·0B–34·9B] (US$710M measles, US$156M cholera, US$30·7B yellow fever, US$97·6M meningitis, US$6·72M Ebola) in societal economic costs. In general, the more rapidly the ORI was initiated the greater the impact.

**Interpretation:** ORI programs are critical for reducing the health and economic impacts of outbreaks of vaccine-preventable diseases.

**Funding:** Gavi,the Vaccine Alliance.

## Introduction

Estimates from the Global Burden of Disease indicate that between 2015-2019 around 2 million deaths per year from were attributable to vaccine preventable diseases (VPDs), with 60% of the burden in low- and middle-income countries (LMICs)^1^. Even diseases with routine childhood immunization programs (e.g., measles, and yellow fever in endemic countries) still have frequent outbreaks and a high health burden due to gaps in vaccine coverage. When an outbreak of a VPD occurs the health impact can be minimized with containment measures, a major component being outbreak response immunization (ORI) campaigns to limit outbreak size by rapidly building immunity^2^.

When a country does not have sufficient vaccine supply for ORI to cover the population at-risk, they can request disbursements from global stockpiles. Since 2000, Gavi, the Vaccine Alliance (Gavi) has funded the introduction and scale-up of new vaccines through routine systems, preventive vaccination campaigns, and ORI campaigns^3^. Gavi funds outbreak response for measles and measles-rubella through the Measles and Rubella Partnership, and emergency stockpiles, managed through the International Coordinating Group for vaccine provision (ICG), for cholera, yellow fever, meningococcal meningitis strains A, C, W, and Y, and Ebola^2,3^.

Global vaccine stockpiles are insurance for reducing the scale of outbreaks when they occur^4^, however they remain unused if no outbreak occurs before the vaccines expire. Evidence is needed to quantify the value of vaccine stockpiles for ORI to inform future investment. A methodological framework exists for estimating the impact of ORI programs in LMICs^5^ using mathematical models, outlining specific health and economic outcomes identified by stakeholders as both feasible to estimate and useful, such as cases, deaths, and, societal economic costs averted^5^.

To inform decisions around investment in vaccine stockpiles, this analysis aimed to estimate the health and economic impact of ORI programs for measles, cholera, yellow fever, meningococcal meningitis, and Ebola in LMICs since 2000. Five disease models were developed, as well as a novel methodology for calibrating them across many outbreaks while accounting for disease, setting, and ORI specific factors that influence outbreak size.

## Methods

We provide a brief overview of key methods here, with Supplement 1 describing the overall analytical approach across all diseases, and Supplements 2-6 describing the five disease-specific models and analyses.

### Outbreak data

Data from outbreaks of measles, cholera, yellow fever, meningococcal meningitis, and Ebola in LMICs between 2000 and 2023 were collated from a variety of sources, including WHO *Weekly Epidemiological Record*^6^ and *Disease Outbreak News*^7^, the MenAfriNet *Meningitis Weekly Bulletin*^8^, the ICG vaccine dashboards^9^, and academic literature which reported relevant outbreaks^10,11^. To be included in the analysis, an outbreak required data on a documented ORI programmatic response, the time between outbreak declaration and vaccine response, the number of vaccines delivered during the response, the response duration, and other details which varied by disease. Full details are available in the ‘Outbreak data’ sections of Supplements 2-6.

### Simulation models

Agent-based models were constructed for each disease. Population characteristics were representative of the target ORI populations reported across the included outbreaks in LMICs, and model structure, parametrization, and disease dynamics were informed by previous modelling work and available literature. Figure 1 shows the schematic representation of each disease model, and details of each model can be found in the ‘Model overview’ and ‘Model population for simulated outbreaks’ sections of Supplements 2-6.

**Figure 1:**
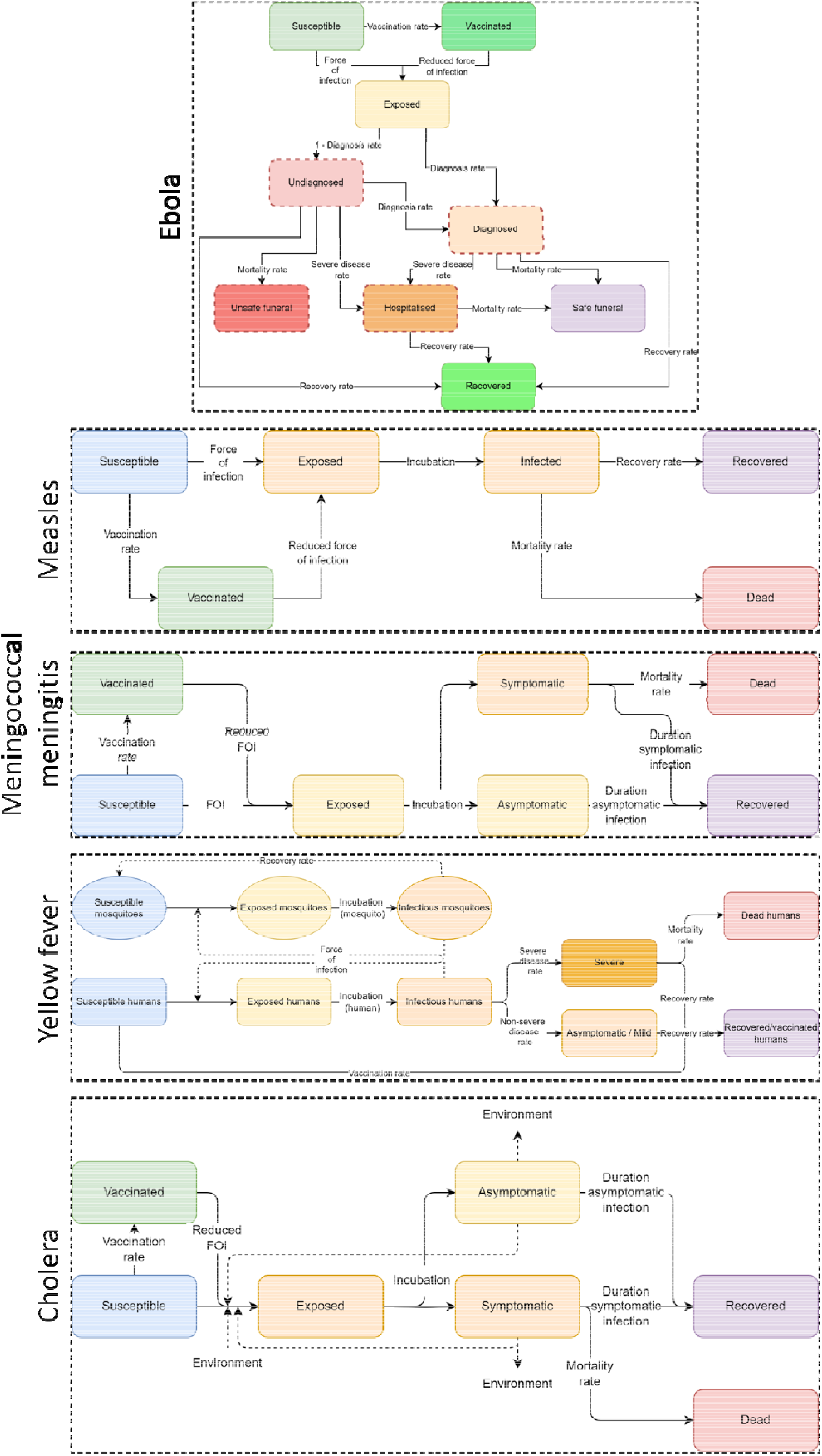
Model schematics. Schematic representations of the states and parameters influencing disease transmission in the agent-based models for each of Ebola, measles, meningococcal meningitis, yellow fever, and cholera.

Modelling was conducted in Python using the *Starsim* framework. *Starsim* is an open-source agent-based modelling framework developed by the Institute for Disease Modeling at the Bill and Melinda Gates Foundation in collaboration with the Burnet Institute and other partners^12^, which allows for rapid development of disease models that can be arbitrarily customized, including transmission mechanisms and interventions.

### Calibration and the lattice-based approach

The number of historical outbreaks under consideration and limited or inconsistent data available from each setting made it computationally impractical to calibrate outbreaks individually. To overcome this, we developed what we refer to as a ‘lattice’-based approach, with outbreak simulations pre-computed on a grid of key parameter values, with individual outbreaks assigned to the best-fitting point in the lattice based on the nearest parameter values. The key parameters were those expected to be influential on model outcomes; for all diseases, this included response delay and response speed (vaccine doses delivered per capita per day). For some diseases additional covariates were included:

- For measles, baseline vaccine coverage before the outbreak to account for differences in susceptibility across settings. Additional details are available in Supplement 2.
- For yellow fever, baseline vaccine coverage before the outbreak, and a ‘transmission level’ parameter capturing differences in environmental factors affecting the mosquito population, changing the level of mosquito-related transmission by a multiplicative factor between 0·5 – 1·5. Additional details are available in Supplement 4.
- For meningitis, the vaccine type used (polysaccharide or conjugate), the target age-group for the response, and asymptomatic carrier prevalence before the outbreak to account for the range of outbreak sizes observed in the data. Additional details are available in Supplement 5.

The range and spacing of the lattice points in each dimension was selected to minimize the discrepancy between individual outbreaks and their assigned lattice points, while also minimizing the number of lattice points to improve computational tractability. Figure 2 shows an example set of lattice points for response time, vaccination rate, and baseline vaccine coverage in the setting. For three disease-specific parameters (vaccine coverage for measles, mosquito transmission level for yellow fever, and asymptomatic carrier prevalence for meningitis), data were not available to inform values in the model. Therefore, these lattice values for each outbreak in the dataset were estimated during calibration, informed by the maximum-likelihood estimate. For each disease at most one lattice dimension was estimated through calibration. Additional details for these methods are in Supplement 1, ‘Kernel Density Estimation’.

**Figure 2:**
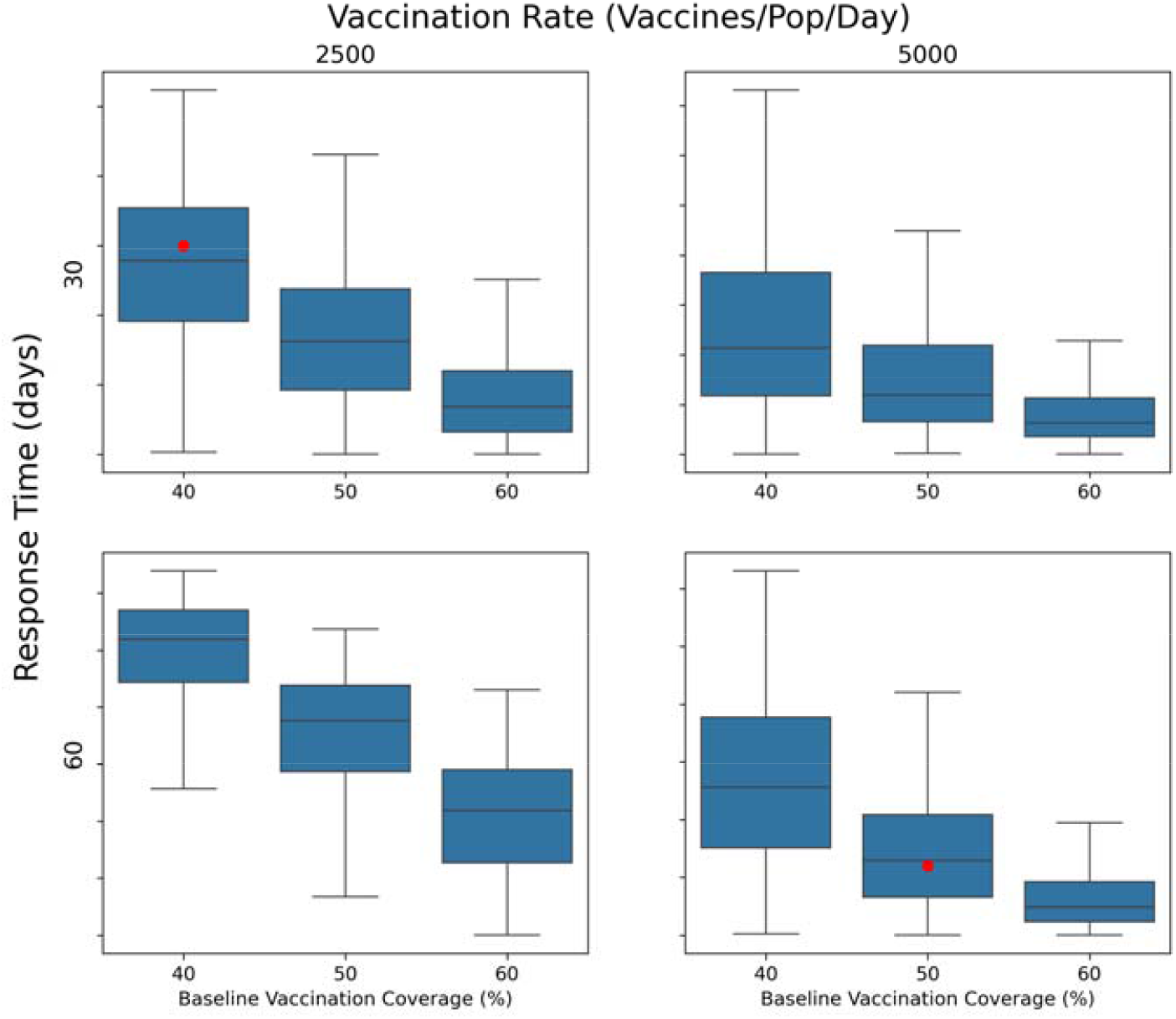
Lattice Approach: Example lattice with two values each for a response time (days) and vaccination rate (vaccines/population/day). In this example, baseline vaccination coverage (%) among the ORI target population is unknown and estimated by best alignment with of the total number of cumulative cases in the outbreak (e.g., positioning of red dots). As such, while the vaccination rate and response time dimensions are informed by observed data for a given outbreak, the third dimension (baseline vaccination coverage) is informed by the maximum-likelihood estimate, and all three dimensions determine the lattice point to which an outbreak is assigned.

Additionally, stochastic effects meant that calibration to an individual outbreak may not result in generalizable fitted parameters. Therefore, calibration aimed to result in all observed outbreaks being within the distribution of stochastically simulated outbreaks with the corresponding lattice parameters.

Outbreak size and population at-risk of infection varies greatly, however model simulations were conducted using a fixed number of agents, and individual outbreaks were scaled down to the number of agents in the model, meaning that in some cases one agent represented more than one person. Additional details are available in Supplement 1, ‘Model population and target population scaling’.

Table 1 details some key epidemiological and response characteristics utilised across the five diseases, and how they vary by disease. Further details are available in Supplements 2-6.

**Table 1:** Summary of key model characteristics for each disease considered.

|  | Modelled population age | Transmission type | Background / routine vaccination | Asymptomatic cases | Non-pharmaceutical interventions considered | Additional lattice dimensions |
| --- | --- | --- | --- | --- | --- | --- |
| **Measles** | 0-5 | Direct | Y | N | N | Baseline vaccine coverage |
| **Cholera** | All ages | Direct + Environmental (water) | N | Y | Water, Sanitation, and Hygiene | - |
| **Yellow fever** | All ages | Vector-borne | Y | Y | N | Baseline vaccine coverage, environmental transmission level |
| **Meningitis** | All ages | Direct (inc. asymptomatic) | N (Meningitis A outbreaks not included) | Y | N | Asymptomatic carrier prevalence, vaccine type, target age group |
| **Ebola** | All ages | Direct (inc. unsafe burial) | N | N | Contact tracing, quarantine, isolation | - |

### Scenarios

For a given set of key parameters (i.e., lattice point), model simulations produced a wide range of outcomes, reflecting a range of possibilities for *any* potential outbreak under those conditions. However, when constructing a set of baseline simulations for a *known* individual outbreak, uncertainty can be reduced by accepting or rejecting individual simulations according to whether they are realistic representations, defined as being within +/-25% of the observed cases (we refer to this as ‘filtering’, see Supplement 1, ‘Trajectory selection’). For outbreaks without estimates of cumulative cases, the entire range of the model uncertainty was included.

Using the set of filtered simulations for each historic outbreak, two scenarios were examined to assess the impact of ORI:

- *Baseline*, the status-quo with ORI as occurred;
- *No ORI* counterfactual, where simulations were run without any ORI intervention. This scenario used the same filtered simulations as the *Baseline* scenario, such that everything was identical up until the date the ORI would have started.

### Analysis

For each outbreak’s *Baseline* and *No ORI* scenarios, the distribution of cumulative cases and deaths across model simulations were recorded. The mean difference in outbreak size between the two scenarios and associated 95% uncertainty intervals (95% UI) for the mean were estimated from these collections of simulations using bootstrap resampling (i.e., for each outbreak to produce estimates of cases averted by ORI).

The total cases averted by ORI across all outbreaks were then estimated by aggregating the cases averted for each individual outbreak. As the outbreaks are independent, this was obtained by summing the cases averted per outbreak, with the variance estimated by summing the variances from each individual outbreak.

Disability-adjusted life years (DALYs) and socio-economic costs averted by ORI were also estimated for each outbreak: DALYs by multiplying cases averted by the disability weight per case^13^ and average duration of symptoms plus years of life lost from deaths, and costs by estimating productive years of life lost or lived with disability, and multiplying this by gross domestic product per capita^14^ (inflated to 2023 USD and discounted at 3% per annum). Additional methodological details can be found in Supplement 1.

The impact of ORI on reducing the risk of outbreaks reaching different thresholds of cumulative cases was estimated by comparing the distribution of cumulative cases across outbreaks in the data to the distribution of cumulative cases across outbreaks in the *No ORI* scenario. For the purposes of this sub-analysis, the median cumulative cases in the *No ORI* scenario was used as a point estimate for each outbreak. Additional details are available in Supplement 1.

## Results

### Outbreaks data

In total, 51 measles, 40 cholera, 88 yellow fever, 24 meningitis, and 7 Ebola outbreaks were included, occurring in 49 LMICs from five world regions. LMIC status was determined based on World Bank classification according to gross national income per capita^15^. Supplements for each disease contain additional summary details of the outbreak data included (Tables S2, S5, S8, S11, and S14).

### ORI impact on health and economic outcomes

We estimate that across 210 outbreaks of these diseases in LMICs from 2000–2023, ORI programs have averted 5·81M (95%UI 5·75M – 5·87M) cases, 327K (317K – 338K) deaths, 14·6M (14·1M – 15·1M) DALYs, and US$31·7B (29·0B – 34·4B) in societal economic costs (Table 2). Differences across diseases were driven by the number of outbreaks included in the analysis, disease severity, and characteristics of the response (both ORI and non-ORI). Outcomes associated with just the subset of ORI in settings eligible for Gavi support are in Supplement 7.

**Table 2:**
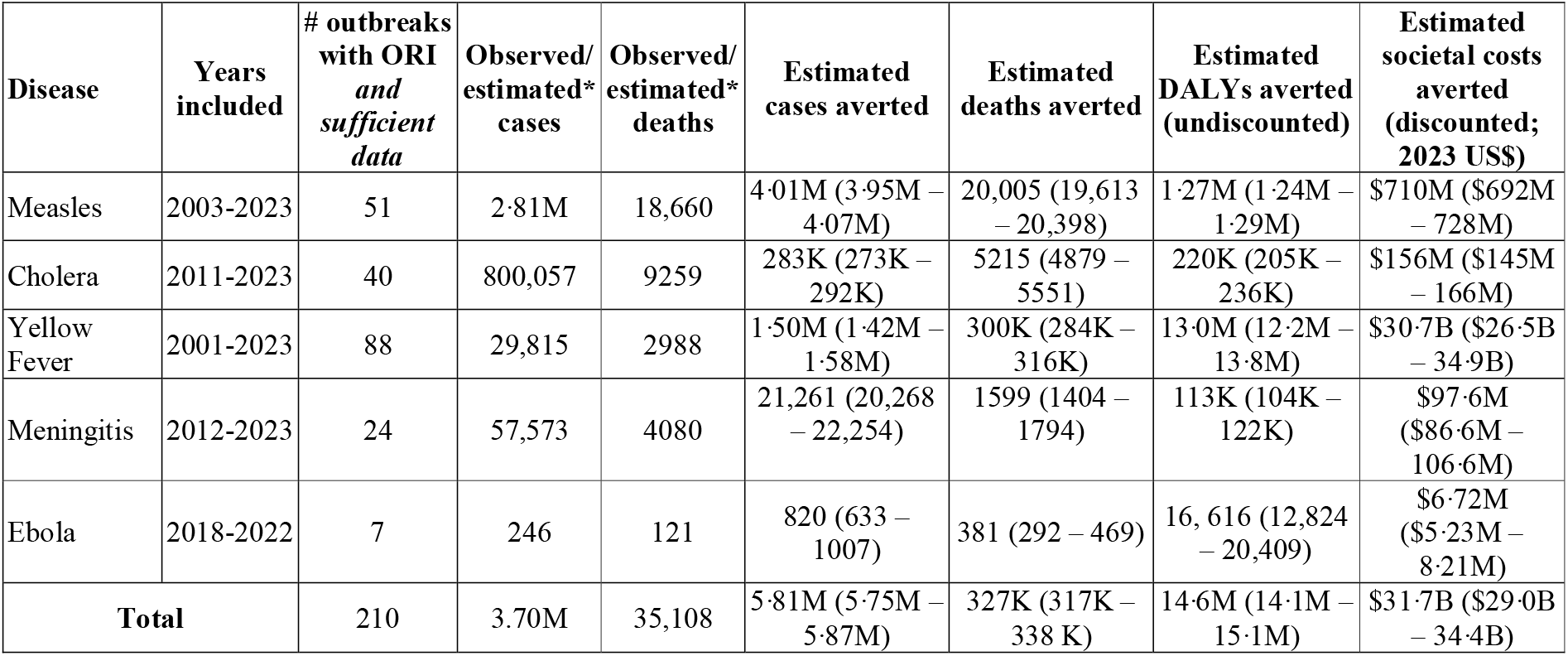
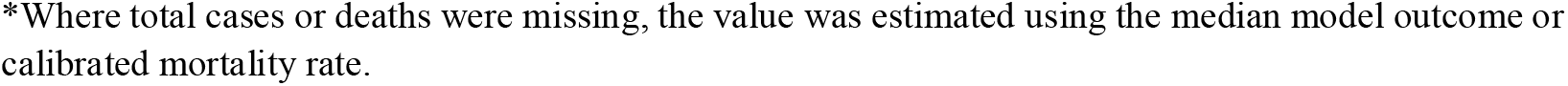
Summary impacts of ORI for the five diseases considered. Estimated cases/deaths/disability-adjusted life years (DALYs)/societal economic costs averted are summarized as the mean and 95% uncertainty intervals.

| Disease | Years included | # outbreaks with ORI and sufficient data | Observed/estimated* cases | Observed/estimated* deaths | Estimated cases averted | Estimated deaths averted | Estimated DALYs averted (undiscounted) | Estimated societal costs averted (discounted; 2023 US\$) |
| --- | --- | --- | --- | --- | --- | --- | --- | --- |
| Measles | 2003-2023 | 51 | 2.81M | 18,660 | 4.01M (3.95M – 4.07M) | 20,005 (19,613 – 20,398) | 1.27M (1.24M – 1.29M) | \$710M (\$692M – 728M) |
| Cholera | 2011-2023 | 40 | 800,057 | 9259 | 283K (273K – 292K) | 5215 (4879 – 5551) | 220K (205K – 236K) | \$156M (\$145M – 166M) |
| Yellow Fever | 2001-2023 | 88 | 29,815 | 2988 | 1.50M (1.42M – 1.58M) | 300K (284K – 316K) | 13.0M (12.2M – 13.8M) | \$30.7B (\$26.5B – 34.9B) |
| Meningitis | 2012-2023 | 24 | 57,573 | 4080 | 21,261 (20,268 – 22,254) | 1599 (1404 – 1794) | 113K (104K – 122K) | \$97.6M (\$86.6M – 106.6M) |
| Ebola | 2018-2022 | 7 | 246 | 121 | 820 (633 – 1007) | 381 (292 – 469) | 16,616 (12,824 – 20,409) | \$6.72M (\$5.23M – 8.21M) |
| **Total** | | 210 | 3.70M | 35,108 | 5.81M (5.75M – 5.87M) | 327K (317K – 338K) | 14.6M (14.1M – 15.1M) | \$31.7B (\$29.0B – 34.4B) |

\*Where total cases or deaths were missing, the value was estimated using the median model outcome or calibrated mortality rate.

### Measles

Across the 51 measles outbreaks from 2001-2023 with sufficient data for analysis, ORI programs were estimated to have averted 4·01M (3·95M – 4·07M) cases and 20,005 (19,613 – 20,398) deaths. In most outbreaks this represents upwards of 40% of cases and deaths averted relative to the *No ORI* counterfactual, except for outbreaks in settings with close to 100% population baseline vaccine coverage in children over 9 months of age (Figure 3). ORI tended to have greater impact in settings with shorter response times (Figure S10 in Supplement 2).

**Figure 3:**
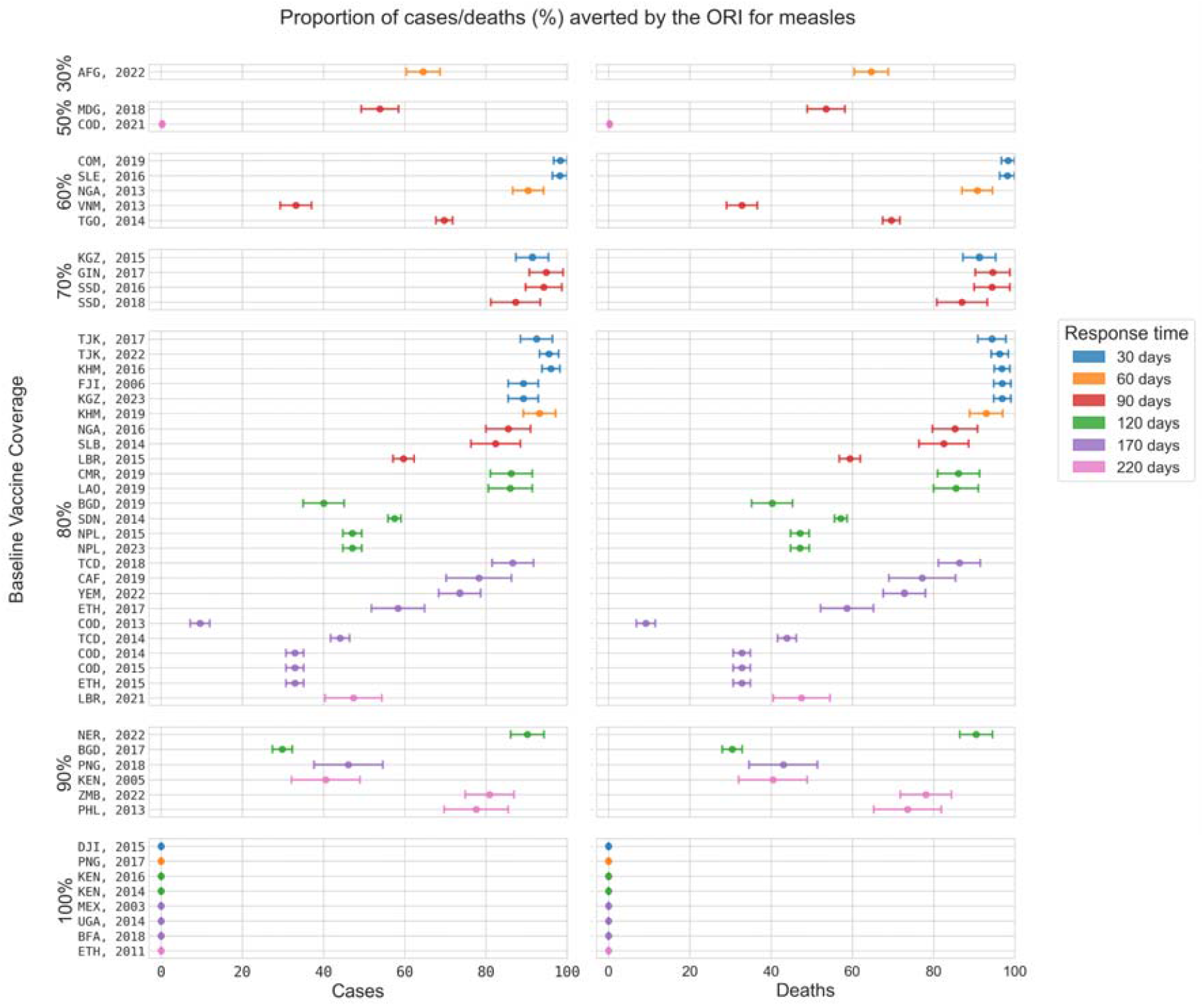
Proportion of cases and deaths averted by the ORI for each modelled measles outbreak. Outbreaks are grouped by baseline vaccine coverage and coloured by response time (lowest to highest). The largest historic outbreaks typically occurred in settings estimated to have lower vaccine coverages (further details in Supplement 2), and the estimated proportional impact from the ORI was lower in these settings. For example, the largest outbreak, in the Democratic Republic of the Congo in 2021, was estimated to have occurred in a population with 50% baseline vaccine coverage and the ORI was estimated to have averted a small proportion of cases because the outbreaks produced in the counterfactual scenario were of a similar size to what was observed. Three letter country codes indicate the location of the outbreak, the year indicates the year when the outbreak began.

### Cholera

Across the 40 modelled cholera outbreaks from 2015-2023 with sufficient data for analysis, we estimate that the ORI programs averted 283K (273K – 292K) cases and 5215 (4879 – 5551) deaths. This represents 10% – 60% of cases and deaths being averted in most outbreaks relative to the *No ORI* counterfactual. A higher proportion of cases and deaths were averted in settings with quicker response times (Figure 4). The smallest proportional impacts were observed when the ORI took 130 days to begin as most cholera outbreaks are observed to be relatively short (see Figure S12 in Supplement 3 for the time series of outbreaks against which the model was calibrated), and therefore most transmission occurs within the model before the vaccines are delivered.

**Figure 4:**
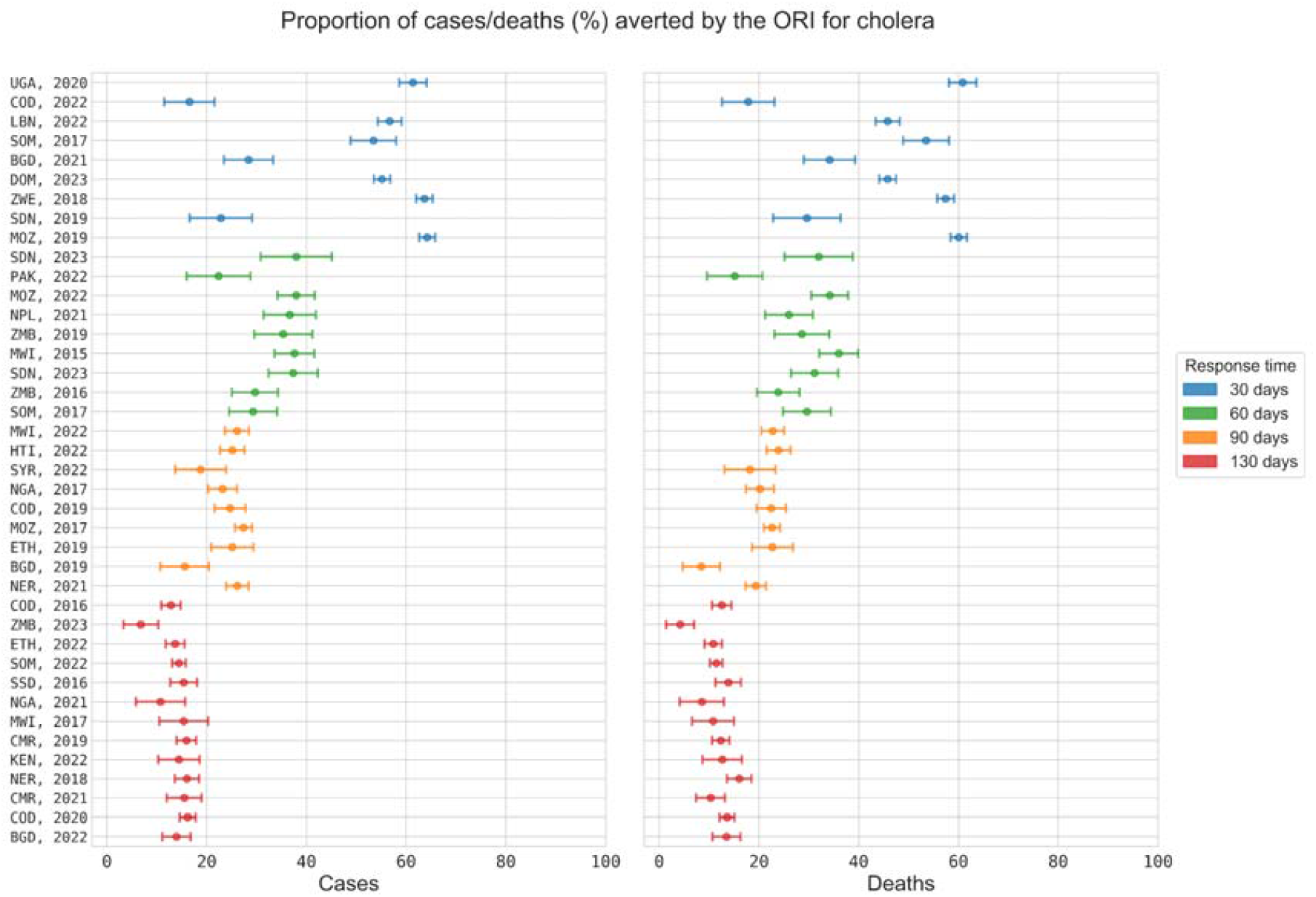
Proportion of cases and deaths averted by the ORI for each modelled cholera outbreak. From top to bottom, outbreaks are ordered and coloured by response time (fastest to slowest). Three letter country codes indicate the location of the outbreak, the year indicates the year when the outbreak began.

### Yellow fever

Across the 88 modelled yellow fever outbreaks with sufficient data for analysis, from 2000-2023 we estimate that ORI programs averted 1·50M (1·42M – 1·58M) cases and 300K (284K – 316K) deaths. The ORI tended to have greater impact in settings with low baseline immunities, high transmission, and short response times (Figure 5). The estimated proportional outcomes averted for most yellow fever outbreaks is small, less than 10% – 20%, as most outbreaks were estimated to have occurred in low transmission settings, producing small case burdens. However, for settings where transmissibility was high due to favourable environmental conditions for the mosquito population, the ORI was highly impactful, averting almost all cases and deaths, and this impact was increased if the population immunity was low before an outbreak occurred.

**Figure 5:**
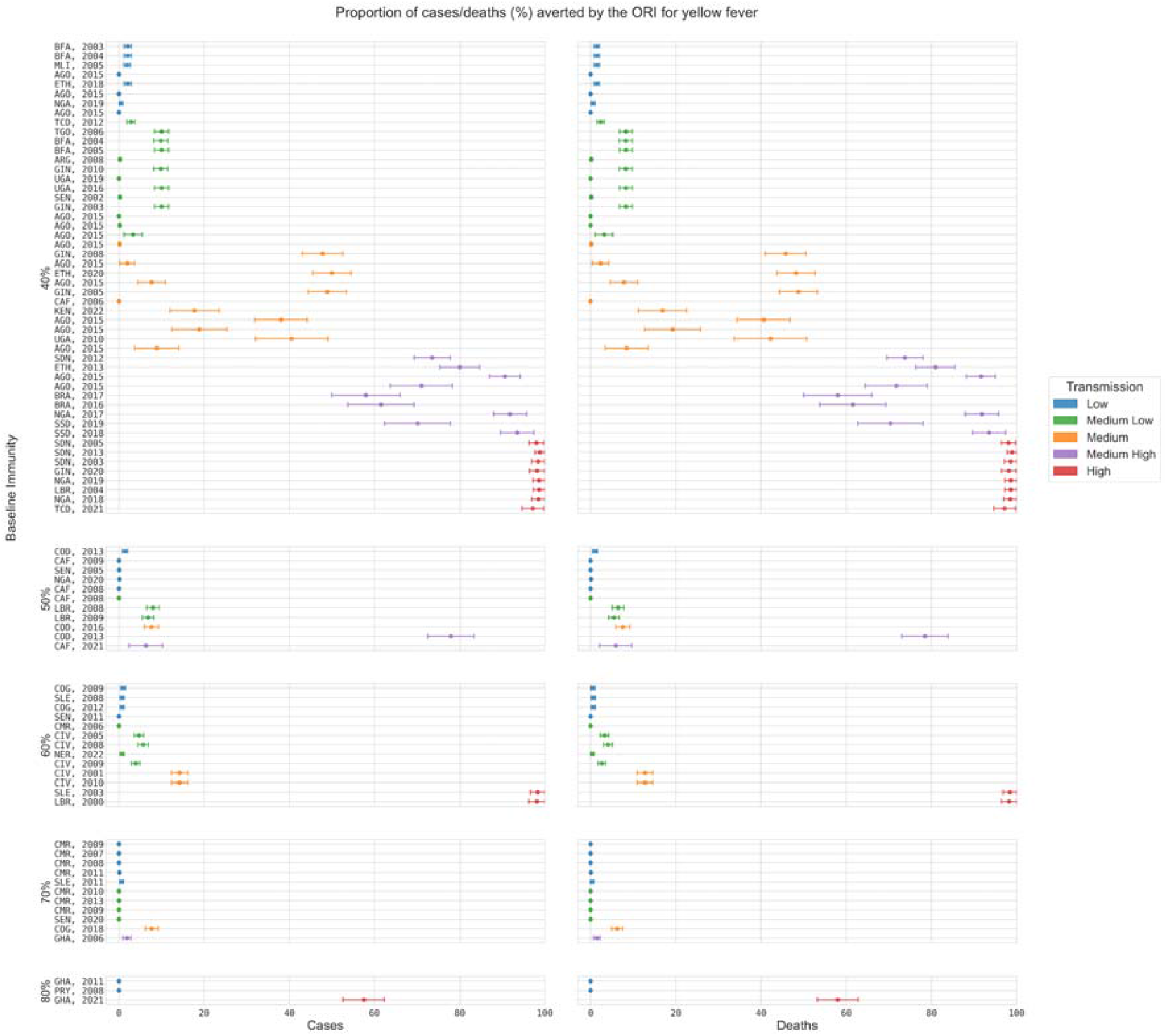
Proportion of cases and deaths averted by the ORI for each modelled yellow fever outbreak. From top to bottom, outbreaks are grouped by baseline coverage (lowest to highest), and within each group the outbreaks are ordered and coloured by the estimated transmission level (low to high). The estimated transmission level acted as a multiplicative modifier for mosquito-to-human transmission probability, capturing variation in mosquito populations and biting rates; further details on the transmission level estimation are in Supplement 4. Three letter country codes indicate the location of the outbreak, the year indicates the year when the outbreak began.

### Meningitis

The ORI is estimated to have averted between 10% – 40% of cases and deaths across most of the 24 modelled meningitis outbreaks with sufficient data for analysis. From 2012-2023, we estimate that the ORI programs averted 21,261 (20,268 – 22,254) cases and 1599 (1404 – 1794) deaths. A higher proportion of cases and deaths were averted relative to the *No ORI* counterfactual in settings when the response time was quicker (Figure 6).

**Figure 6:**
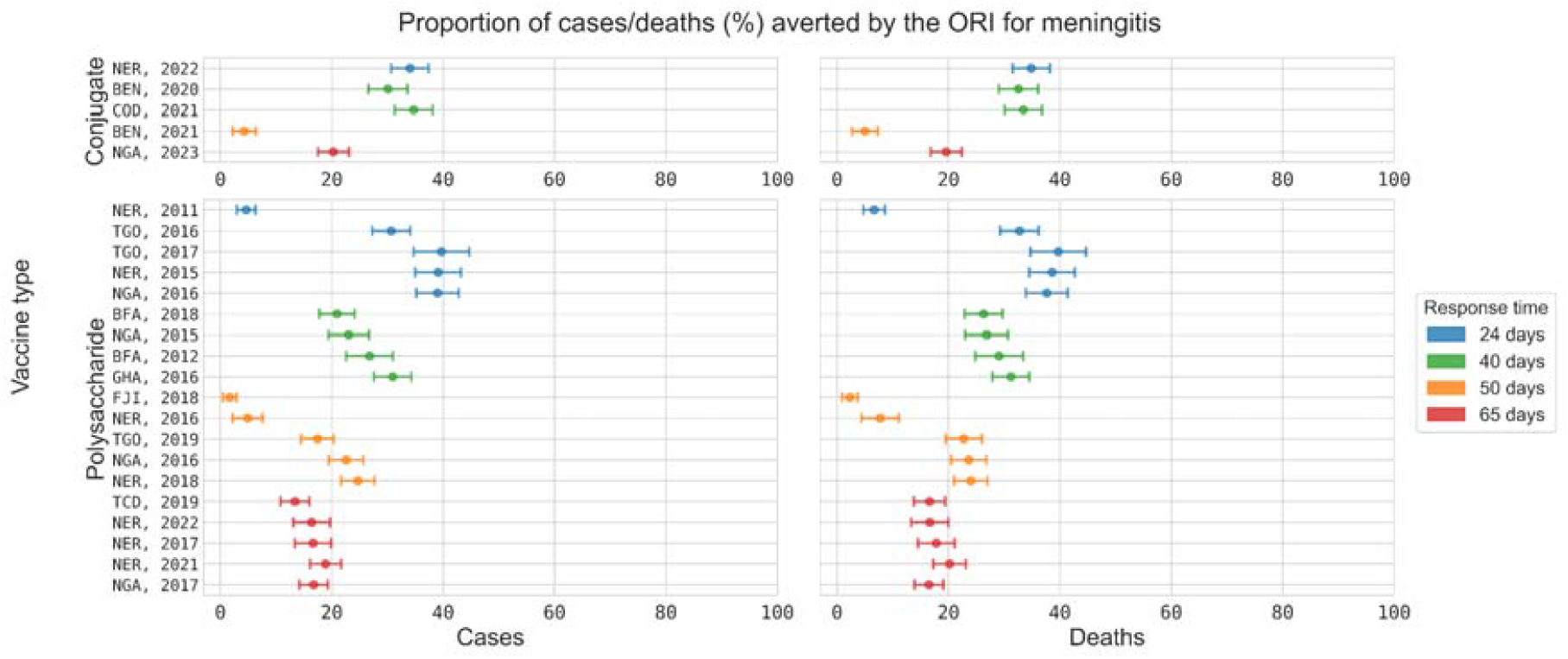
Proportion of cases and deaths averted by the ORI for each modelled meningitis outbreak. Outbreaks are grouped by the type of vaccine used in the response (conjugate or polysaccharide); further details on the differences between the modelled vaccines are in Supplement 5. Within each group, outbreaks are ordered and coloured by response time (fastest in blue to slowest in red). The smallest proportional impacts were observed for outbreaks with low case burdens, such as Niger in 2011 and 2016, Fiji in 2018, and Benin in 2018. This likely occurred because the outbreaks ‘fizzled out’ before the ORI began and therefore the counterfactual simulations also only produce small outbreaks and little difference is observed when comparing against the baseline scenario. Three letter country codes indicate the location of the outbreak, the year indicates the year when the outbreak began.

### Ebola

Across the seven modelled outbreaks with sufficient data for analysis, from 2018-2022 we estimate that the ORI programs averted 820 (633 – 1007) cases and 381 (292 – 469) deaths. ORI is estimated to have averted a relatively consistent proportion of cases and deaths across all seven outbreaks, typically 35 – 55% relative to the *No ORI* counterfactual (Figure 7).

**Figure 7:**
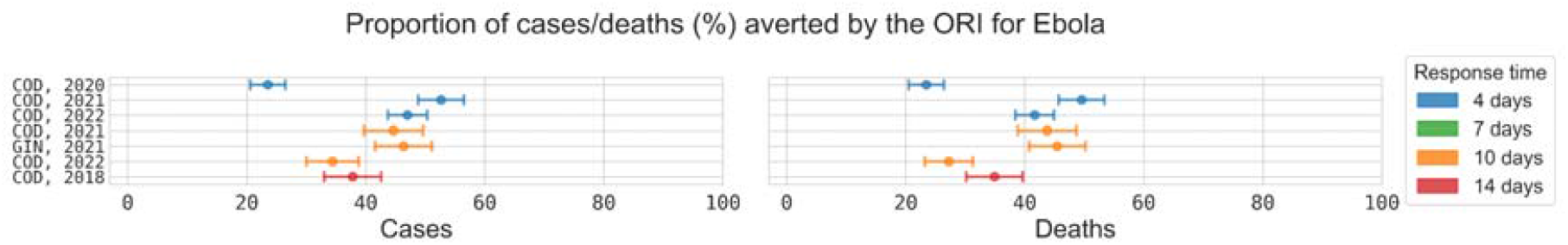
Proportion of cases and deaths averted by the ORI for each modelled Ebola outbreak. From top to bottom, outbreaks are ordered and coloured by response time (fastest in blue to slowest in red). Notably the 2020 outbreak in the Democratic Republic of Congo averts a smaller proportion of outcomes than the other outbreaks considered. The model estimates that approximately 20% of cases and deaths were averted despite a rapid response time. This is in line with the proportional impact estimated for this outbreak in a previous analysis^5^, where contact tracing was believed to have been particularly effective even in the absence of a vaccine response. Three letter country codes indicate the location of the outbreak, the year indicates the year when the outbreak began.

ORI impact on outbreak size

For all diseases considered, the presence of ORI is estimated to have reduced the expected number of outbreaks reaching different threshold sizes. The definition of a ‘large outbreak’ will be setting and disease specific and may be interpreted differently by different stakeholders, but some examples of key ORI impacts include (Figure 8):

**Figure 8:**
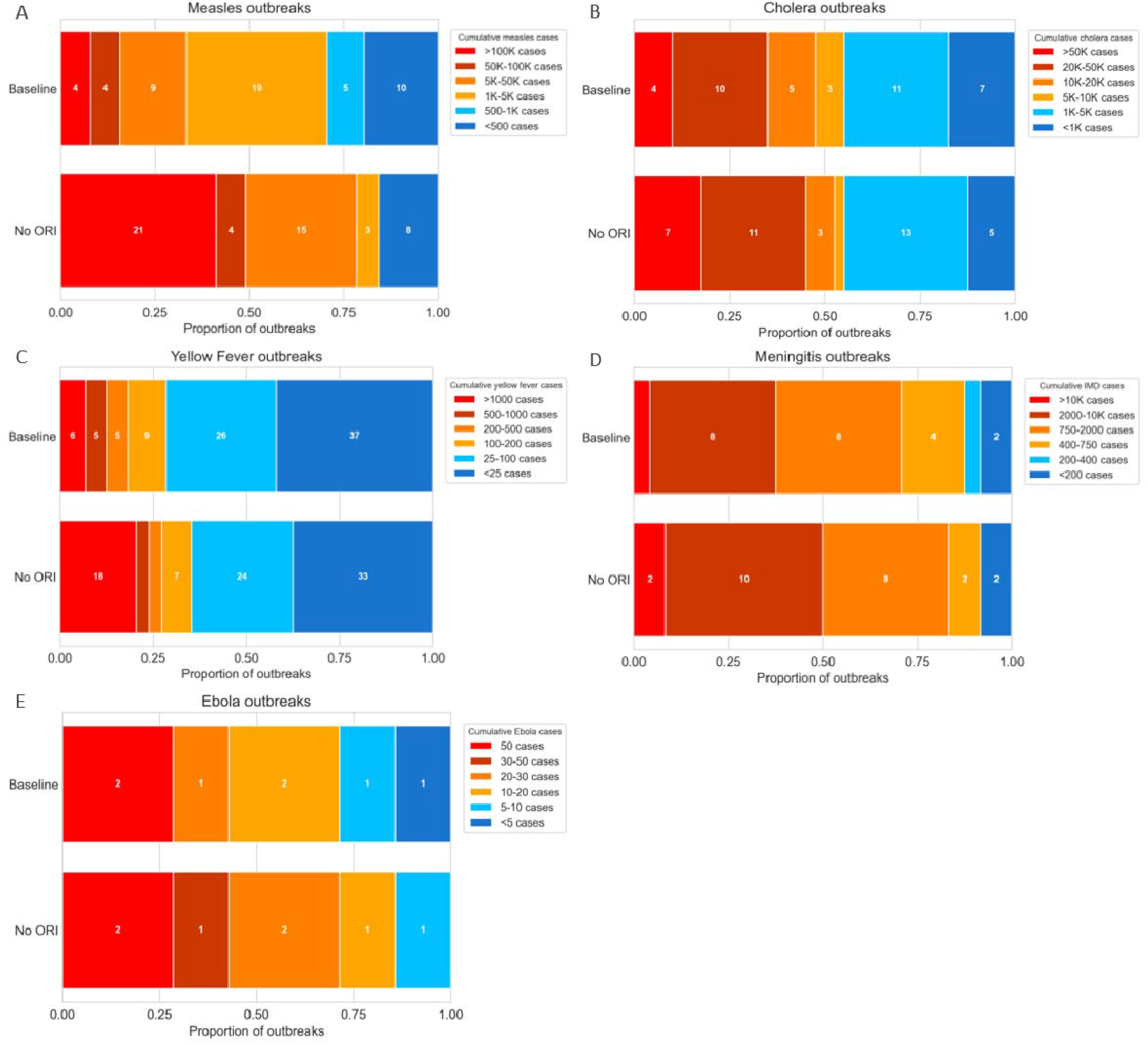
The number of outbreaks which exceed certain case thresholds, with or without ORI, for each disease. The *Baseline* plot represents where the obeserved outbreak data fall within the defined case thresholds.The *No ORI* plots represents where the median cumulative cases estimate produced by the counterfactual simulations for each outbreak falls within the defined case thresholds. Subplots A-E show the thresholds and numbers of outbreaks in each threshold for measles, cholera, yellow fever, meningitis, and Ebola. Cumulative case thresholds are shown from largest outbreaks on the left to smallest outbreaks on the right of each plot, although thresholds vary for each disease.

- Reducing the percentage of measles outbreaks with more than 100,000 cases from 41% to 8% (i.e. from twenty-one to four)
- Reducing the percentage of cholera outbreaks with more than 50,000 cases from 17.5% to 10% (i.e. from seven to four)
- Reducing the percentage of yellow fever outbreaks with more than 1000 cases from 20% to 6% (i.e. from eighteen to six)
- Reducing the percentage of meningitis outbreaks with more than 500 cases from 42% to 29% (i.e. from ten to seven)
- Reducing the percentage of Ebola outbreaks with more than 20 cases from 71% to 43% (i.e. from five to three)

## Discussion

We estimate that over the 210 outbreaks of measles, Ebola, yellow fever, cholera, and meningococcal meningitis modelled across 49 LMICs, the presence of ORI programs has prevented 327K deaths and almost $32B in societal economic costs since 2000. This is likely to be an underestimate, as the estimates do not include future impact of the vaccines provided during ORI, the societal economic costs do not include direct health system costs averted due to lack of data, and the social and macro-economic disruptions averted by preventing large outbreaks are not considered, which are known to be considerable, particularly for Ebola^16^. The burden of the diseases considered here is focused in LMICs, and the results reinforce the importance of maintaining high vaccine coverage where possible, and minimizing the time taken to initiate a vaccine response when an outbreak occurs.

For measles and yellow fever, ORI programs typically averted the largest proportional burden in settings with lower baseline vaccine coverage. However, for one of the measles outbreaks with the lowest baseline vaccine coverages (50% coverage, Democratic Republic of the Congo in 2021) the proportional impact of the ORI was surprisingly low. This is because the observed outbreak was large, likely spreading through most of the population at-risk (defined as the target population for the ORI), and the counterfactual simulations grew to a similar size, equally infecting most of the population at-risk. In reality, without ORI such a large outbreak may have spread beyond the estimated population at-risk; future work could consider this specific outbreak in detail, using comprehensive setting-specific data. For yellow fever, climate and rainfall also influence transmission risks because of their impact on mosquito populations^17-19^, and ORI was found to be proportionally more impactful in settings where environmental risks were higher.

For cholera and meningitis serogroups C, W, X, and Y, routine vaccination is not the norm due to low vaccine stock or the short duration of protection provided by polysaccharide vaccines^20,21^. For these diseases, the results indicate that the impact of the ORI is affected by the speed of the response. The shorter the time between outbreak declaration and initiation of the ORI program, the more impactful the program. This relationship is also observable for measles and yellow fever; however, it is more difficult to draw strong conclusions because of the confounding impact of the baseline immunity.

Ebola is an outlier from these conclusions as the vaccine is new and expensive^22^, and the disease itself is rare, therefore routine immunization is not recommended. However, the severity of the disease means that responses are fast and include many non-ORI elements^23,24^. While this analysis was unable to assess which aspects of the Ebola vaccine response were most impactful, previous analyses have identified that contact tracing plays a key role in outbreak response, allowing for targeted, ring-based vaccination, which can limit chains of transmission and assist with outbreak control^25^.

These results are complementary with the impacts of preventive immunisation programs, including a recent estimate that the Expanded Programme on Immunization has saved 154 million lives globally since its introduction in 1974^26^. Other analogous studies have estimated the impact of preventive vaccines for ten pathogens (including overlap with those considered here) in LMICs, and found that from 2000-2019, the presence of vaccines reduced deaths from those pathogens by 37 – 50 million^27,28^. Both preventive and reactive vaccination play important roles in outbreak containment.

We found that the key factors for reducing outbreak size in LMIC settings are high levels of vaccine coverage through routine programs, and rapid provision of vaccines upon detection of an outbreak. The success of the MenAfriVac program at reducing the incidence of meningitis A in the meningitis belt in Africa is a strong motivator for preventive vaccination for additional serogroups, and the development of a pentavalent conjugate vaccine is expected to make this a reality soon^29^, alongside the WHO recommending the use of multivalent meningococcal conjugate vaccines in routine programs in the meningitis belt^30^. However, the maintenance of high levels of immunity, particularly in LMICs, is a difficult task and outbreaks still occur when pathogens enter vulnerable or hard to reach communities.

This analysis also considered the impact ORI has had on the risk of observing large outbreaks in LMICs. Larger outbreaks result in worse health and economic outcomes, and can necessitate more disruptive public health measures such as travel interruptions or school and business closures. The outcomes from a large outbreak can also go beyond the impacts captured in this analysis, into broad and long-lasting secondary impacts such as decreases in employment, economic growth, and government spending^31^. In previous work the risk of large outbreaks was identified as a metric of interest for decision-makers^5^, and we found here that the presence of the ORI consistently resulted in smaller outbreaks (e.g., reducing the percentage of measles outbreaks with more than 100,000 cases from 41% to 8%).

There are limitations associated with this analysis, partially because of the collective calibration approach using the parameter lattice, and partially because of the level of data available to inform the disease models and outbreaks. Limitations with the calibration consistent across all diseases are outlined in Supplement 1, and limitations for the disease-specific models are provided in detail in Supplements 2-6. For the main limitations we consider whether they are likely to bias the results to be underestimates, overestimates, or to influence them in unknown ways. In the models, the same at-risk population was used for the *Baseline* and *No ORI* scenarios, however the *No ORI* scenarios are likely to have a larger population at-risk due to the possibility of additional spreading, and hence the ORI impacts are likely underestimated. Outbreak data was scaled to fit the fixed model population size, meaning that detection of some outbreaks in the simulations may have been optimistic, leading to an underestimate of ORI impact. Conversely, due to a lack of data we assumed ORI programs delivered a constant number of vaccines per day, and reached 100% coverage of their stated target population, which likely produced overestimates. Finally, due to missing data, 14 measles outbreaks were not fully calibrated and impact estimates were based on the median outbreak size at a given lattice point. For all other outbreaks, our calibration process implicitly assumed that outbreaks were close to the median model simulation. It is unclear how these factors influenced the results.

There is a large scope for future work in this space: more outbreaks could be considered with more comprehensive data, additional diseases could be considered, the methods could be extended to consider the benefits of vaccination over longer time periods, and economic impacts could be calculated to include direct health system costs.

## Conclusion

The implementation of ORI programs across 210 outbreaks of measles, Ebola, yellow fever, cholera, and meningococcal meningitis in LMICs since 2000 are estimated to have averted 5·81M cases and saved 327K lives. For diseases with routine vaccination programs, maintaining high levels of population immunity is vital for preventing large outbreaks, and when outbreaks do occur a rapid vaccine response typically provides the greatest protective impact for the population at-risk.

## Data Availability

All data produced in the present study are available upon reasonable request to the authors.

## Author contributions

Conceptualization, D.D., S.V., T.M., D.H., R.G.A. and N.S.; formal analysis, D.D. and A.M.M.; funding acquisition, N.S.; investigation (data gathering), D.D., A.M. M. and G.M.; investigation (modelling), D.D. and A.M. M.; methodology, D.D., R.G.A. and N.S.; project administration, T.M., D.H. and N.S.; software, D.D., A.M. M. and R.G.A.; supervision, R.G.A. and N.S.; validation, D.D., A.M. M., S.V., T.M., D.H., R.G.A. and N.S.; visualization, D.D. and A.M. M.; writing—original draft, D.D.; writing—review and editing, D.D., A.M. M., G.M., S.V., T.M., D.H., R.G.A. and N.S. All authors have read and agreed to the published version of the manuscript.

## Acknowledgements

The authors thank the VIMC JHU cholera modeling team for sharing data used in the cholera model, and for providing feedback and validation on the model and results. We thank Prof Andre Arsene Bita Fouda and the meningitis team at the World Health Organization Regional Office for Africa for sharing data used in the meningococcal meningitis model, and for providing feedback and validation on the model and results. We thank the eliminate yellow fever epidemics (EYE) Secretariat for sharing data used in the yellow fever model, and for providing feedback and validation on the model and results. We thank the Measles and Rubella Partnership Outbreak Response Fund Secretariat for sharing data used in the measles model. We would also like to thank Prof Mark Jit and Dr Han Fu, Prof Caroline Trotter, Dr Katy Gaythorpe and Dr Keith Fraser, and the Cholera and Vaccine Programs teams at Gavi for providing feedback and validation on the model structures and results for each of measles, meningococcal meningitis, yellow fever, and cholera respectively. Finally, we would like to thank Dr Rachel Sacks-Davis for providing suggestions and validation on some statistical methods during our analysis and Dr Jane Greig for providing insights into measles detection during outbreaks.

