## Supplementary appendices for "Estimating the historical impact of outbreak response immunization programs across 210 outbreaks in LMICs"

### **Supplement 1: Overall approach across diseases**

This supplementary section describes the general approach used to estimate the impact of outbreak response immunization (ORI) across our analyses. The standard method was to construct matching sets of scenarios in the model for each outbreak, with and without ORI interventions. For each outbreak, the impact of ORI is estimated by comparing scenario outcomes, with the overall impact per disease obtained by aggregating the impact of ORI for each individual outbreak.

#### **Imputing missing variables in outbreak data**

The model requires outbreak-specific information regarding each ORI campaign, including:

- Target population: For many of the outbreaks, a target population was available in the data. If the target population was missing, but the number of vaccines delivered or the amount funded was available, the target population was imputed assuming a linear relationship with the number of vaccines or the amount funded (Figure S1).
- Response delay and ORI duration: The response delay describes the time between the official declaration of an outbreak and the start of the ORI (i.e. the first delivery and distribution of vaccines). The ORI duration is the time from the start to end of the ORI.
  - For measles, many outbreaks included dates when the ORI was requested and approved and when it started and ended. If the ORI start date was unknown, the date of approval or request was used plus adding an average delay for the ORI start. If neither the start, approval nor request date were known or the date of the outbreak declaration was unknown, the average response delay from outbreaks in the same or a nearby country was used. If the ORI end date was not available, the ORI duration was approximated based on outbreaks in the same or a nearby country.
  - For other diseases, the ORI start and end date reported in the data were used where available. If the ORI start date, date of the outbreak declaration or end date were unknown, the average from outbreaks in the same or a nearby country were used.
- Total number of vaccines: For many of the outbreaks, the total number of vaccines was provided. If the total number of vaccines was missing, but the target population was available, the total number of vaccines was imputed using the linear relationship between the number of vaccines and the target population (Figure S1).
- Response speed: The response speed, i.e. the vaccine delivery rate, is calculated as the total number of vaccines divided by the ORI duration. These two parameters are either provided by the data or imputed following the steps from above.

Furthermore, for many of the outbreaks, vaccines were delivered in multiple phases throughout the outbreak. Multiple response phases were approximated with a single rollout, by summing the target populations associated with each outbreak, and averaging the vaccination rates.

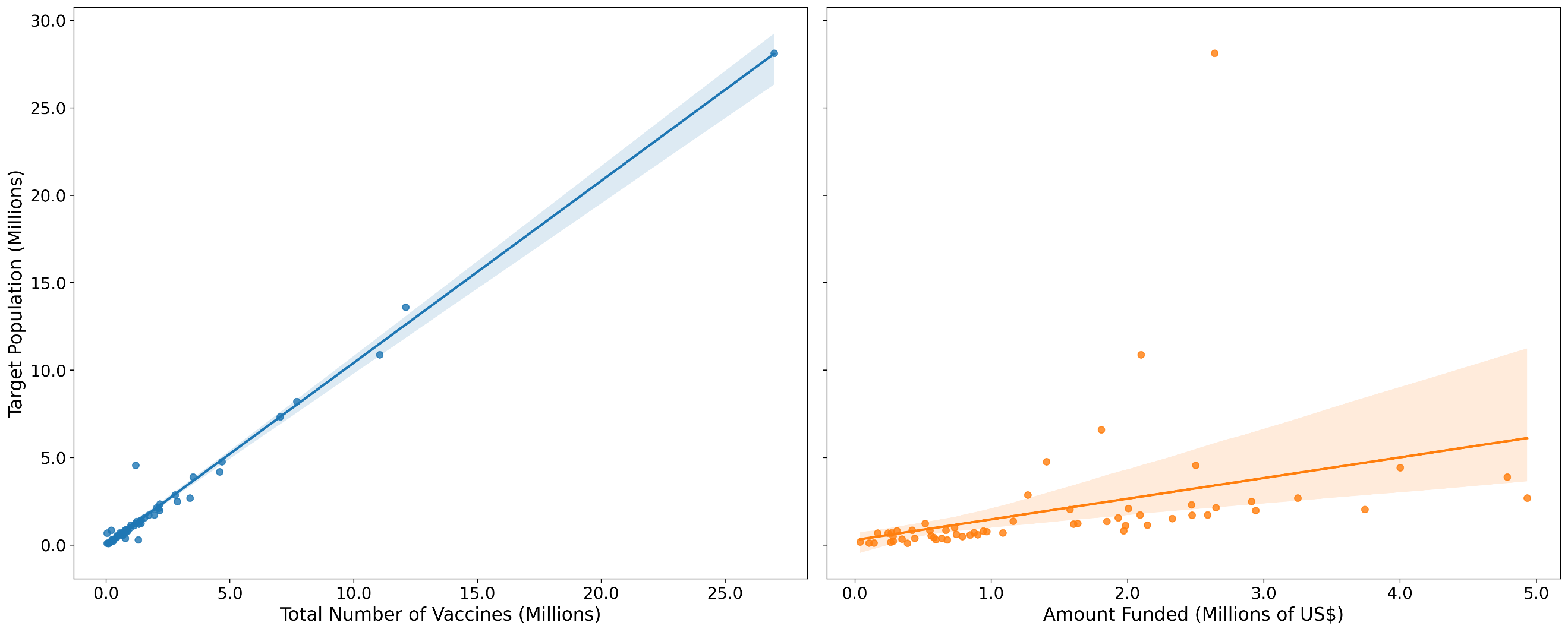

**Figure S1: Imputing outbreak data**: Scatterplot showing the linear relationship between the number of vaccines (Millions), the amount funded (Millions of US$) and the target population (Millions) using the pairwise comparison of number of vaccines and target population for each phase, across outbreaks included in the impact estimation.

#### **Exclusion criteria for some outbreaks**

For each disease, the quantity and quality of available data differed significantly across outbreaks, which allowed for only a subset of outbreaks to be included in the analysis. Outbreaks were excluded from the analysis using the following criteria:

- No ORI occurred.
- Multiple phases of the response: For many outbreaks, vaccines were delivered in multiple phases throughout the outbreak. If data was available for the second or later phases, but not for the very first phase, the outbreak was excluded.
- Insufficient data on target population: If an outbreak was missing data on the target population, and there was no data on amount funded, people vaccinated or vaccine doses delivered to impute the target population (see Imputing missing variables in outbreak data), the outbreak was excluded.
- Number of cases not representative of entire outbreak: A few outbreaks reported cases/deaths before the outbreak was declared over or before the ORI started. As this is not representative of the entire outbreak, these outbreaks were excluded.
- Long response delay: For each disease, the distribution of response delays of each ORI was considered, and outliers identified. Outlier outbreaks with long response delays were excluded. E.g. the measles and yellow fever modelling excluded outbreaks with a response delay of more than 241 days and 330 days, respectively.
- Target populations inconsistent with model structure: If an outbreak and response was localised to a highly specific setting (such as a prison) then the model’s population structure would not accurately capture the transmission dynamics and therefore these outbreaks were excluded.
- Significant disruptions to the outbreak response: If an outbreak occurred when additional factors are known to have disrupted the public health response (such as in a high conflict setting) then the implementation of the vaccine response (and other interventions such as contact tracing) in the models would not be able to capture the impact of these disruptions. As such these outbreaks were excluded.

#### **Starsim framework**

The *Starsim* framework was used to build the models in this study to provide a single common software and analysis architecture for all the models^1^, while simultaneously having the flexibility to account for very different modes of transmission and available interventions across the diseases being modelled. Key components of *Starsim* include:

- Disease ‘modules’: capturing different health states and disease progression within individuals.
- Contact networks: to simulate interactions between agents that can give rise to transmission. Multiple networks can be used to capture different transmission mechanisms and different settings (e.g., household networks, school networks, random community contacts).
- Interventions: dynamic throughout model simulations, and can include delivering vaccines, performing contact tracing, or changing transmission levels due to non-pharmaceutical interventions that are brought in following outbreak detection.

Alongside transmission mechanisms, population demographics, and interventions, *Starsim* also has functionality to simulate vital dynamics and pregnancies, although these features were not used in the present study due to the relatively short duration of outbreaks.

#### **Model population and target population scaling**

The size of counterfactual outbreaks without ORI can be sensitive to the total population size in the model, which can limit the extent to which an outbreak can grow. For example, without ORI would an outbreak have spread to neighbouring geographical areas. The size of counterfactual outbreaks is likely to depend on many factors including mobility patterns for the specific local areas in which an outbreak took place, as well as behavioural changes and other health system responses that may change as the size of an outbreak increases. In the absence of data to inform these factors, we assume uniform spread with no behavioural changes or other interventions, with the maximum possible size of the outbreaks corresponding to the population size in the model. In the absence of detailed estimates of the at-risk population, we use the target population for outbreak responses as a proxy.

Due to computational limitations, simulations were all run with a fixed number of agents (50,000 or 100,000 agents). Outbreak data was scaled onto this population based on the target population size for each outbreak, with the exception of the Ebola outbreaks. For example, if an outbreak totalling ten thousand cases occurred in an at-risk population of one million people, our model only has a fixed population of fifty thousand agents so we scaled each agent to represent 20 people and calibrated to an outbreak size of 500 cases in the model. Calculated ORI impact in the model was then scaled back up to the original outbreak size for the purpose of estimating the overall impact for each outbreak.

#### **Outbreak declaration and response threshold**

For all diseases except meningitis, the threshold for outbreak declaration in the models was defined as one case. This may be optimistic because the models do not distinguish between suspected and confirmed cases, and so does not consider delays related to laboratory confirmation requirements. Conversely, it may be pessimistic because due to the population scaling factor that is applied to the measles, meningitis, yellow fever, and cholera models, one case being detected in the model typically represents multiple cases in the real world.

These factors do not impact the Ebola model as it does not scale the outbreak size to the model population and therefore one case in the model reflects one observed case. The meningitis model uses a separate definition of five detected cases within a week for outbreak declaration, aligning with the threshold of 10 cases per 100 thousand population used for districts larger than 30 thousand people.

Additional assumptions specific to some models may also impact these thresholds. The measles model only includes children aged 0-5 years, and so background cases which may occur in the older population could contribute to the outbreak detection threshold. The meningitis model is initialised with an assumed background prevalence of asymptomatic disease built up over the endemic period of the year, and it is likely that background cases of invasive disease would have occurred as well.

#### **Assigning outbreaks to lattice points**

For quantities such as the response time and response speed, values (or imputed values) are available as part of the input data, and outbreaks can be directly assigned to these dimensions of the lattice. In the case of the disease-specific free dimensions described above (vaccine coverage for measles, mosquito transmission level for yellow fever, and asymptomatic carrier prevalence for meningitis), data were not available to inform parameter values in the model. Therefore, these quantities were estimated as part of model calibration. For each disease at most one lattice dimension is left as a free variable. After assigning an outbreak to all lattice dimensions apart from one (if necessary), the final lattice dimension was assigned by a maximum-likelihood estimate of the total number of cumulative cases in the outbreak.

#### **Kernel Density Estimation**

To assign the final lattice dimension by the maximum-likelihood estimate of the total number of cumulative cases in the outbreak, we estimated the probability density function (PDF) of each lattice point using a gaussian kernel density estimator. The choice of the kernel bandwidth has a great impact on the shape of the generated PDF and due to the large range of outbreak sizes associated with each disease, the bandwidth was chosen as a proportion of the median model outcome for each lattice point. This way, we avoid under-fitting of smaller outbreaks (if the bandwidth is too wide) and over-fitting of larger outbreaks (if the bandwidth is too narrow).

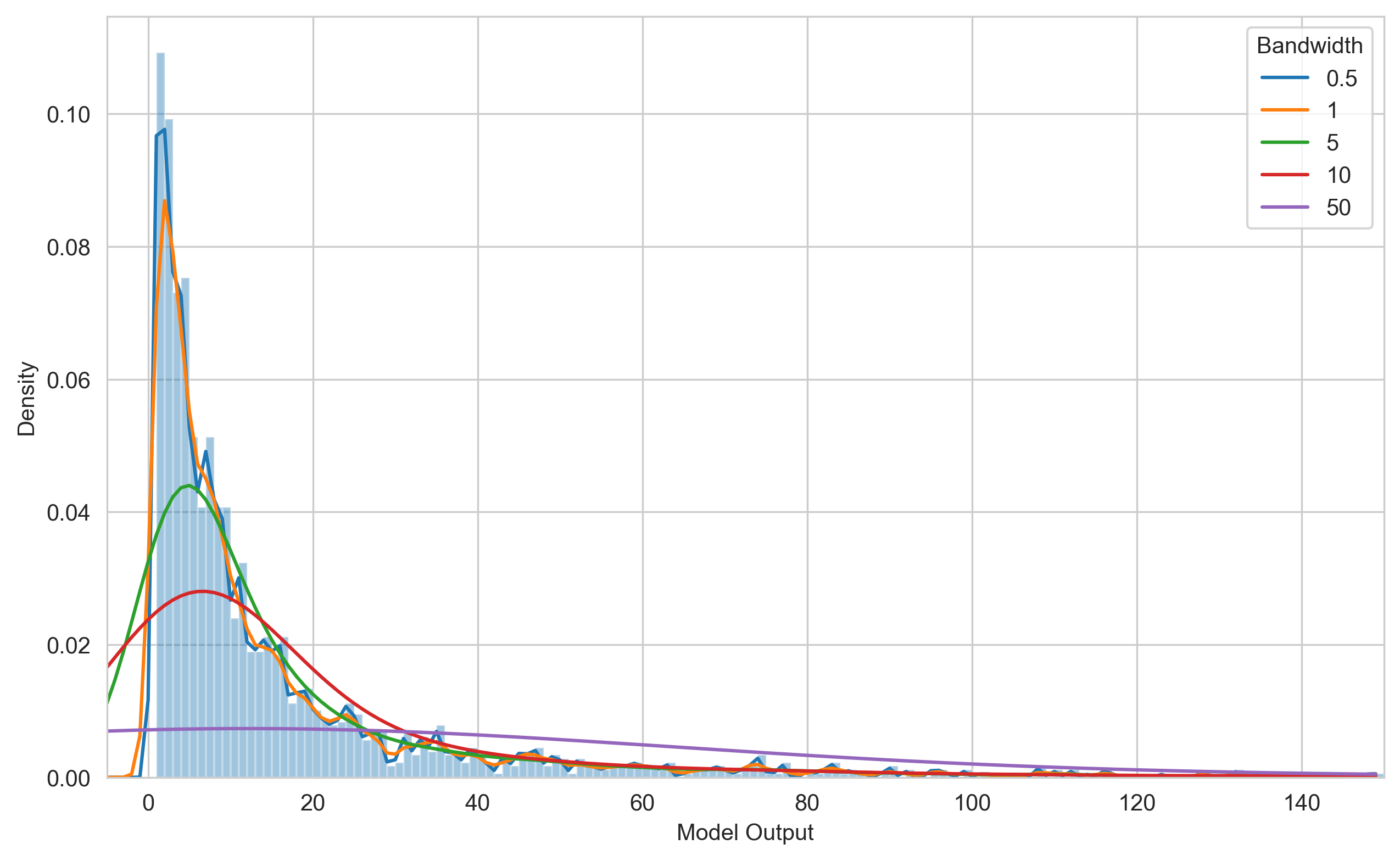

**Figure S2: Kernel Density Estimation**: Example histogram of model output with corresponding kernel density estimate for different bandwidths. Smaller bandwidths lead to over-fitting (high variance), whereas too wide a bandwidth leads to under-fitting (high bias).

#### **Trajectory selection**

In some cases (particularly for small outbreaks as seen in Ebola), stochastic variability early on in the outbreak can strongly influence the trajectory of an outbreak, even before an outbreak response takes place. For example, a small number of transmission events early on may result in an outbreak ‘fizzling out’ without requiring any response. Alternatively, rapid initial transmission could lead to a much larger outbreak, even with the same model parameters. For a given set of parameters, the model will produce a spectrum of possible outbreak trajectories, of which the actual outbreak is just one realization. If the entire spectrum of simulations is included, many of the simulations will have a considerably different number of cases to the actual observed outbreak. This introduces a large amount of uncertainty into the estimated impact, but this uncertainty arises from simulations that are not consistent with the outbreak data.

An additional complicating factor is the impact of stochasticity in the model. A small outbreak with ORI may be accounted for by a low-risk sequence of initial transmission events. If the ORI was taken away, this same initial sequence of transmissions could still result in a small, self-limiting outbreak. The delay in vaccine response means that simulations with the same random seed in the model will have the same sequence of transmission events up to the point where the vaccine response begins. In considering the counterfactual of ORI vs no-ORI, it is important to preserve the initial sequence of transmissions so that the level of risk prior to ORI commencing is the same in both cases. This is achieved through ‘trajectory selection’ or simulation ‘filtering’ which takes place in three steps:

1. A set of simulations is produced for each lattice point/set of parameters examined, containing the full range of possible model outcomes.
2. For each outbreak, this set of simulations is filtered to identify simulations that lie within a threshold distance (in terms of number of cumulative cases) to the observed number of cases. Decreasing the threshold distance will decrease uncertainty in the estimated impact arising due to mismatches between the simulations and the data. However, given the finite number of simulations conducted, this will also reduce the number of simulations used to produce the impact estimate. An insufficient number of filtered simulations may not provide an adequate sample size to estimate impact – typically we target at least 100 simulations per outbreak. Selection of the threshold distance balances these two competing factors. In this study, we use a threshold distance of +/- 25%.
3. A matched set of counterfactual simulations is obtained by running the model with the same random seeds selected in the previous step, but without any ORI. These simulations are ‘matched’ in the sense that they are identical (with the same transmission events and number of cases) up until the point that ORI begins. The ORI impact is obtained by examining the distribution of pairwise impact between matched simulations (i.e., using the distribution of difference in cases with/without ORI, as opposed to comparing the distributions of cases with/without ORI).

Figure S3 illustrates the steps above. Two example outbreaks (shown as the green and orange crosses) are assigned the same lattice point, for which a set of simulations is produced, containing the full range of possible model outcomes (blue boxplot, left). The green and orange boxplots on the left reflect the subset of the full model outcome after identifying simulations that lie within 25% to the observed number of cases. The matched sets of counterfactual simulations in comparison to the full range of counterfactual model outcomes are shown on the right, indicating that for the same setting, larger outbreaks produce larger counterfactual outcomes than smaller outbreaks.

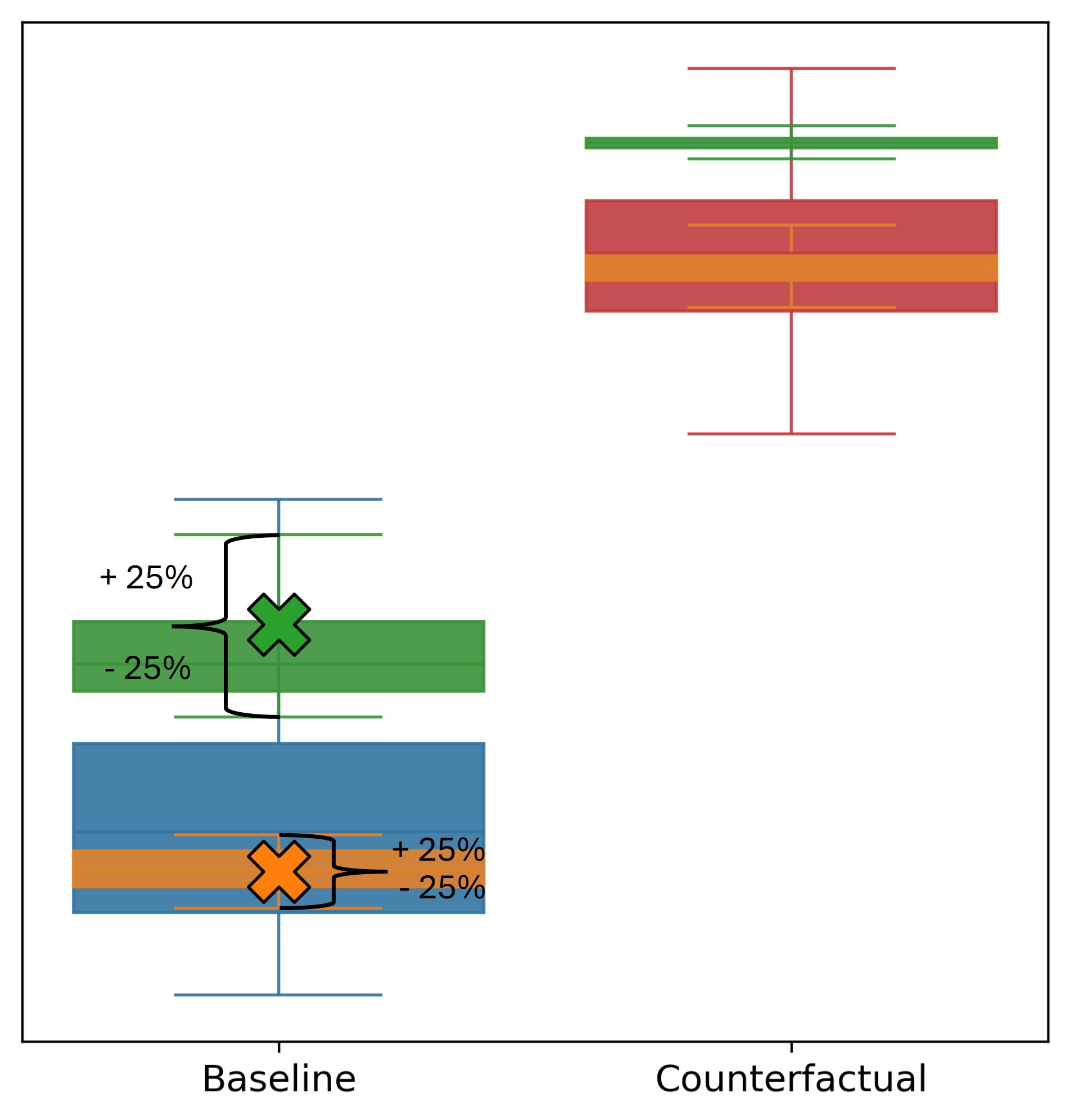

**Figure S3: Trajectory Selection Methodology**: The left boxplots show the full baseline model output (blue) and the model output filtered to +-25% of cumulative cases of two example outbreaks (green and orange). The right boxplots show the full range of counterfactual model output (red) and the matched counterfactual output for the two example outbreaks (green and orange). Crosses indicate cumulative cases of the two example data points.

It should be noted that stochastic dynamics in the model means that even with a very small threshold, the model will still produce a range of counterfactual outcomes depending on how sensitive the outcomes are to the ORI. For example, with very late ORI there may be little impact because many infections took place prior to the response and therefore the trajectory selection mainly reflects pre-ORI transmission events resulting in a very narrow range of outcomes for the counterfactual. In contrast, with very early ORI the trajectory selection could mainly reflect stochastic dynamics after the ORI, and therefore the counterfactuals associated with the selected trajectories may still have a wide range of outcomes. Therefore, selection of a threshold distance does not directly ‘set’ the distribution of counterfactual outcomes, which still reflect sensitivity to ORI.

It is important to note that the use of trajectory selection depends on the specific question being asked. As we are focused on ‘how many cases were averted in the historical outbreak’ trajectory selection is required to select stochastic realizations from the model that are consistent with the historical outbreak. In contrast, if the question was ‘how many cases would be averted for similar outbreaks’ then trajectory selection would not be performed, because it is important to retain the full range of possible model outcomes.

#### **Disability-adjusted life years (DALYs) averted**

Using the estimates of cases and deaths averted by ORI for each outbreak across disease models, disability-adjusted life years (DALYs) averted can be estimated. This is done by multiplying cases averted by the years of healthy life lost due to disability (YLDs), plus years of life lost (YLLs) from deaths (for each outbreak, the average life expectancy in that country and year^2^ compared to the age of deaths in the model). Disability weights^3^ for acute conditions were estimated only for symptomatic cases in each model, and assumed to last for the average duration of infection. For disability weights from chronic sequelae, incidence was disease-specific and assumed to be life-long unless otherwise stated. Details for these parameters for each disease are available in the ‘Model overview’ section of Supplements 2-6.

#### **Socio-economic costs averted**

Socio-economic costs averted by ORI were calculated by using a human capital approach, with the method being similar to Watts et al.^4^. Productive years of life lost or lived with disability were multiplied by GDP per capita, with productive years of life lost estimated as the difference between the average age of death in the model and a retirement age of 64, or the life expectancy in that country and year^2^, whichever was lower. To account for mortality and morbidity in people younger than 15 years (the assumed minimum working age) we estimated the proportion of model population which were younger than 15, multiplied by the probability of survival to age 15^2^, and only counted DALYs accrued after reaching age 15. We used GDP per capita from the corresponding country and year^5^, with future costs discounted at 3% per annum as a standard method. Costs were then inflated to 2023 US$. All estimated impacts are calculated only over the time spans of the modelled outbreaks and do not account for the benefits of vaccination or immunity via infection on potential future outbreaks.

#### **Categorising outbreak sizes**

Using the total suspected cases observed for the set of historical outbreaks we defined a series of outbreak size thresholds for each disease, assigned each outbreak to its appropriate threshold range, and produced a distribution of outbreak sizes for each disease. In order to compare how the distribution of outbreak sizes changes when ORI is not implemented in the counterfactual scenario, we examined the median model estimated number of cumulative cases for each outbreak without a vaccine response and assigned each outbreak to the appropriate threshold range. This allows us to estimate the frequency with which the presence of ORI prevented outbreaks from exceeding certain total case thresholds.

#### **Analysis steps**

For each disease, the analysis followed a similar sequence:

1. Design a model incorporating key aspects of transmission and progression for each disease. The models are all developed with the same agent-based model framework, but differ considerably in structure due to factors like differences in the ages of people affected, mode of transmission (direct, environmental, vector-borne), whether asymptomatic carriage is possible, and whether diagnosis impacts outcomes (e.g., whether it prevents transmission due to quarantine or is used as a basis for ring vaccination strategies).
2. Collate outbreak datasets from available sources and impute any missing outbreak variables.
3. Produce a ‘lattice’ containing a distribution of model outcomes for a grid of pre-determined parameter values, assigning outbreaks to each lattice point.
4. Calibrate the model by
   a. Incorporating/fitting to any data available. This step can differ greatly across disease areas depending on what data is available.
   b. Assessing the size of the outbreaks in the data relative to the corresponding lattice points, ensuring that the calibration produces realistic outputs.
5. Perform trajectory selection to produce sets of simulations that are filtered to match each outbreak’s total reported cases.
6. Run counterfactuals and estimate cases/deaths/DALYs/costs averted by the ORI for each outbreak.

#### **Limitations**

- Outbreak detection thresholds using a scaled model population: For four of the five diseases, a model population of 50K is used to represent outbreaks that can occur in much larger settings, and hence a single case in the model can represent dozens of cases. This means that a threshold of one case for outbreak declaration in the model does not truly represent only a single case, which may delay outbreak detection and responses in the models, underestimating ORI impact. However, this is partly mitigated through model calibration steps.
- Not accounting for direct health system costs: The estimated economic costs averted that we produce only represent societal costs based on years of life lost and years of healthy life lost due to disability; we do not produce estimates of direct health system costs averted by ORI due to lack of data. This means that we are underestimating the true economic costs averted by the presence of ORI across all outbreaks considered.
- Epidemiological and response data were often missing: The set of outbreaks which we calibrate against for each disease, and use to produce our estimates of impact averted were predominantly informed by data from publicly available sources, and these data were often incomplete. This means that we had to impute epidemiological or response data for a subset of outbreaks, or use model-estimated cases/deaths if imputation was not feasible.
- Bias when calibrating to observed data: During calibration Baseline simulations are selected to be centered around observed outcomes. Implicitly this assumes that the observed outbreaks are instances of the most likely outcomes, given the epidemiological and demographic setting. In reality, each observed outbreak may have been particularly large or small relative to a most likely outcome, which could lead to over- or under-estimating the impacts. As there is no data to inform the likelihoods for each observed outbreak, it is unclear what overall impact this has on the results.
- Impact of random number generation: High stochasticity is a key feature of agent-based models, where initial conditions and random number generation during transmission calculation can have large impacts on outbreak trajectory. This is particularly noticeable for small outbreaks, such as the ones calibrated against here. In each Baseline scenario the stochastic impacts are truncated by the simulation filtering, and we retain only simulations which are close to the observed outbreak size. However, in the counterfactual scenario the lack of an ORI changes the random number generation used for the transmission calculation. This can lead to counterfactual simulations which have no vaccine response yet produce fewer infections and deaths than their Baseline equivalent. This is not a frequent outcome, but it likely leads to an underestimate of the ORI impact across our disease models.
- Potential issue with counterfactual size in early responses: If the estimated target population of an ORI is small because the response is fast, we might underestimate the size of the counterfactual and therefore underestimate the value of the vaccines. The large response sizes for all reported outbreaks suggest however that responses are typically broad even for small outbreaks, reducing the risk of this being an issue.
- Multi-phase responses: For outbreaks where the response had multiple phases, no data was available on the timing of the cases (e.g., if multiple responses were due to multiple waves of infections) nor on the specific locations of the responses (e.g., if the responses were in different cities within the same subnational region). As an approximation, we grouped and averaged the responses by considering the target population for the outbreak to be the sum of the target populations for all the responses applied to that outbreak and modelling a single vaccine response with a vaccination rate equal to the average of vaccine rates for each individual response.

### **Supplement 2: Measles**

This supplementary section describes the context and more detailed methods for our measles model and the ORI impact analysis.

#### **Background and motivation**

Measles is a highly contagious, serious airborne disease caused by a virus that can lead to severe complications and deaths. Measles is so contagious that if one person has it, up to 90% of the people close to that person who are not immune will also become infected. Symptoms include a high fever, cough, runny nose, and a rash all over the body. The highest burden is in unvaccinated or under vaccinated children under the age of five. The WHO states that measles vaccination averted 56 million deaths between 2000 and 2021. However, in 2022, only about 83% of the world’s children received one dose of measles vaccine by their first birthday through routine health services – the lowest since 2008^6^.

#### **Model overview**

The *Starsim* framework was used to create an agent-based model of measles among humans^1^, with states for susceptible, exposed, infected, and recovered agents (Figure S4).

Agents in the model represent humans, who begin as susceptible, and each day have a probability of becoming infected that depends on their immunity status and is proportional to the prevalence of infection among the population. Following infection, humans enter a latent infection ‘exposed’ state, before becoming infectious to others. Humans in the infectious state can recover and develop immunity to further infection during the modelled outbreak, and humans can die based on a disease-specific mortality rate. Agents can also be vaccinated, which will provide a level of immunity against future infection.

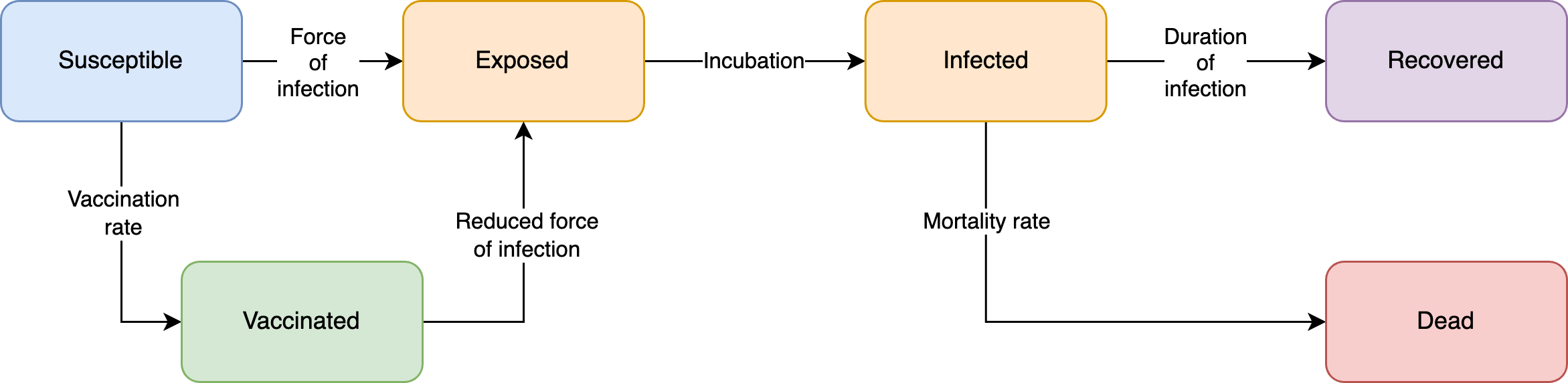

**Figure S4: Measles model schematic**. The Starsim framework was used to develop an agent-based model of measles among humans (S-E-I-R).

**Table S1: Measles model parameters and sources.**

| Parameter | Value | Source / notes |
| --- | --- | --- |
| **Population parameters** | | |
| Population age distribution | Empirical distribution | United Nations, Department of Economic and Social Affairs, Population Division^2^; averaged over countries with outbreaks used in this analysis. |
| Mean ‘community’ contacts per day | 3·5 | Prem et al.^7^; averaged over all non-household contact rates and 0–5-year-olds for LMIC countries. |
| **Disease parameters** | | |
| Probability of death, given severe disease | 0·5% | Calibrated value. |
| Average duration of exposed period | 9-12 days | Measles is infectious 4 days before the rash onset, which follows 2-4 days after prodromal symptoms that appear 11-12 days from exposure^8^. |
| Average duration of infection | 8 days | Measles is infectious 4 days before and 4 days after the rash onset^8^. |
| Measles Testing Probability | 10% | Assumption. |
| Isolation Factor | 0·7 | Assumption based on mild isolation in LMICs. |
| Vaccine protection against infection | 83%; one dose | Based on vaccine efficacy for >9months olds. Vaccine efficacy increases linearly over time and reaches peak of 83% after 10 days ^9^. |
| Maternal Immunity | Age calculated in the model | Age calculated as weighted average of 3·78 and 0·97 months based on proportion of naturally infected and vaccinated mothers^10^ |
| **Health economic parameters** | | |
| Disability weights for yellow fever infection | Moderate: 0·051  Severe: 0·133 | Global Burden of Disease (2017) Disability Weight estimates^3^. |
| Average life expectancy | Specific to country and year of outbreak | United Nations, Department of Economic and Social Affairs, Population Division^2^. Used to estimate years of life lost for each death. |
| Gross Domestic Product (GDP) per capita* | Specific to country and year of outbreak | World Bank, World Development Indicators^5^. |
| Discounting | 0% for DALYs; 3% for costs |  |

*Inflated to 2023 USD using average annual inflation rates since 2000^11^.

#### **Outbreak data**

Measles outbreaks have been recorded by the WHO in its *Weekly Epidemiological Record* and *Disease Outbreak News* since 2000, with information on epidemiological and programmatic responses available online^12^ and additional response information provided by Gavi.

Between 2000 and 2023 there were 236 recorded outbreaks. 74 outbreaks were recorded as having received a vaccine response, and 51 outbreaks had sufficient response data for inclusion in the analysis, 63% of which were in Africa. Data on the ORI target population size and number of cumulative cases per outbreak were the most complete, while information about the ORI response time and duration of the outbreak were the most incomplete (Table S2). The recorded outbreaks varied widely in scale (40-1M cumulative cases), vaccines delivered (44k-12M) and time to respond (3-274 days).

**Table S2: Summary characteristics of outbreak data used for measles analysis.**

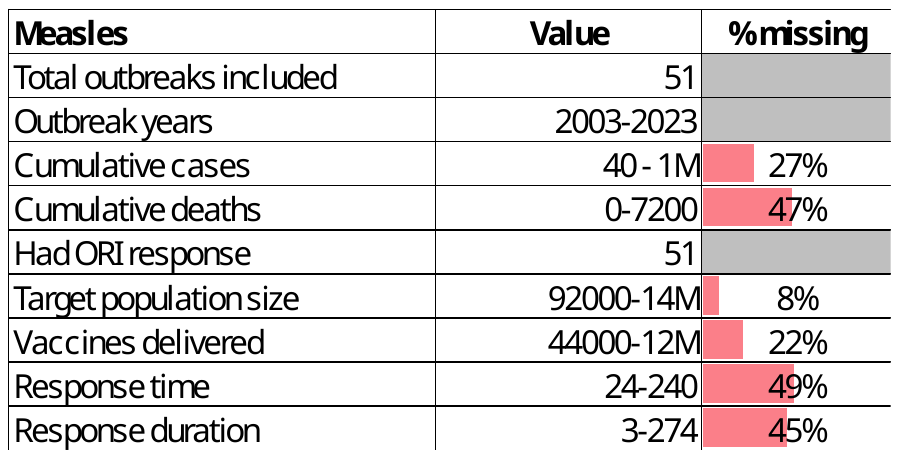

Some model parameter values were set based on the following quantities from the outbreak dataset, and imputed where necessary (see Imputing missing variables in outbreak data):

- Target population size: Directly from data, otherwise estimated based on number of vaccines (using linear relationship between number of vaccines and target population, fitted to all data with a target population and number of vaccines).
- Response time: Directly from data (time between outbreak declared and vaccine response started) if available, otherwise the average response time from outbreaks in the same or nearby country.
- Vaccine doses delivered: Directly from data where available, otherwise estimated based on target population size (using linear relationship between target population size and number of doses).
- Response duration: Directly from the data where available, otherwise based on average response duration from outbreaks in the same or nearby country.
- Response rate of vaccination: Calculated based on number of doses and response duration.

The lattice of parameter values for which the model was run was obtained by binning these variables, as well as the baseline vaccine coverage, into the following levels:

- Response time: 30, 60, 90, 120, 170, 220 days
- Vaccination rate: 500, 2500, 5000, 7000, 8500 vaccines per 50,000 population per day
- Baseline vaccine coverage: 10-100%: assigned through calibration process described below

The baseline vaccine coverages in the model describe the proportion of the population that is seeded as vaccinated. A baseline vaccine coverage of 100% implies that the entire proportion of the population eligible for a vaccine is seeded as vaccinated. Additionally, the model considered maternal immunity for infants, which is described in more detail below.

#### **Model population for simulated outbreaks**

The model population for each outbreak simulation represents the population of the specific geographic location where the outbreak occurred, based on the ‘target population size’ from the outbreak response data (i.e. people identified as being eligible for vaccination after the outbreak was declared). For computational reasons the model contains a maximum of 50,000 agents, and so for outbreaks with larger population sizes a scaling factor was used such that one agent in the model represents multiple people, based on the target population size and the proportion of the population eligible for the vaccine.

Measles outbreaks predominantly occur in children between 0 and 5 years old, as older people typically have immunity acquired either from vaccination or from prior infection^6^. As the ORI program target population is also limited to children under 5 years of age, we elected to only model this age group. The model was parametrized by age structure from the United Nations, Department of Economic and Social Affairs, Population Division^2^ for ages 0 to 5 years. Agents in the model are assigned integer ages, so to capture the vaccine eligibility of children >9 months, 25% of <1-year olds were assumed eligible (i.e. assuming a uniform distribution of age within this group).

Additionally, the model considers protection against measles at birth for infants up to a certain age. It is estimated that the presence of maternal antibodies endured for a median of 2·61 – 3·78 months for infants of naturally infected women and 0·97 months for infants of vaccinated women^10^. Using the assumed baseline vaccine coverage in the model, a weighted average was calculated to estimate the age where infants are no longer considered immune. It was then assumed that maternal immunity decreases linearly up to that age.

Transmission in the model occurred through community contacts between agents. These networks are randomly generated at each time step (representing a day in the model). Vaccine coverage, transmission risk and disease outcomes were not modelled to vary by age given limited data from the outbreak settings. As we only model the 0–5-year-old population, some transmission between agents is indirect and mediated by older people is not captured in the model, and instead assume it makes up a proportion of the direct transmission used in the model.

#### **Diagnosis of cases, outbreak declaration and ORI**

Agents within the model are assumed to seek a test when symptomatic, with a probability of 10% per day that they are symptomatic. Measles outbreaks are typically declared if there are five or more epidemiologically linked cases^13^. As we simulate a scaled version of the population with a limited age range and do not model background cases, an outbreak is declared in the model after the diagnosis of a single agent (see Outbreak declaration and response threshold), and once this occurs the ORI will begin after N days, where N is the response time for a given outbreak. Some people with a diagnosed infection will undergo an isolation or otherwise limit their contact with others to reduce risk of onward transmission. However, for measles it is expected that effective isolation will be challenging, so we only include a small reduction in transmission for agents that are diagnosed.

#### **Calibration**

Calibration involves estimating the transmissibility of measles in the model to produce outbreaks of a sufficient size, as well as the probability of death given an infection in order to capture the observed case fatality rate.

For each outbreak, an estimate of baseline immunity is also required, which can be challenging as baseline immunities can vary significantly across subnational regions of a country for various reasons. Eight of the 51 outbreaks included in the analysis reported a subnational area, including one outbreak with a reported subnational baseline coverage. For the remaining seven outbreaks with a known subnational area, IHME^14^ modelled estimates of the MCV1 vaccine coverage (mean and confidence interval) were used to approximate the underlying baseline vaccine coverage for each outbreak.

The outbreaks with known baseline vaccine coverage were used to estimate the transmission parameter as follows. For each outbreak, simulations were run for a range of initial baseline vaccine coverage and transmission parameters (and corresponding response time and vaccination rates fixed based on the data). For each value of the transmission parameter, a model-estimated baseline vaccine coverage was derived for each outbreak based on the maximum-likelihood estimate of the total number of cumulative cases (see Kernel Density Estimation). The transmission parameter that was chosen was the one that minimized, across the 8 outbreaks with known subnational coverage, the difference between the known subnational coverage and the model-estimated vaccine coverage (Figure S5).

For other observed outbreaks with no reported subnational area, simulations were run with corresponding response times and vaccination rates from the data, the transmission parameter estimated as above, and different levels of baseline coverage. The corresponding baseline coverage was estimated based on the maximum-likelihood estimate of the total number of cumulative cases in the outbreak (see Kernel Density Estimation).

All other relevant model parameters were constrained by estimates from the literature.

Once parameters were estimated, outbreaks could be simulated for a range of lattice points representing different response times (30, 60, 90, 120, 170, or 220 days), vaccination rates (500, 2500, 5000, 7000, or 8500 vaccines per 50,000 population per day) and baseline vaccine coverages (10-100% among the eligible population). The set of 51 outbreaks with sufficient data could then be assigned to their nearest lattice point (Figure S6; boxplots representing the range of simulated outcomes and crosses representing the observed outbreaks where they have occurred). Please note that 27% of outbreaks were missing data on cumulative cases and are therefore not shown in Figure S6.

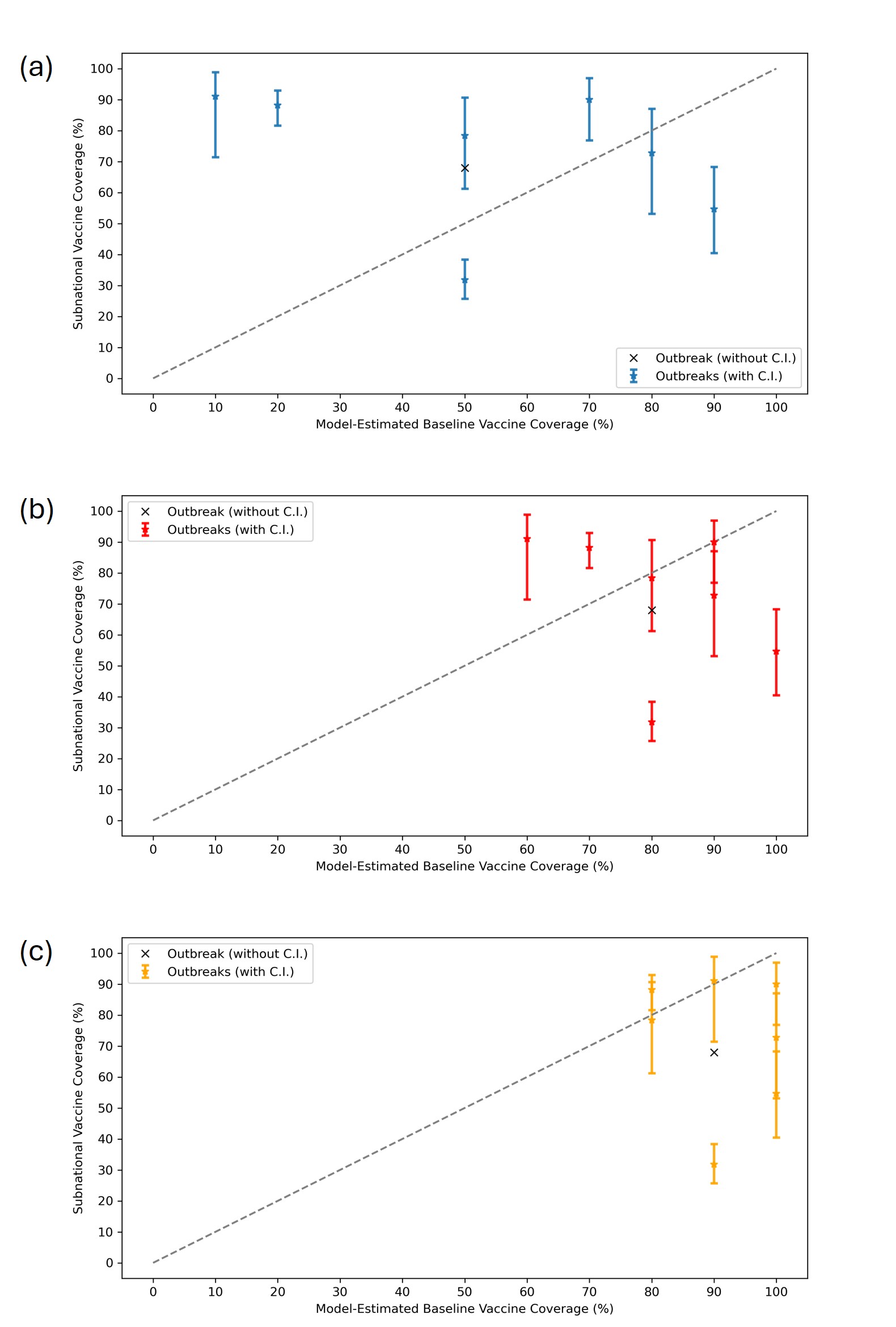

**Figure S5: Measles calibration of transmission rate.** Simulations with response time, vaccination rate and baseline coverage were run for different transmission parameters. Model-estimated vs. subnational vaccine coverage were then compared, and the final transmission parameter selected to minimize difference between model-estimated and subnational baseline coverage. The transmission rate in (a) is too low as a subset of the outbreaks are assigned to baseline coverages that are too low compared to the subnational coverages. The transmission rate in (c) is too high as many outbreaks are assigned to baseline vaccine coverages that are too high compared to the subnational coverages. The transmission rate in (b) minimizes the difference between subnational and model-estimated coverages. Bars indicate upper and lower values of IHME subnational MCV1 coverages, where available.

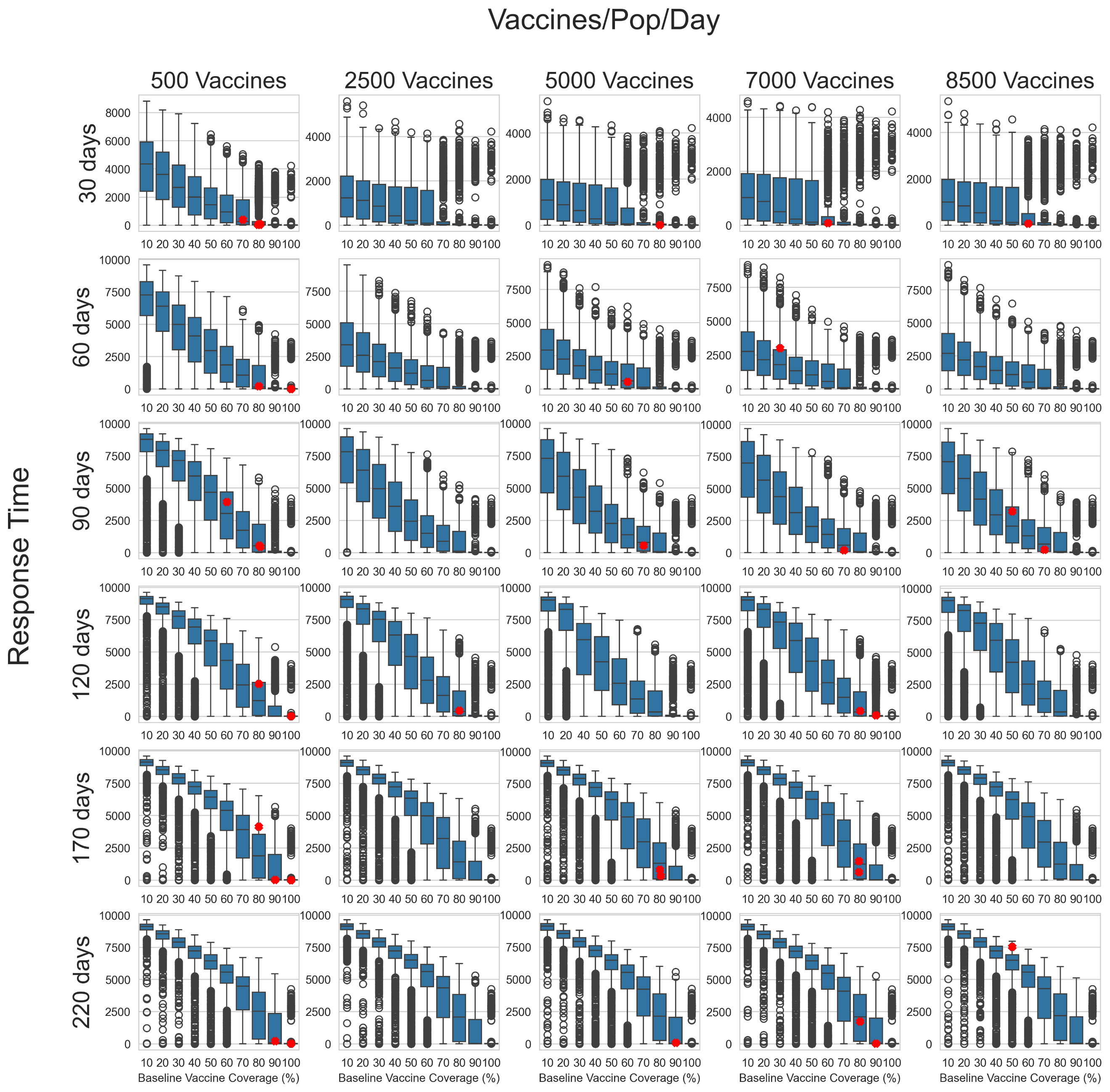

**Figure S6: Lattice of measles simulations before outbreak filtering was applied.** Each boxplot represents the distribution of total cases from simulations run with response parameters defined by the row/column lattice point. The red crosses represent the cumulative suspected cases from observed outbreaks which have been assigned to the lattice point and scaled to a population of 50K. Please note that 27% of outbreaks were missing data on cumulative cases.

#### **Scenarios**

For the purpose of running counterfactuals specific to the historical outbreaks, for each outbreak in the data with a reported number of cumulative cases, the 2000 simulations associated with the lattice point that the outbreak was assigned to were filtered to retain only those simulations where the cumulative cases were within +/- 25% of the reported cases (see Trajectory selection) For all other outbreaks, the entire range of the model outcome was considered. Two scenarios were then examined:

- Baseline: ORI as occurred, using the filtered simulations from the lattice
- No ORI counterfactual: Simulations were run without any ORI intervention using the same filtered simulation seeds from the base Baseline line scenario, such that everything was identical up until the date the ORI would have started.

#### **Outcomes**

For each outbreak and scenario, the distribution of cumulative cases and deaths across selected model simulations were recorded. The mean difference in outbreak size between scenarios and associated uncertainty in the mean were estimated from these collections of simulations using bootstrap resampling (i.e. for each outbreak to produce estimates of cases averted by ORI).

Total cases averted by ORI across all historic outbreaks was then estimated by aggregating the cases averted for each individual outbreak. As the outbreaks are independent, this was obtained by summing the cases averted per outbreak, with the variance estimated by summing the variances from each individual outbreak.

DALYs averted by ORI were estimated by multiplying cases averted by the disability weight per case and average duration of symptoms, plus years of life lost from deaths (for each outbreak, the average life expectancy in that country and year compared to the age of deaths in the model). Socio-economic costs averted by ORI were calculated by estimating productive years of life lost or lived with disability, and multiplying this by GDP per capita. Productive years of life lost were estimated as the difference between the average age of death in the model and a retirement age of 64, or the life expectancy in that country and year, whichever was lower. Costs were inflated to 2023 USD, with future costs discounted at 3% per annum as a standard method.

The impact of ORI on reducing the risk of large outbreaks was also estimated, by comparing the distribution of cumulative cases across outbreaks in the data to the distribution of cumulative cases across outbreaks in the no ORI scenario. For the purposes of this sub-analysis, cumulative cases in the no ORI scenario for each outbreak were approximated as the median from the no ORI counterfactual simulations.

#### **Results**

##### *Baseline*

The modelled cumulative cases and deaths for the Baseline scenario is shown for each outbreak considered in the analysis in Figure S7. Outbreaks are grouped and coloured by baseline vaccine coverage. The data points are scaled to the vaccine-eligible population in the model.

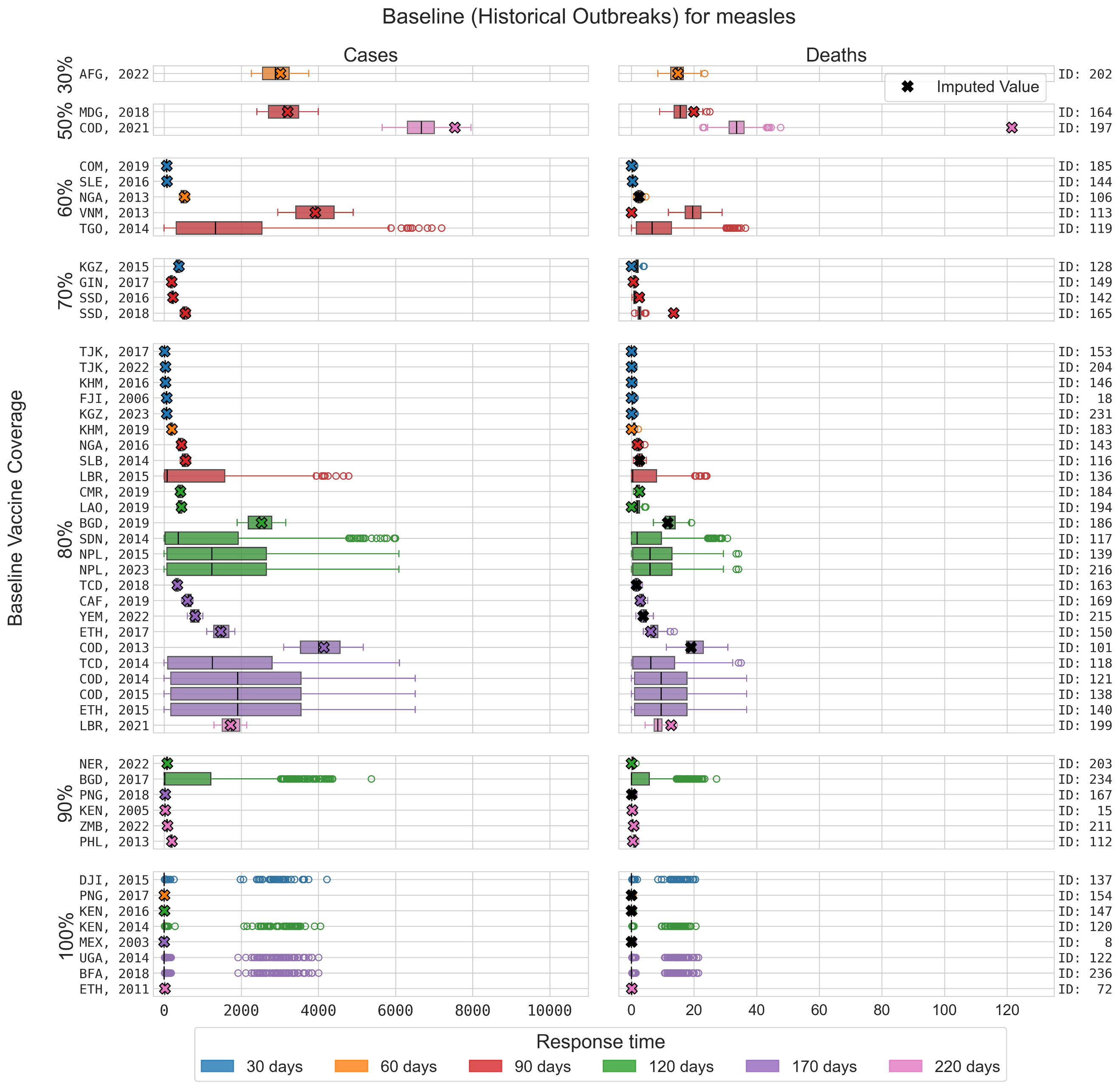

**Figure S7: Distribution of cases and deaths for each outbreak for model simulations retained after filtering.** Simulations which did not match within 25% of cumulative cases of historical outbreak were rejected. Crosses represent the data for each outbreak, scaled to a 50K population, and the colours represent the model-estimated baseline immunity.

##### *Baseline vs No ORI*

The distribution of simulated outcomes under the No ORI scenario is typically wider than the equivalent Baseline scenario and almost always larger (Figure S8).

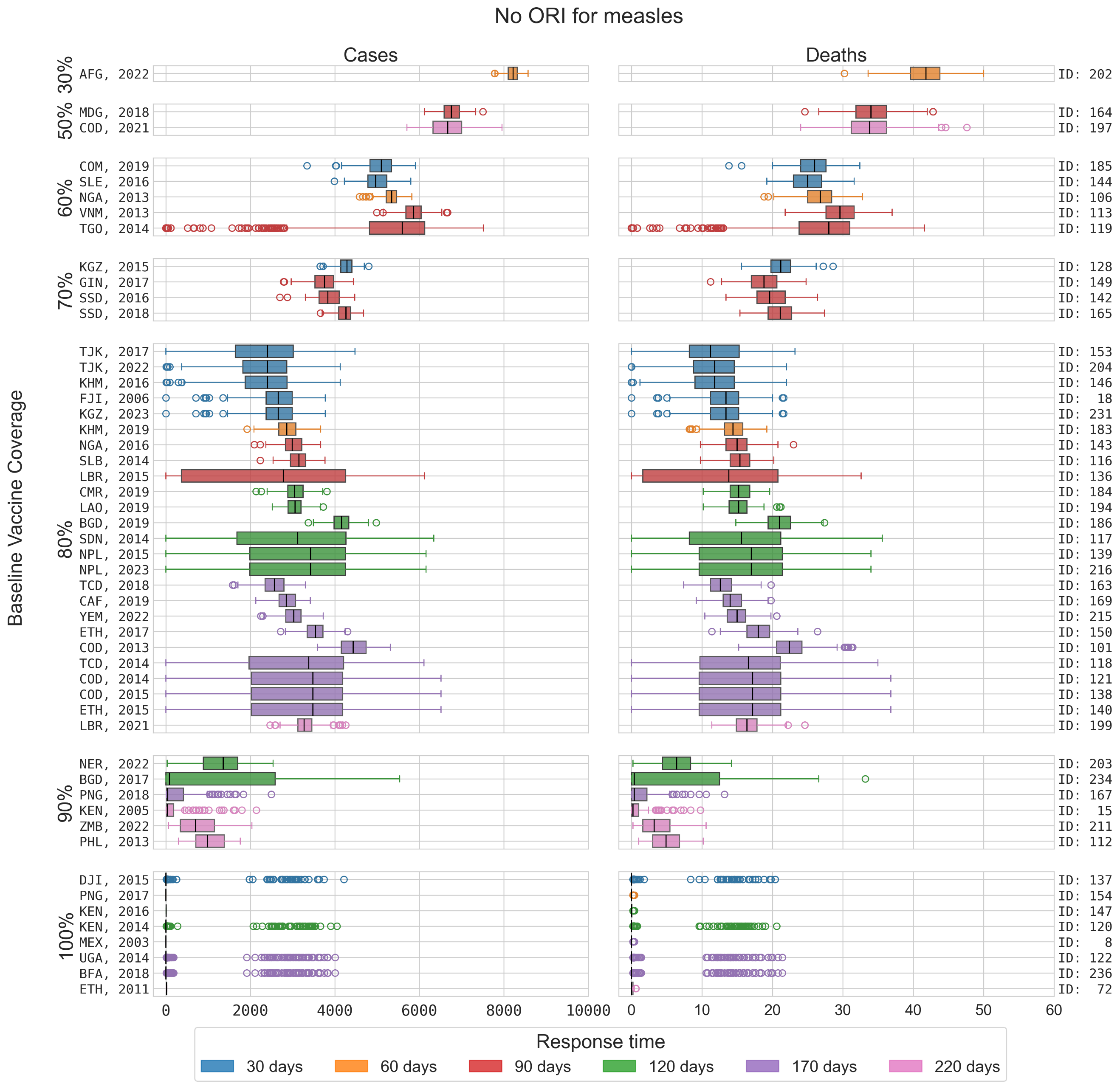

**Figure S8: Distribution of cases and deaths for each measles outbreak for counterfactual simulations with no ORI implemented.** Counterfactual simulations use equivalent transmission parameters and infection seeding to the matched Baseline simulations.

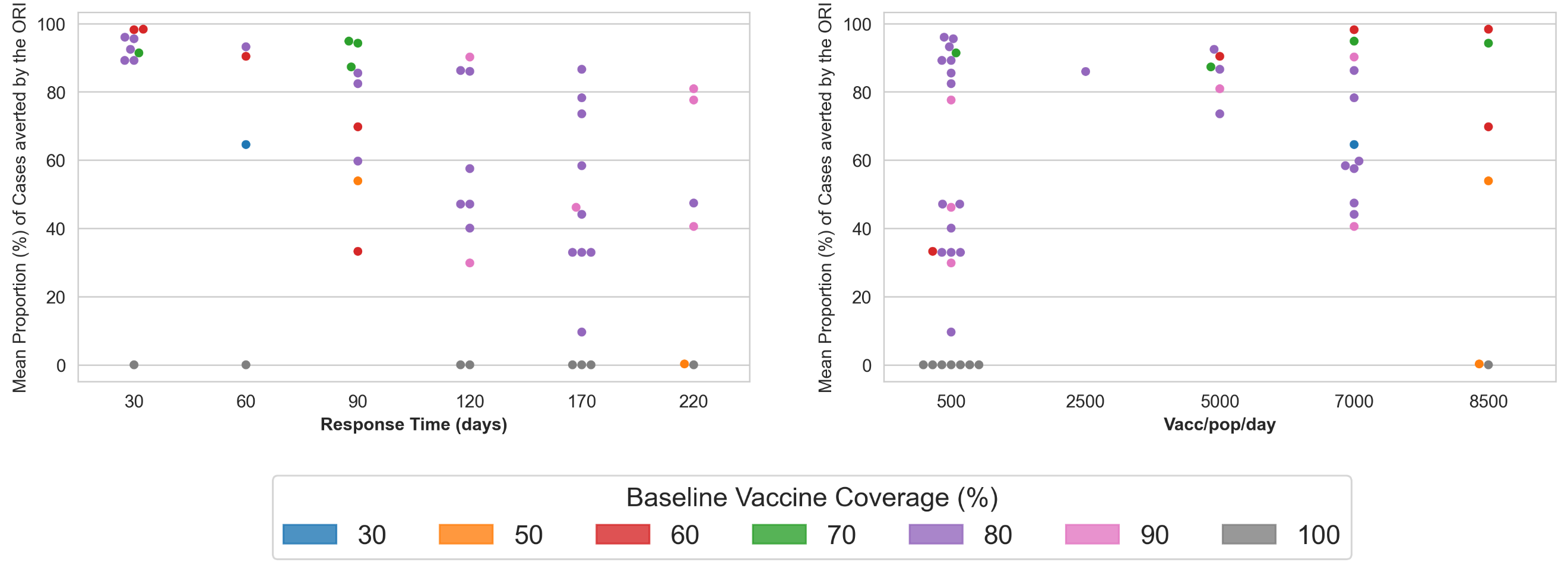

**Figure S9: ORI Impact estimation by lattice dimension**: Scatterplots showing the mean proportion of cases averted by the ORI, coloured by the assigned transmission level for each (fixed) lattice dimension.

**Table S3:** **Summary of impact estimation in 5-year increments.** Please note that 27% and 47% of outbreaks are missing the number of cumulative cases and deaths, respectively. Missing number and cumulative cases were estimated using the median model outcome and mortality rate, respectively. Estimated Cases/ Deaths/ DALYs/ societal costs averted are summarized as mean (95% uncertainty interval).

|  | **# outbreaks** | **Observed/ Estimated cases** | **Observed/ Estimated deaths** | **Cases averted** | **Deaths averted** | **DALYs averted** | **Costs averted** |
| --- | --- | --- | --- | --- | --- | --- | --- |
| 2001-2005 | 2 | 2588 | 27 | 20,495 (0 – 77,868) | 95 (0 – 358) | 5174 (0 – 19,504) | $1·40M ($0 – $5·29M) |
| 2006-2010 | 1 | 132 | 0 | 5138 (3026 – 7250) | 25 (13 – 37) | 1749 (932 – 2,567) | $2·68M ($1·44M – $3·93M) |
| 2011-2015 | 18 | 1·71M | 8143 | 1·45M (1·36M – 1·53M) | 7185 (6680 – 7691) | 466K (433K – 499K) | $345M ($323M – $371M) |
| 2016-2020 | 21 | 472k | 2457 | 2·11M (2·06M – 2·15M) | 10,564 (10,197 – 10,931) | 653K (631K – 676K) | $319M ($303M – $335M) |
| 2021+ | 9 | 619k | 8033 | 427K (396K – 457K) | 2133 (1958 – 2309) | 143K (131K – 155K) | $39·9M ($35·3M – $4·44M) |
| **Total** | 51 | 2·81M | 18,660 | 3·95M – 4·07M | 19·6K – 20·4K | 1.24M – 1.29M | $710M ($692M - 728M) |

ORI also reduced the risk of large outbreaks. There was a wide variation in outbreak size within the data, with 20% of outbreaks having fewer than 500 cumulative cases, 25% having 5000 – 100,000 cumulative cases, and 8% having more than 100,000 cases. However, with no ORI, the percentage of outbreaks with more than 100,000 cumulative cases increased from 8% to 41%.

#### **Key messages**

Of the 51 measles outbreaks over 2003-2023 where an ORI occurred and sufficient data was available for analysis, we estimate that the ORI averted a cumulative:

- 3·95M – 4·07M cases, compared to ~2·81M observed/estimated^[1]^
- 19,613 – 20,398 deaths compared to ~18,660 observed/estimated^1^
- 1·24M – 1·29M DALYs; and
- $692M - 728M societal economic costs

ORI was found to have higher impact in settings with lower baseline immunity, highlighting the importance of routine vaccination campaigns.

The presence of ORI was also estimated to have:

- Reduced the percentage of outbreaks with more than 100,000 cumulative cases from 41% to 8% (i.e. from twenty-one to four)
- Increased the percentage of outbreaks with fewer than 1000 cases from 16% to 29% (i.e. from eight to fifteen)

#### **Limitations**

- High uncertainty around case ascertainment: The model relies on 20% based on the ratio of cumulative cases and the target population size as observed in our data set. However, the literature reports a case ascertainment rate of 1-5% for measles^15^. If the case ascertainment rate is much lower than 20%, then the model may have underestimated cases, deaths, and vaccine impact.
- Uncertain case fatality rate: The model relies on 0·5% based on the observed mortality rate in our dataset, and the literature reports a mortality rate of 2·2% for measles^16^ differentiating between community-based and hospital-based settings. If the case fatality rate is greater than 0·5%, then the model may have underestimated deaths and vaccine impact.
- Uncertainty around vaccine efficacy: The efficacy value used in the model is from a study estimating the vaccine effectiveness in infants younger and older than 9 months but is for vaccine effectiveness rather than efficacy^9^. As such it does not directly translate to the model’s parameter and this potentially leads to an overestimate of its impact.
- Uncertainty around re-vaccination: Re-vaccination of a proportion of the target population is expected as prior determination of the vaccination status is challenging in LMICs. The model incorporates re-vaccination of agents; however, there is no change in immunity. The results might therefore underestimate the impact of the ORI.
- Modelling young children age group: The model considers young children age group. However, there could be important differences in transmission among older people, an indirect transmission from adults to children that are not captured.

### **Supplement 3: Cholera**

This supplementary section describes the context and more detailed methods for our cholera model and the ORI impact analysis.

#### **Background and motivation**

Cholera is a waterborne bacterial disease caused by the bacteria *Vibrio cholerae*, which can cause severe watery diarrhea and kill its host within hours if not treated^17, 18^. Infection occurs after the consumption of food or water contaminated by the bacteria, and rapid transmission can occur in populations without sources of safe drinking water or poor hygiene and sanitation conditions. Infectious people will shed bacteria, leading to further contamination of food and water, and freshly shed bacteria (within 5-18 hours) are highly infectious^19^, which can cause increased risk of transmission within households. A significant proportion of cholera infection is asymptomatic ^20, 21^, so people can spread the infection without knowing and this can make detection and accurate estimation of disease burden difficult. There are around 3 million cases and 100 thousand deaths due to cholera globally per year ^22^, with most of the burden occurring in sub-Saharan Africa.

Outbreaks of cholera are responded to with vaccines, when possible, but supplies of the vaccine are limited and differentiation of cholera from other causes of watery diarrhea can be challenging. Provision of fresh water and improved sanitation conditions are also effective tools for preventing cholera transmission, as they reduce the risk of exposure to the bacteria^23^. In order to estimate the historical impact of vaccines delivered in response to outbreaks of cholera we have developed a dynamic cholera transmission model which incorporates both Water, Sanitation, and Hygiene (WASH) interventions and vaccination.

#### **Model overview**

The *Starsim* framework was used to create an agent-based model of cholera among humans^1^, with states for susceptible, exposed, infected (symptomatically and asymptomatically), and recovered agents (Figure S10). The model also has a dynamic parameter for risk of environmental transmission to humans, which is calculated at each time step based on the relative concentration of the bacteria *Vibrio cholerae* in communal water sources (assuming that infectious humans shed the bacteria and increase the concentration).

Agents in the model represent humans, who begin as susceptible, and each day have a probability of becoming infected that depends on their immunity status and is proportional to the prevalence of infection among the population and the concentration of cholera bacteria in the environment. Following infection, humans enter a latent infection ‘exposed’ state, before becoming infectious to others; this infectious state can be either symptomatic or asymptomatic. Humans in either infectious state can recover and develop immunity to further infection during the modelled outbreak, and humans with symptomatic infection can die based on a disease-specific mortality rate. People can also be vaccinated, which will provide a level of immunity against future infection.

Transmission in the model can occur through two dynamic mechanisms. First, infected humans will shed bacteria during their infectious period, leading to contamination of food and water, increasing the prevalence of *Vibrio cholerae* in the environment and the risk of environmental transmission to all susceptible humans in the model. Second, people in the model are assigned a network of contacts (household and community; details in model population section), and there is a risk of human-human transmission between infected and susceptible contacts, which captures the increased infectiousness of freshly shed bacteria^19^. The prevalence of bacteria in the environment varies over time, increasing as infected humans shed fresh bacteria and decreasing as bacteria in the environment die. This prevalence is used to estimate a dynamic concentration of *Vibrio cholerae* in the environment via a dose-response relationship (parameterised as in Mukandevire et al.^24^.

The model assumes a baseline level of WASH interventions are present, which reduce the transmission risk from both human-human and environment-human transmission by improving hygiene and access to clean water, respectively. After an outbreak is detected and declared, the effectiveness of these WASH interventions is assumed to increase as people act to avoid infection.

The duration of vaccine immunity was set to well beyond the scope of the model period as it is assumed that no waning of immunity effects would be relevant over the outbreak period. Table S4 presents key epidemiological parameters used in the model and their justification or source.

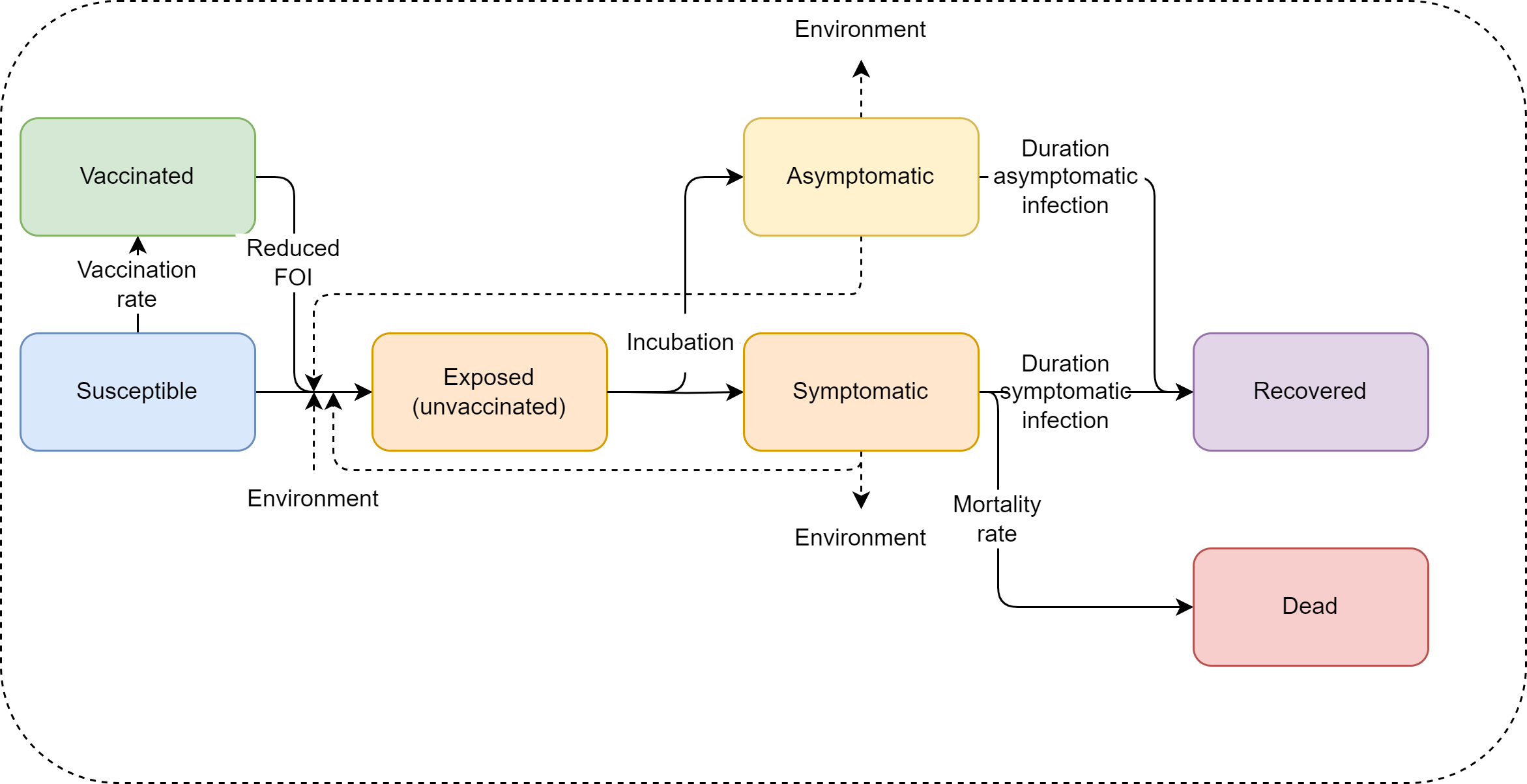

**Figure S10:** **Cholera model schematic**. The Starsim framework was used to develop an agent-based model of cholera among humans (S-E-I-R), which was paired with a dynamic prevalence of Vibrio cholerae in the environment to parametrize risk of environmental transmission.

**Table S4: Cholera model parameter values and sources.**

| Parameter | Value | Source / notes |
| --- | --- | --- |
| **Population parameters** |  |  |
| Population age distribution | Empirical distribution | United Nations, Department of Economic and Social Affairs, Population Division^2^; averaged over countries with outbreaks used in this analysis. |
| Household size distribution | Empirical distribution | United Nations, Department of Economic and Social Affairs, Population Division^2^; averaged over countries with outbreaks used in this analysis. |
| Mean ‘community’ contacts per day | 9 | Prem et al.^7^; averaged over all non-household contact rates and all ages, for all countries with outbreaks used in this analysis. |
| **Disease parameters** |  |  |
| Average duration of exposed period | 2·8 days | Model uses a lognormal distribution fit to results of systematic review by Azman et al.^25^ |
| Average duration of asymptomatic infection | 5·5 days | Model uses a uniform distribution of 1-10 days, based on WHO databook on cholera^26^. |
| Average duration of symptomatic infection (to recovery) | 5 days | Commonly used estimate in modelling literature, according to review by Fung^27^. Model uses a lognormal distribution fit to 5 day mean and range of 2-14 days. |
| Average duration of symptomatic infection (to death) | 1 day | According to Somboonwit et al.^18^, death can occur within 6-12 hours of symptoms for severe disease. Minimum timestep in the model is one day, and it uses a lognormal distribution with mean of 1 day and range of 1-3 days. |
| Duration of hyperinfectious bacteria | 1 day | New infected agents shed hyperinfectious bacteria for first day, with 50x lower threshold for infection. Based on approach by Hartley et al. and other modelling studies^19, 24, 28^. |
| Probability of death given symptomatic infection | 0·5% | Calibrated value, case fatality rate is < 1% for treated cholera^29^. |
| Probability of symptomatic cholera | 50% | Calibrated assumption from upper range of ~25% and 57% estimates from Jackson et al.^20^ Literature reports wide range of estimates for asymptomatic disease^21^. |
| Relative transmissibility from asymptomatic infection | 10% | Assumption, based on reduced bacteria shedding from asymptomatic cases reported by Nelson et al.^21^ |
| Vaccine protection against infection | One dose: 52·7% | Based on single dose effectiveness study by Malembaka et al.^30^ Maximum protection is reached 10 days after vaccination, based on 7-10 day estimate after two doses from Song et al.^31^ |
| **Health economics parameters** |  |  |
| Disability weights for cholera infection | Mild: 0·074  Moderate: 0·188  Severe:  0·247 | Global Burden of Disease (2017) Disability Weight estimates^3^. |
| Average life expectancy | Specific to country and year of outbreak | United Nations, Department of Economic and Social Affairs, Population Division^2^. Used to estimate years of life lost for each death. |
| Gross Domestic Product (GDP) per capita* | Specific to country and year of outbreak | World Bank, World Development Indicators^5^. |
| Discounting | 0% for DALYs; 3% for costs |  |

*Inflated to 2023 USD using average annual inflation rates since 2000^11^.

#### **Outbreak data**

Cholera outbreaks have been recorded by the WHO in its *Weekly Epidemiological Record* and *Disease Outbreak News* since 2000, with information on epidemiological and programmatic responses available online^12, 32, 33^.

Between 2000 and 2022 there were 335 recorded outbreaks, 81% of which were in Africa. 62 outbreaks were recorded as having received a vaccine response, and 40 outbreaks had sufficient response data for inclusion in the analysis. Data on cumulative cases, cumulative deaths, the time taken to start a response, and vaccines delivered per outbreak were the most complete, while information about the duration of the outbreak was the most incomplete (Table S5). The recorded outbreaks varied widely in scale: 1-2·6 million cumulative cases, 80 thousand-22·2 million vaccines delivered, and 7-274 days to respond.

**Table S5: Summary of outbreak data used for cholera analysis.**

   
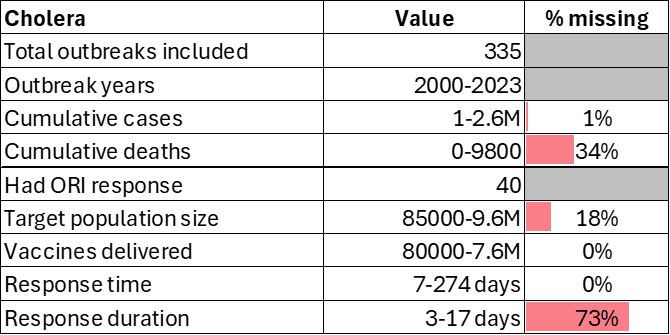

The following attributes were ascribed to each outbreak:

- Target population size: Directly from data, otherwise estimated based on number of vaccines (using linear relationship between number of vaccines and target population, fitted to all data with a target population and number of vaccines).
- Response time: Directly from data (time between outbreak declared and vaccine response started), which was then assigned to the closest of 30, 60, 90, or 130 days for simulation.
- Vaccine doses delivered: Directly from data where available, otherwise estimated based on target population size (using linear relationship between target population size and number of doses).
- Response duration: Directly from the data where available, otherwise based on average response duration from outbreaks in the same or nearby country.
- Response rate of vaccination: Directly from data where available, otherwise estimated based on target population size (using linear relationship between target population size and number of doses), which was then assigned to the closest of 1500, 2700, 6000, or 10,000 vaccines per day for simulation.

#### **Model population for simulated outbreaks**

The model population for each outbreak simulation represents the entire population of the specific geographic location where the outbreak occurred, based on the ‘target population size’ from the outbreak response data (i.e. people identified as being eligible for vaccination after the outbreak was declared). For computational reasons the model contains a maximum of 50,000 agents, and so for outbreaks with larger population sizes a scaling factor was used such that one agent in the model represents multiple people, based on the target population size and the proportion of the population eligible for the vaccine.

The model population was parametrized by age structure and household size distribution from the United Nations, Department of Economic and Social Affairs, Population Division^2^, and age-specific household contact rates from Prem et al.^7^. Each of these distributions and rates in the model were estimated using the average of the distributions and rates for each country with an outbreak included in the analysis. The age structure and household sizes are used to assign agents household contact networks, which are important for human-human transmission in the model. The model also randomly generates ‘community’ contacts between agents, which have a much lower risk of transmission compared to household contacts^34^, to capture potential transmission via freshly shed bacteria outside of an agent’s household. These networks are randomly generated at each time step (representing a day in the model). Vaccine coverage, transmission risk and disease outcomes were not modelled to vary by age given limited data from the outbreak settings

#### **Diagnosis of cases, outbreak declaration and ORI**

Symptomatically infected agents in the model have a daily probability of being detected as suspected cases of cholera (henceforth ‘cases’), assuming presentation at a health care facility or a diagnostic test being taken; the model does not distinguish between suspected and confirmed cases. An outbreak is declared in the model after the detection of a single symptomatic agent, and once this occurs the ORI will begin after N days, where N is the response time for a given outbreak.

#### **Calibration**

Calibration involves estimating the transmissibility of cholera in the model (human-human transmission risk per contact and proportionality constant for environment-human transmission), as well as the probability of symptomatic cholera and probability of death given symptoms. This was done by fitting to outbreaks in a separate dataset, maintained by Johns Hopkins University (with summary characteristics presented by Zheng et al.^35^), which includes disaggregated time series data for 1000 cholera outbreaks from 2010-2020. Many of these outbreaks were not in the WHO databases as they are informed by confidential surveillance reports or were likely too small to have been reported, and most of which did not have a vaccine response so could not be included in the main analysis. These data included: outbreak location, duration, threshold for declaration, total suspected cases, attack rate, total deaths, case fatality rate, reporting frequency, country population, mean R_0_, population density, rural/urban split, start date, end date, total confirmed cases, and the time to peak for each outbreak.

The time series data included daily or weekly case numbers for each outbreak, and an observed feature is a rapid initial growth followed by a slower phase. These two phases were used to constrain the two transmission mechanisms in the model; rapid initial transmission drive by human-human household contacts that is limited by the clustering of household contact networks, followed by slower but more widespread environment-human transmission, which must be high enough to take over but not so high as to produce rapid, widespread infection in most simulations. The human-human transmission risk per contact and proportionality constant for environment-human transmission in the model were adjusted to reproduce these features. The probability of symptomatic cholera and the probability of death given symptoms were adjusted to reproduce the average case fatality rate observed within the data, while fitting within the range of values described by the literature.

As Figure S11 indicates, most outbreaks are less than 1000 cases (after scaling to a model population of 50,000), but there is a small subset which grows to be much larger. In order to constrain cholera transmission in the absence of a vaccine response and produce (a) the rapid initial growth, (b) the slower secondary growth, and (c) a typically contained final size, the model contains WASH interventions to reduce transmission via both human-human and environment-human mechanisms (implemented as a relative reduction in the force of infection). These interventions are initialised with relatively low impact/coverage in the model, but after outbreak declaration the impact is strengthened as people in the model are assumed to prioritise sources of clean water and improve hygiene practices as much as possible during an outbreak. The timing and extent to which they increase following the detection of the outbreak was calibrated to reduce the rapid human-human transmission in households once cholera prevalence in the environment was high enough to drive the second phase of transmission. The WASH interventions were also calibrated to limit the environmental force of infection during the second phase of transmission, such that very large outbreaks were much less frequent than smaller outbreaks.

Figure S11 shows how the model captures the growth rate and range of outbreak sizes without a vaccine response, compared with the Johns Hopkins University data set.

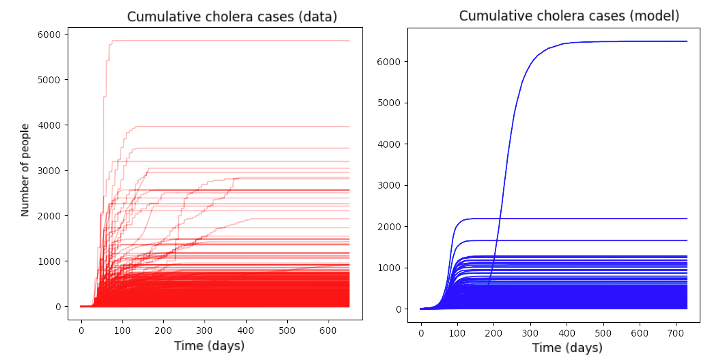

**Figure S11: Comparison of Johns Hopkins University cholera outbreak time series (scaled to a population of 50K) against an example set of model time series (using a population of 50K agents) after calibration.** The model is able to reproduce the rapid initial epidemic growth and produces typically small outbreaks with some infrequent larger outbreaks, as is observed in the data.

Once the transmission, mortality, and symptomatic parameters had been calibrated, outbreaks were simulated by infecting three humans in the model at random, for a range of parameter lattice points representing vaccine response times (30, 60, 90, 130 days) and vaccination rates (1500, 2700, 6000, 10,000 vaccines per day). The set of 40 outbreaks with a vaccine response and sufficient data were assigned to their nearest lattice point based on the observed response time and vaccination rate and compared to the distribution of model outbreak simulations (Figure S12). It can be seen that the parameters which were calibrated to the Johns Hopkins University dataset also produce outcomes which align well with the 40 outbreaks we considered, as all case counts fall within the range of cases produced by the model. For each lattice point, a variable number of simulations were run, depending on the number of outbreaks assigned to the lattice point and how frequently the model would simulate outbreaks of a similar size.

 
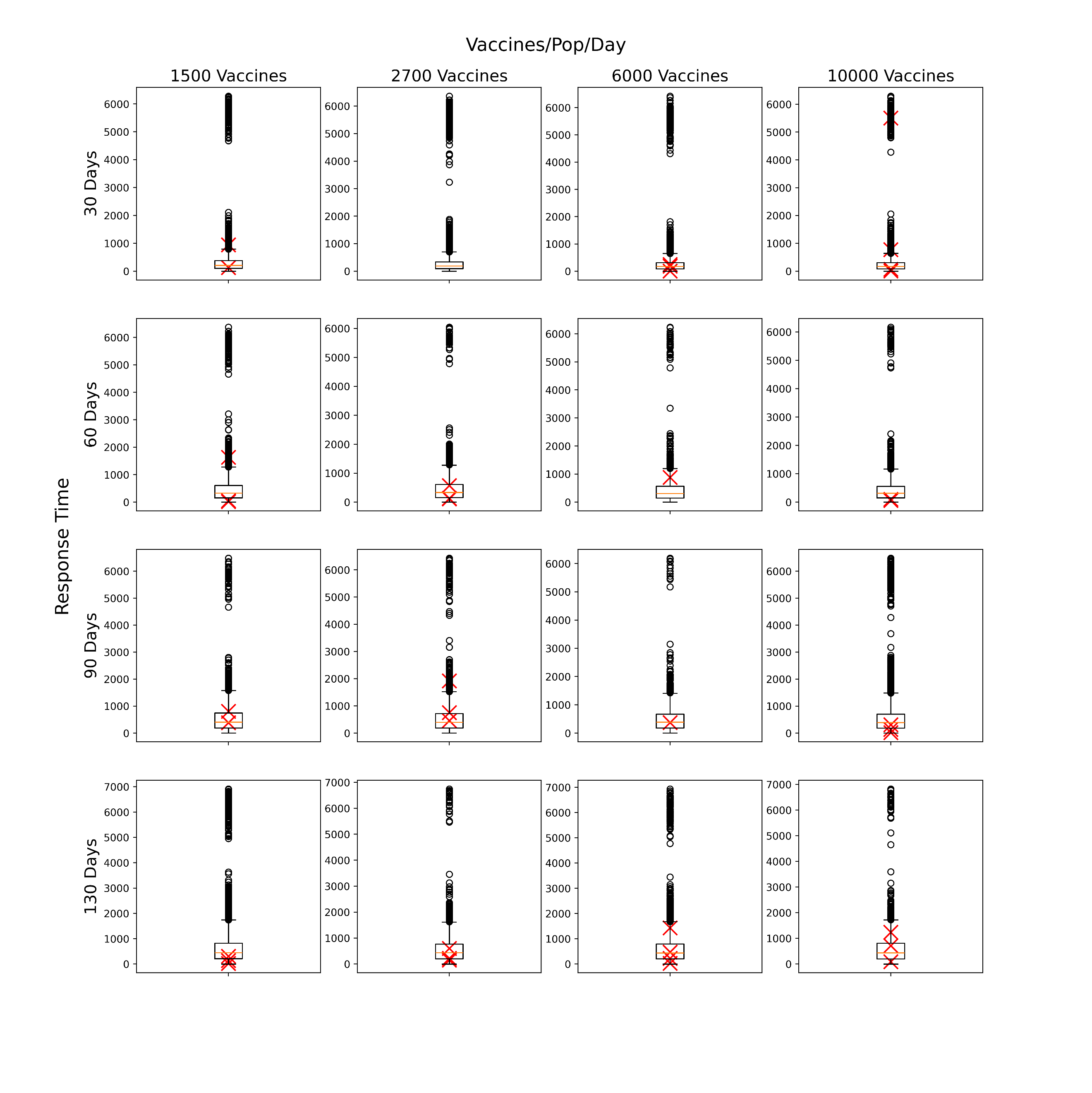

**Figure S12: Lattice of cholera simulations before outbreak filtering was applied.** Each boxplot represents the distribution of total cases from simulations run with response parameters defined by the row/column lattice point. The red crosses represent the cumulative suspected cases from observed outbreaks which have been assigned to the lattice point and scaled to a population of 50K.

#### **Scenarios**

For the purpose of running counterfactuals specific to the historical outbreaks, for each outbreak in the data, the set of simulations associated with the lattice point that the outbreak was assigned to were filtered to retain only those simulations where the cumulative cases were within +/- 25% of the reported cases (see Trajectory selection). Two scenarios were then examined:

- Baseline: ORI as occurred, using the filtered simulations from the lattice
- No ORI counterfactual: Simulations were run without any ORI intervention using the same filtered simulation seeds from the baseline scenario, such that everything was identical up until the date the ORI would have started.

#### **Outcomes**

For each outbreak and scenario, the distribution of cumulative cases and deaths across selected model simulations were recorded. The mean difference in outbreak size between scenarios and associated uncertainty in the mean were estimated from these collections of simulations using bootstrap resampling (i.e., for each outbreak to produce estimates of cases averted by ORI).

Total cases averted by ORI across all historic outbreaks was then estimated by aggregating the cases averted for each individual outbreak. As the outbreaks are independent, this was obtained by summing the cases averted per outbreak, with the variance estimated by summing the variances from each individual outbreak.

DALYs averted by ORI were estimated by multiplying cases averted by the disability weight per case and average duration of symptoms, plus years of life lost from deaths (for each outbreak, the average life expectancy in that country and year compared to the age of deaths in the model). Socio-economic costs averted by ORI were calculated by estimating productive years of life lost or lived with disability, and multiplying this by GDP per capita. Productive years of life lost were estimated as the difference between the average age of death in the model and a retirement age of 64, or the life expectancy in that country and year, whichever was lower. Costs were inflated to 2023 USD, with future costs discounted at 3% per annum as a standard method.

The impact of ORI on reducing the risk of large outbreaks was also estimated, by comparing the distribution of cumulative cases across outbreaks in the data to the distribution of cumulative cases across outbreaks in the no ORI scenario. For the purposes of this sub-analysis, cumulative cases in the no ORI scenario for each outbreak were approximated as the median from the no ORI counterfactual simulations.

#### **Results**

##### *Baseline*

The modelled cumulative cases and deaths for the baseline scenario is shown for each outbreak considered in the analysis in Figure S13. Outbreaks are coloured by the categorised response time or the ORI, and data points are scaled to the vaccine-eligible population in the model. The fit of the simulations to the case data presented in Figure S13 differs from what is presented in Figure S12 (i.e., the median of the model simulations sits close to the data) as we are considering only the filtered simulations which align well with the observed cases. It should be noted that as the model applies a uniform probability of death given symptomatic disease (estimated from the average case fatality rate observed during calibration), if the fatality rate for a given outbreak differs then the model is unable to capture the cumulative deaths as well as the cumulative cases.

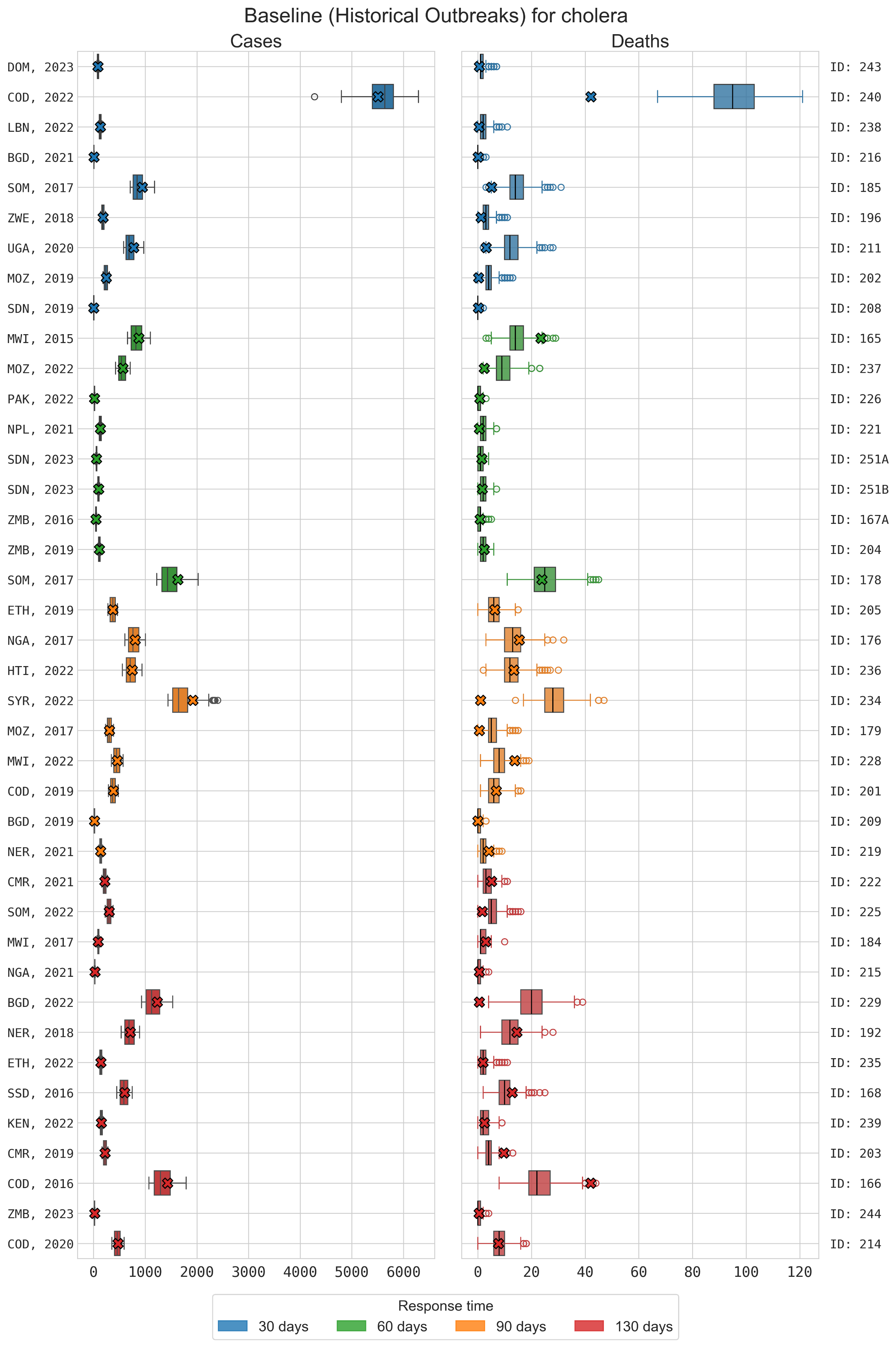

**Figure S13: Distribution of cases and deaths for each cholera outbreak for model simulations retained after filtering.** Simulations which did not match within 25% of cumulative cases of historical outbreak were rejected. Crosses represent the data for each outbreak, scaled to a 50K population, and the colours represent the response time used for each outbreak.

##### *Baseline vs. No ORI*

The distribution of simulated outcomes under the No ORI scenario are typically wider than the equivalent Baseline scenario, as they are not constrained by filtering against observed case data, and almost always larger (Figure S14).

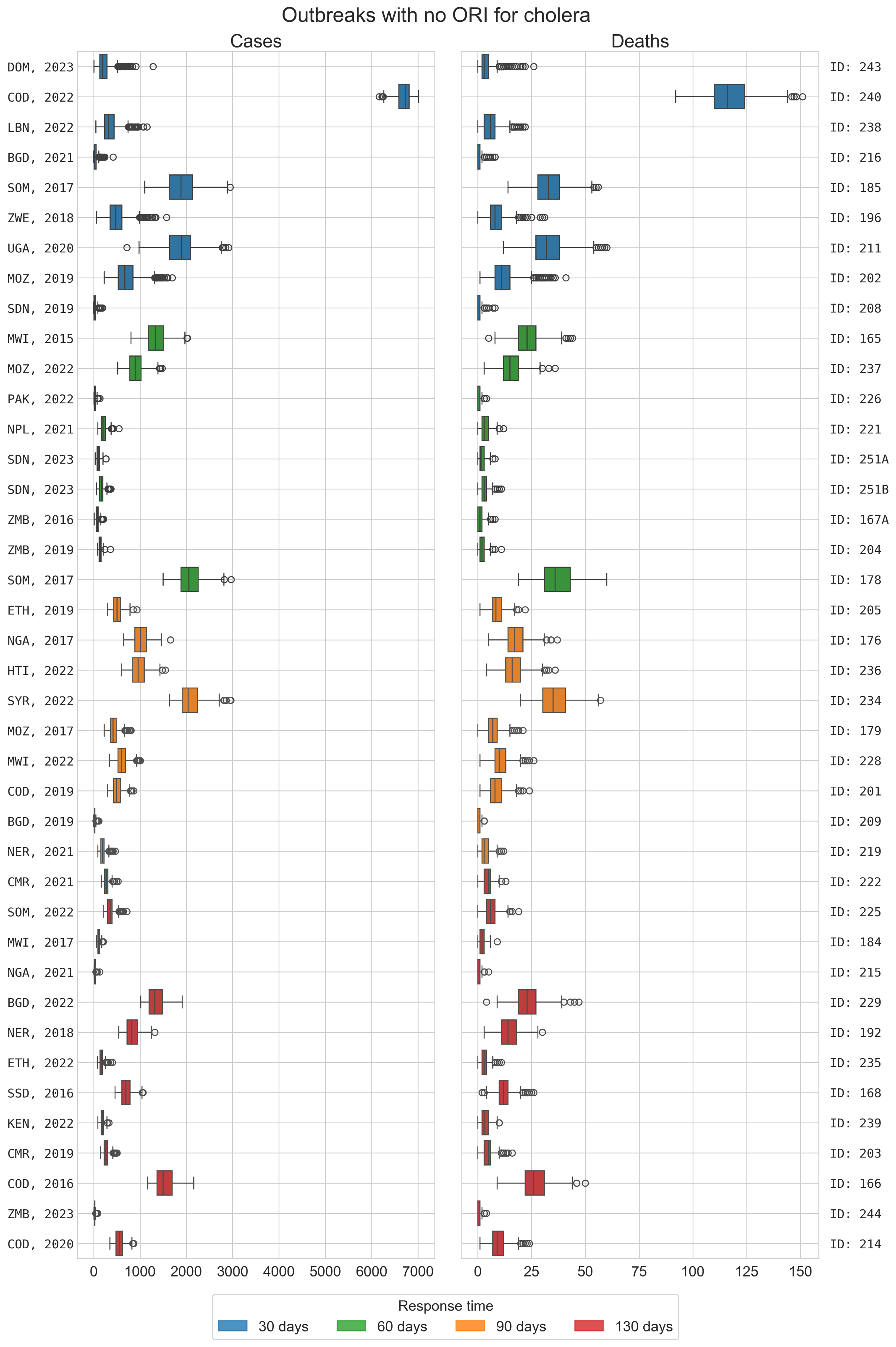

**Figure S14: Distribution of cases and deaths for each cholera outbreak for counterfactual simulations with no ORI implemented.** Counterfactual simulations use equivalent transmission parameters and infection seeding to the matched baseline simulations.

**Table S6: Summary of estimated ORI impacts across cholera outbreaks in five-year periods.** Estimated Cases/Deaths/DALYs/societal costs averted are summarized as the mean and 95% uncertainty intervals.

| Years | # outbreaks | Observed cases | Observed deaths | Cases averted | Deaths averted | DALYs averted | Costs averted (2023 USD) |
| --- | --- | --- | --- | --- | --- | --- | --- |
| 2011-2015 | 1 | 1795 | 48 | 1053 (470 – 1637) | 18 (-3 - 39) | 723 (-154 – 1602) | 275K (-52K – 601K) |
| 2016-2020 | 19 | 283,844 | 4721 | 136K (128K – 144K) | 2464 (2190 – 2738) | 101K (89K – 112K) | 66·6M (56·4M – 76·7M) |
| 2021-2023 | 20 | 514,868 | 4490 | 145K (137K – 154K) | 2728 (2357 – 3099) | 119K (102K – 136K) | 88·5M (74·5M – 102·4M) |
| Total | 40 | 800,057 | 9259 | 283K (273K – 292K) | 5215 (4879 – 5551) | 220K (205K – 236K) | $156M ($145M – 166M) |

ORI also reduced the risk of large outbreaks. There was a wide variation in outbreak size within the data, with 45% of outbreaks having fewer than 5000 cumulative cases, 25% having 20,000 – 50,000 cumulative cases, and 10% having more than 50,000 cases. However, with no ORI, the percentage of outbreaks with more than 50,000 cumulative cases increased from 10% to 17·5%.

#### **Key messages**

The vaccine responses delivered to 40 cholera outbreaks from 2015-2023 averted significant burdens of disease, with more than half of the estimated impact being accrued since 2021. Notably, responses are estimated to have been more effective when response times were faster.

We estimate that the ORI averted a cumulative:

- 273K – 292K cases, compared to 800K observed
- 4879 – 5551 deaths compared to 9259 observed
- 205K – 236K DALYs; and
- $145M – $166M societal economic costs

The presence of ORI was also estimated to have:

- Reduced the percentage of outbreaks with more than 50,000 cumulative cases from 17·5% to 10% (i.e. from seven to four)
- Increased the percentage of outbreaks with fewer than 1000 cases from 12·5% to 17·5% (i.e. from five to seven)

#### **Limitations**

- Actual transmission might be heterogeneous and could be a factor that limits outbreak size: there are multiple sources of heterogeneity that the model cannot capture, across the populations, transmission networks, and environmental reservoirs for cholera. This includes:
  - Age-based contact networks beyond the household
  - Other highly connected contact networks (e.g., workplaces, schools, social groups)
  - Geographical heterogeneities in where people interact with contaminated water
  - Coverage and efficacy of WASH interventions
- High uncertainty around vaccine efficacy: only a small number of studies have investigated the efficacy or effectiveness of a single dose of oral cholera vaccines, with highly variable reported values. The efficacy value used in the model is from the most recent study with a large sample size and robust methods, but is for vaccine effectiveness rather than efficacy. As such it does not directly translate to the model’s parameter and this potentially leads to an overestimate of its impact. There was only a single study found which investigated single dose efficacy, and multiple experts suggested that the value was too low during validation of the model, so we aligned with the effectiveness value.
- Reporting of cholera cases: the model is calibrated to cumulative suspected cases reported for each outbreak:
  - This may be inflated by cases of acute watery diarrhea which are not due to cholera. If that is the case, then the model may overestimate the transmissibility of cholera which would produce overestimates of ORI impact.
  - On the other hand, reported cases are almost certainly an underestimate of true symptomatic infections due to underreporting, in which case the model may underestimate the transmissibility of cholera which would produce underestimates of ORI impact. We attempt to account for this with a calibrated case ascertainment rate of 20-25%, however there is little data to validate this against and it is likely highly variable across outbreaks.
- Outbreak detection thresholds using a scaled model population: the model population of 50,000 is used to represent outbreaks which occurred in much larger settings, and hence a single case in the model can represent dozens of cases. This means that a threshold of one case for outbreak declaration in the model does not truly represent only a single case, which may be effectively delaying outbreak detection in the model compared to reality.
- Uncertain case fatality rate: The model relies on 0·5% based on the observed mortality rate in our dataset, and the literature reports a mortality rate of <1% for treated cholera^29^. However, recent cholera outbreaks have had significantly higher fatality rates^36^. If the case fatality rate is greater than 0·5%, then the model may have underestimated deaths and vaccine impact.

**Supplement 4: Yellow fever**

This supplementary section describes the context and more detailed methods for our yellow fever model and the ORI impact analysis.

#### **Background and motivation**

Yellow Fever is an epidemic-prone mosquito-borne vaccine preventable disease that is transmitted to humans by the bites of infected mosquitoes, mostly from bites occurring during the day^37^. WHO considers it a high-impact high-threat disease that has the potential to spread internationally^37^. Yellow Fever outbreaks have been recorded in 34 countries in Africa and 13 countries in Central and South America and are classified as either domestic (around houses), sylvatic (in forests or jungles) or semi-domestic outbreaks (both habitats)^37^. Outbreaks can also be differentiated by the transmission cycle, with ‘spillover’ outbreaks involving non-human primates and *Aedes africanus, Haemagogus spp.* and *Sabethes spp.* mosquitos, and ‘human-to-human’ outbreaks with transmission mediated by *Aedes aegypti* mosquitos^38^. The latter transmission cycle is typically associated with large, high-risk outbreaks and we therefore focus on outbreaks involving only humans in this study.

In countries where yellow fever occurs, the WHO strongly recommends routine vaccination for everyone older than 9 months, and prevention of outbreaks in affected regions requires at least 80% vaccine coverage of the population at-risk^39^.

#### **Model Overview**

The *Starsim* framework was used to create an agent-based model of yellow fever among humans^1^, which was coupled to a compartmental susceptible-exposed-infectious-susceptible (S-E-I-S) model among mosquitos so that risk of environmental transmission to humans could be approximated by a dynamic parameter for prevalence among mosquitoes (represented in Figure S15).

Agents in the model represent humans, who begin as susceptible or vaccinated, and each day have a probability of becoming infected that is proportional to the prevalence of infection among the mosquito population, and that further depends on their vaccination status. Following infection, humans enter a latent infection ‘exposed’ state, before developing severe or non-severe disease and becoming infectious to mosquitoes. Humans in either of these infectious states can recover and develop immunity, and humans in the severe infectious state can die based on a disease-specific mortality rate. We did not disaggregate asymptomatic infections because little data is available to inform differences in disease duration and infectiousness, and because testing behaviour is mainly driven by severe vs non-severe disease.

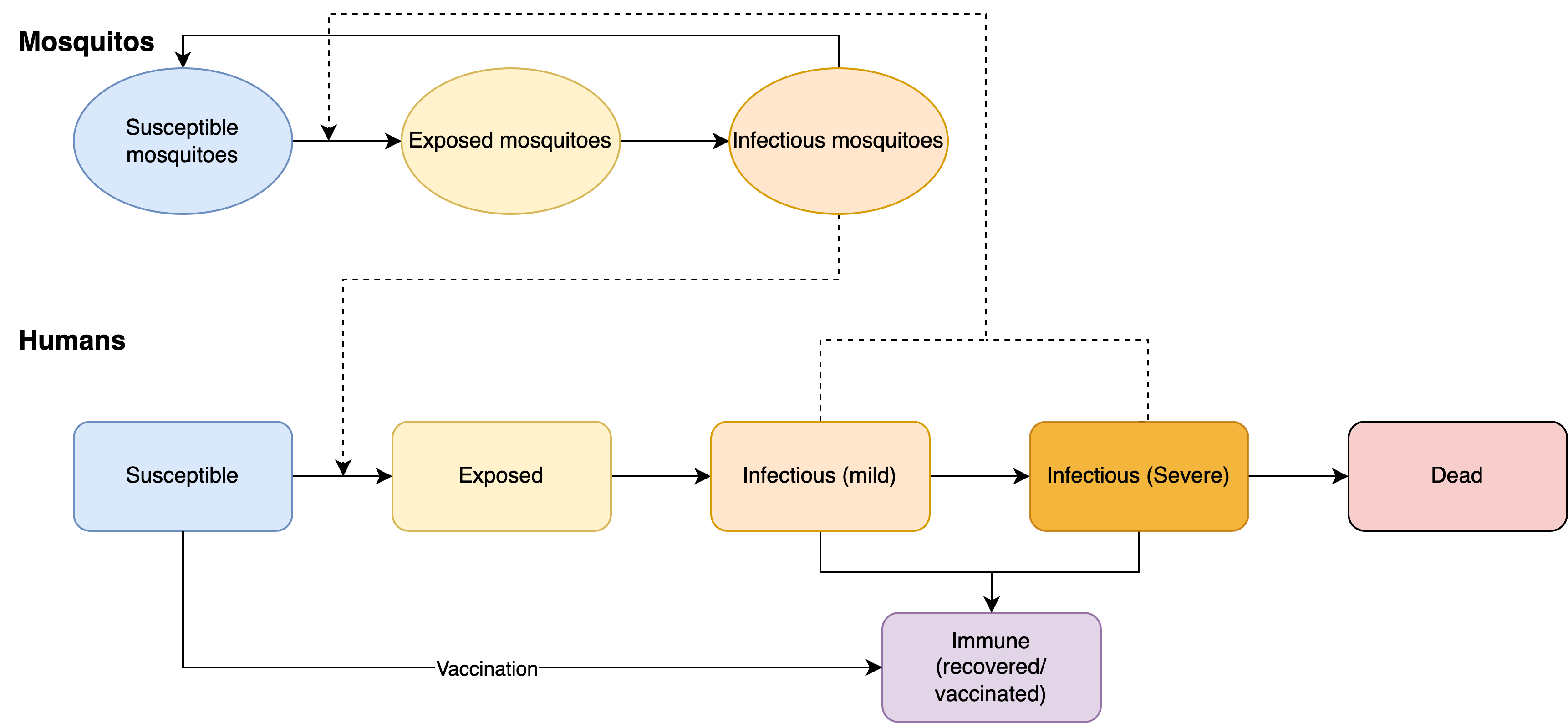

**Figure S15: Yellow fever model schematic**. The Starsim framework was used to develop an agent-based model of yellow fever among humans (S-E-I-R), which was paired with a dynamic compartmental model of infection among mosquitoes to parametrize risk of environmental transmission.

**Table S7: Yellow fever model parameters and sources.**

| Parameter | Value | Source / notes |
| --- | --- | --- |
| **Population parameters** | | |
| Population age distribution | Empirical distribution | United Nations, Department of Economic and Social Affairs, Population Division^2^; averaged over countries with outbreaks used in this analysis. |
| **Disease parameters** | | |
| Probability of death, given severe disease | 20% | Calibrated value. |
| Probability of mild/asymptomatic infection | 33% + 55% (mild + asymptomatic) | Based on probability of yellow fever virus infections being asymptomatic, mild or severe^40^. |
| Probability of developing severe disease | 12% |  |
| Average duration of exposed period | 3-6 days | Model uses a uniform distribution of 3-6 days, based on WHO data book on yellow fever^37^. |
| Average duration of mild infection (to recover) | 3-4 days | Initial symptoms usually disappear after 3-4 days. Model uses a uniform distribution of 3-4 days, based on WHO data book on yellow fever^37^. |
| Average time to develop severe disease | 4-5 days | Within 24hrs of recovering from initial symptoms, a small percentage of patients enter a second, more toxic phase. Therefore, the model uses a uniform distribution of 4-5 days (duration of mild disease 3-4 days + 24hrs), based on WHO data book on yellow fever^37^. |
| Average duration of severe infection (to recovery/death) | 7-10 days | Proportion of patients who enter the toxic phase die within 7-10 days. Model uses a uniform distribution of 7-10 days, based on WHO data book on yellow fever^37^. |
| Vaccine protection against infection | 97·5% | Based on random-effects meta-analysis and pooled estimate of serological response. Vaccine protection increases linearly over time; it reaches 80% after 10 days and 97·5% after 30 days. Time courses from WHO^37^. Peak vaccine protection from Jean et al.^41^ |
| **Health economic parameters** | | |
| Disability weights for yellow fever infection | Severe: 0·133 | Global Burden of Disease (2017) Disability Weight estimates^3^. |
| Average life expectancy | Specific to country and year of outbreak | United Nations, Department of Economic and Social Affairs, Population Division^2^. Used to estimate years of life lost for each death. |
| Gross Domestic Product (GDP) per capita* | Specific to country and year of outbreak | World Bank, World Development Indicators^5^. |
| Discounting | 0% for DALYs; 3% for costs |  |

*Inflated to 2023 USD using average annual inflation rates since 2000^11^.

#### **Outbreak data**

Yellow fever outbreaks have been recorded in WHO reports since 2000, with information on epidemiological and programmatic responses available online^12^. Data on yellow fever outbreaks was extracted from WHO reports, the literature and the Gaythorpe et al. 2021 Git Repository^42^.

Between 2000 and 2023 there were 215 recorded outbreaks. Out of these, 112 outbreaks had data on a response. However, 24 outbreaks were excluded from the analysis due to insufficient response data or unreasonably long response times. A total of 88 outbreaks were included in the impact estimation, 95% of which were in Africa and 5% in Central or South America. Data on cumulative cases and vaccines delivered per outbreak were the most complete, while information about the ORI response time and duration of the outbreak were the most incomplete (Table S8). The recorded outbreaks varied widely in scale (1-9000 cumulative cases), vaccines delivered (35,000-22·2M) and time to respond (6-330 days).

**Table S8: Summary characteristics of outbreak data used for yellow fever analysis.**

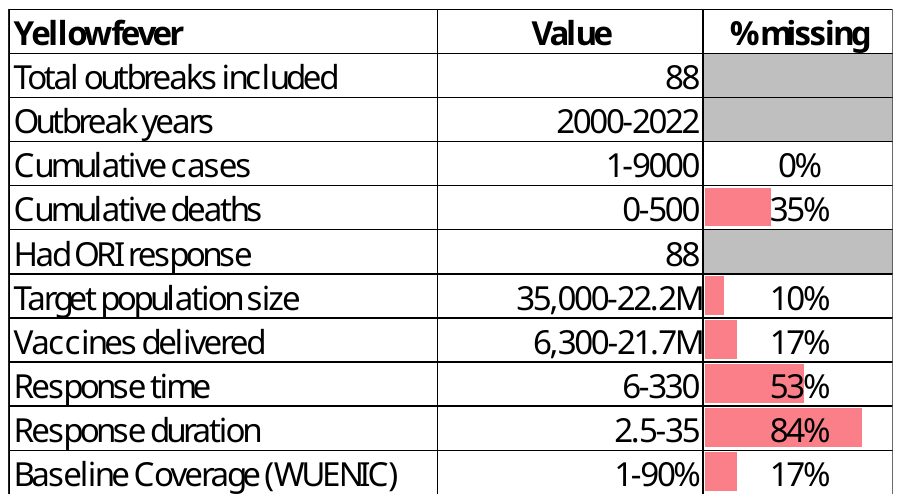

Some model parameter values were set based on the following quantities from the outbreak dataset, and imputed where necessary (see Imputing missing variables in outbreak data):

- Baseline vaccine coverage: If WUENIC data was available for the years prior to the outbreak, the average coverage across years prior to (and excluding) the outbreak year. If there was no WUENIC data for the years prior the outbreak, estimated as the average across all available years.
- Target population size: Directly from data, otherwise estimated based on number of vaccines (using linear relationship between number of vaccines and target population, fitted to all data with a target population and # of vaccines).
- Response time: Directly from data (time between outbreak declared and vaccine response started) if available, otherwise the average response time from outbreaks in the same or nearby country.
- Vaccine doses delivered: Directly from data where available, otherwise estimated based on target population size (using linear relationship between target population size and number of doses).
- Response duration: Directly from the data where available, otherwise based on average response duration from outbreaks in the same or nearby country.
- Response rate of vaccination: Calculated based on number of doses and response duration.

The lattice of parameter values for which the model was run was obtained by binning these variables, as well as the transmission parameter beta, into the following levels:

- Baseline vaccine coverage: 40%, 50%, 60%, 70%, 80%
- Response time: 30, 60, 90, 120, 170, 220, 300 days
- Vaccination rate: 500, 2500, 5000, 7500, 10000, 12500 vaccines per 50,000 population per day
- Transmission setting: assigned through calibration process described below, with classifications of low, medium low, medium, medium high, high, implemented as factor multipliers (0·5, 0·75, 1, 1·25, and 1·5) on the calibrated baseline beta value, to modify mosquito-to-human transmission.

#### **Model population for simulated outbreaks**

The model population for each outbreak simulation represents the entire population of the specific geographic location where the outbreak occurred, based on the ‘target population size’ from the outbreak response data (i.e., people identified as being eligible for vaccination after the outbreak was declared). For computational reasons the model contains a maximum of 50,000 agents, and so for outbreaks with larger population sizes a scaling factor was used such that one agent in the model represents multiple people, based on the target population size and the proportion of the population eligible for the vaccine. For this model, the proportion of the population eligible for the vaccine corresponds to ~97·8% excluding children younger than 9 months.

The model population for each outbreak was parametrized by age structure from the United Nations, Department of Economic and Social Affairs, Population Division^2^, which provides an age distribution in one-year increments. The baseline vaccination coverage was estimated from the WHO/UNICEF Estimates of National Immunization Coverage (WUENIC)^43^. Agents in the model are assigned integer ages, so to capture the vaccine eligibility of children >9 months^37^, 25% of <1-year olds were assumed eligible (i.e. assuming a uniform distribution of age within this group). Aside from vaccine eligibility for children <9 months old, vaccine coverage, transmission risk and disease outcomes were not modelled to vary by age given limited data from the outbreak settings. Transmission in the model occurred through interactions with the mosquito population (e.g. per day probability of infection that depended on prevalence) rather than direct human-human contact, hence no contact network structure among agents in the model was required.

#### **Diagnosis of cases, outbreak declaration and ORI**

An estimated 12% of infections become severe (Table S7), and so assuming only severe cases get reported a 12% case ascertainment rate was used^38^. The smallest outbreak in the data set recorded one case, i.e. one severe case, indicating that outbreaks are declared with just one diagnosis. WHO also states that outbreaks are declared in the ‘presence of at least one confirmed case of yellow fever’ ^44^. Therefore, an outbreak was considered declared as soon as one agent progressed to the severe state, and all severe cases were assumed to be reported. The ORI then begins after N days, where N is the response time for a given outbreak.

As we assume a high diagnosis rate for severe infections and negligible diagnosis for non-severe infections, in the following analysis ‘cases’ is synonymous with the number of severe infections.

#### **Calibration**

Calibration involves estimating the proportionality constants beta linking transmission from mosquitoes to humans and from humans to mosquitoes. The transmission from humans to mosquitoes affects the amount of change in the mosquito prevalence, which gets updated at each time step (representing a day) in the simulation using the human prevalence. The transmission from mosquitoes to humans and the mosquito prevalence control the probability of a susceptible agent becoming infected. The calibration of the proportionality constants was done by using known estimates of the reproduction number R_0_ (provided by Fraser et al.^45^) and the final size equation:

$$\pi=1- e^{-R_{0}*\pi}$$

The final size equation links R_0_ to the expected cumulative number of infections when an outbreak is simulated in a naïve population in an SEIR model. In a naïve population, the transmission between mosquitoes to humans and humans to mosquitoes is likely to be very similar and so assumed to be equivalent. Using the maximum estimated R_0_ of 1·45, we expect that ~55% of a naïve population gets infected in the absence of a response. In the model, the baseline transmission probability was therefore calibrated such that this occurred when an outbreak was simulated without any prior immunity or any response.

However, the baseline R_0_ value alone does not capture variability in transmission levels for different settings, with differences likely arising from climate and other factors that influence heterogeneity in transmission across settings. To account for this, an additional calibration variable was introduced to classify settings as low, medium low, medium, medium high, high transmission risks, implemented as factor multipliers on the calibrated baseline beta value, to modify mosquito-to-human transmission.

To produce the lattice of baseline simulations (Figure S16), the model was initialized with a value for baseline vaccine coverage (40%, 50%, 60%, 70% or 80%), response time (30, 60, 90, 120, 170, 220 or 300 days), vaccination rate (500, 2500, 5000, 7500, 10,000, or 12,500 vaccines per 50,000 population per day) and transmission level (low, medium low, medium, medium high or high). Stochastic outbreak simulations were produced by infecting five agents (which was sufficient to produce an outbreak) in the model at random at the start of the simulation, and then running the model 2000 times.

Finally, observed outbreaks (i.e. historical outbreaks with a given baseline vaccine immunity, response time and vaccination rate) were then assigned a transmission level based on a maximum-likelihood estimate of the total number of cumulative cases in the outbreak (see Kernel Density Estimation) (Figure S16; boxplots representing the range of simulated outcomes and crosses representing the observed outbreaks where they have occurred).

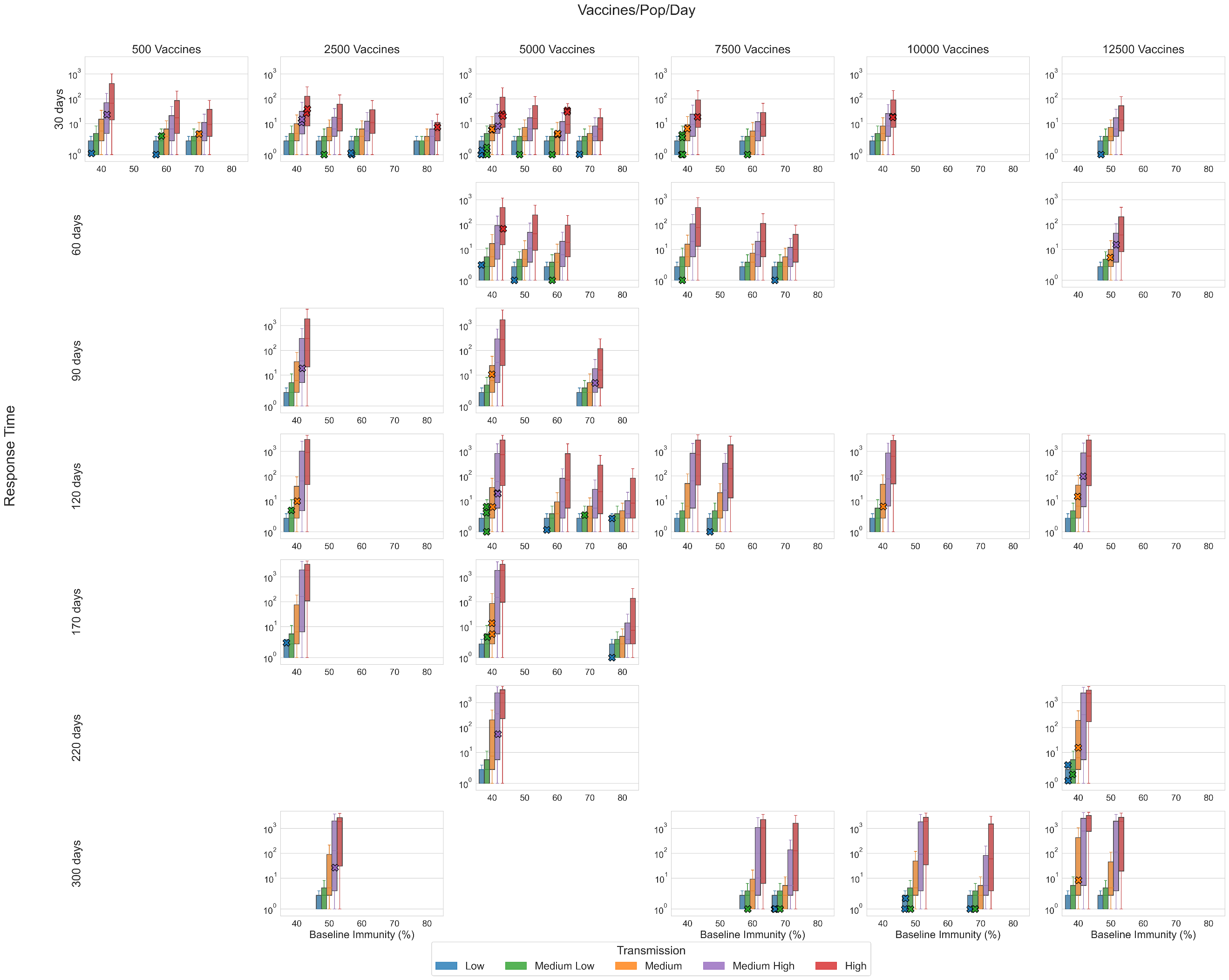

**Figure S16: Lattice of yellow fever simulations before outbreak filtering was applied.** Each boxplot represents the distribution of total cases from a simulation run with response parameters defined by the lattice point. The crosses represent the cumulative suspected cases observed from an outbreak which has been assigned to the lattice point and scaled to a population of 50K. Cumulative cases below 1 have been rounded up to 1 for plotting purposes.

#### **Scenarios**

For the purpose of running counterfactuals specific to the historical outbreaks, for each outbreak in the data, the 2000 simulations associated with the lattice point that the outbreak was assigned to were filtered to retain only those simulations where the cumulative cases were within +/- 25% of the reported cases (see Trajectory selection). Two scenarios were then examined:

- Baseline: ORI as occurred, using the filtered simulations from the lattice
- No ORI counterfactual: Simulations were run without any ORI intervention using the same filtered simulation seeds from the baseline scenario, such that everything was identical up until the date the ORI would have started.

#### **Outcomes**

For each outbreak and scenario, the distributions of cumulative cases and deaths across selected model simulations were recorded. The mean difference in outbreak size between scenarios and associated uncertainty in the mean were estimated from these collections of simulations using bootstrap resampling (i.e. for each outbreak to produce estimates of cases averted by ORI).

The total cases averted by ORI across all historic outbreaks was then estimated by aggregating the cases averted for each individual outbreak. As the outbreaks are independent, this was obtained by summing the cases averted per outbreak, with the variance estimated by summing the variances from each individual outbreak.

DALYs averted by ORI were estimated by multiplying cases averted by the disability weight per case and average duration of symptoms, plus years of life lost from deaths (for each outbreak, the average life expectancy in that country and year compared to the age of deaths in the model). Socio-economic costs averted by ORI were calculated by estimating productive years of life lost or lived with disability, and multiplying this by GDP per capita. Productive years of life lost were estimated as the difference between the average age of death in the model and a retirement age of 64, or the life expectancy in that country and year, whichever was lower. Costs were inflated to 2023 USD, with future costs discounted at 3% per annum as a standard method.

The impact of ORI on reducing the risk of large outbreaks was also estimated, by comparing the distribution of cumulative cases across outbreaks in the data to the distribution of cumulative cases across outbreaks in the no ORI scenario. For the purposes of this sub-analysis, cumulative cases in the no ORI scenario for each outbreak were approximated as the median from the no ORI counterfactual simulations.

#### **Results**

##### *Baseline*

The modelled cumulative cases and deaths for the baseline scenario is shown for each outbreak considered in the analysis in Figure S17. Outbreaks are grouped by baseline immunity and the corresponding boxplots are coloured by the model-estimated transmission setting. The data points are scaled to the vaccine-eligible population in the model. The model indicates that bigger outbreaks are more likely to occur in settings with lower baseline immunity and higher transmission settings. The largest outbreak, observed in South Sudan in 2018, was modelled using a 40% baseline immunity and was estimated to have occurred in a medium-high transmission setting.

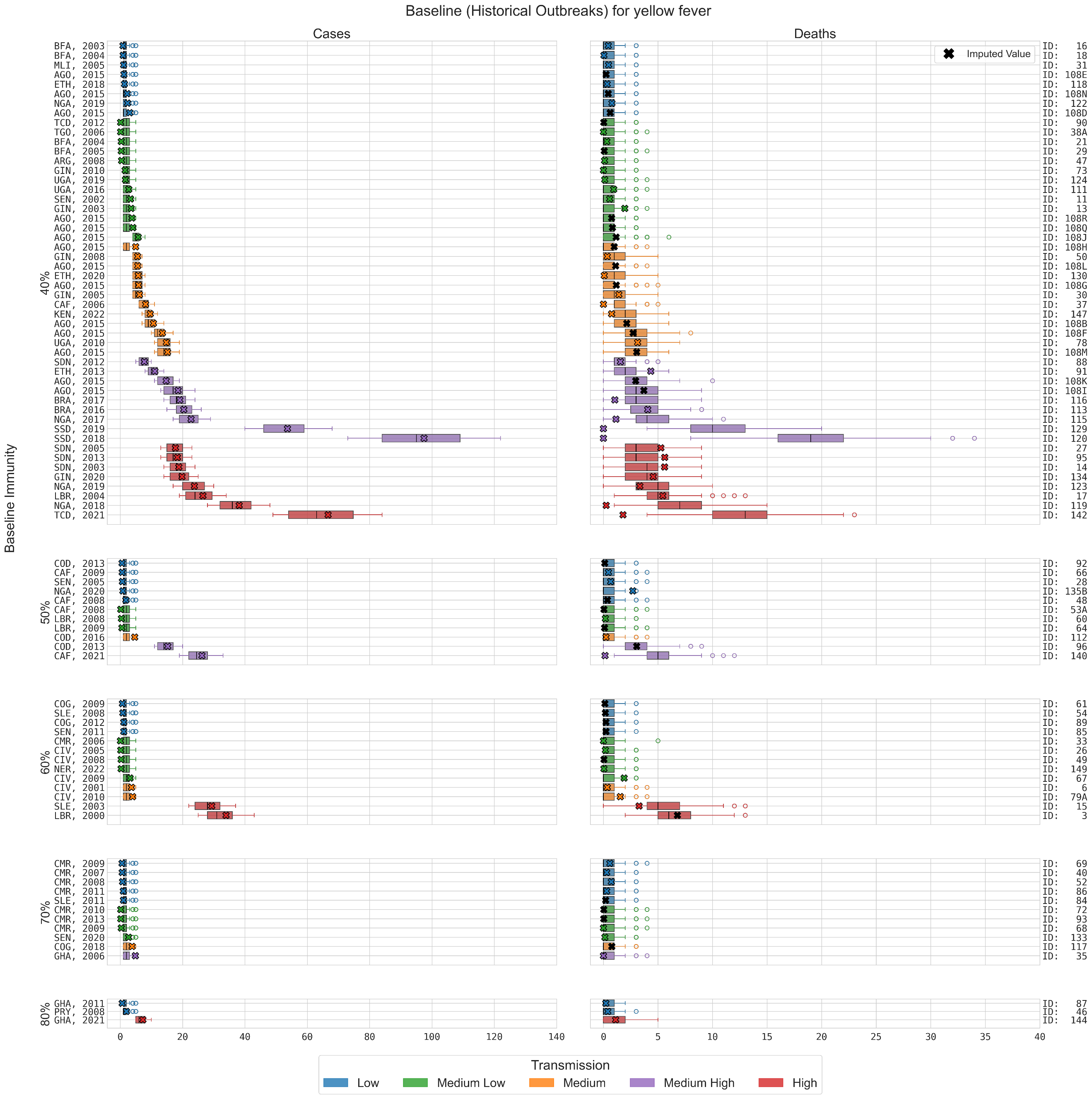

**Figure S17: Distribution of cases and deaths for each outbreak for model simulations retained after filtering.** Simulations which did not match within 25% of cumulative cases of historical outbreak were rejected. Crosses represent the data for each outbreak, scaled to a 50K population, and the colours represent the model-estimated transmission.

##### *Baseline vs. No ORI*

The distribution of simulated outcomes under the No ORI scenario is typically wider than the equivalent Baseline scenario and almost always larger (Figure S18).

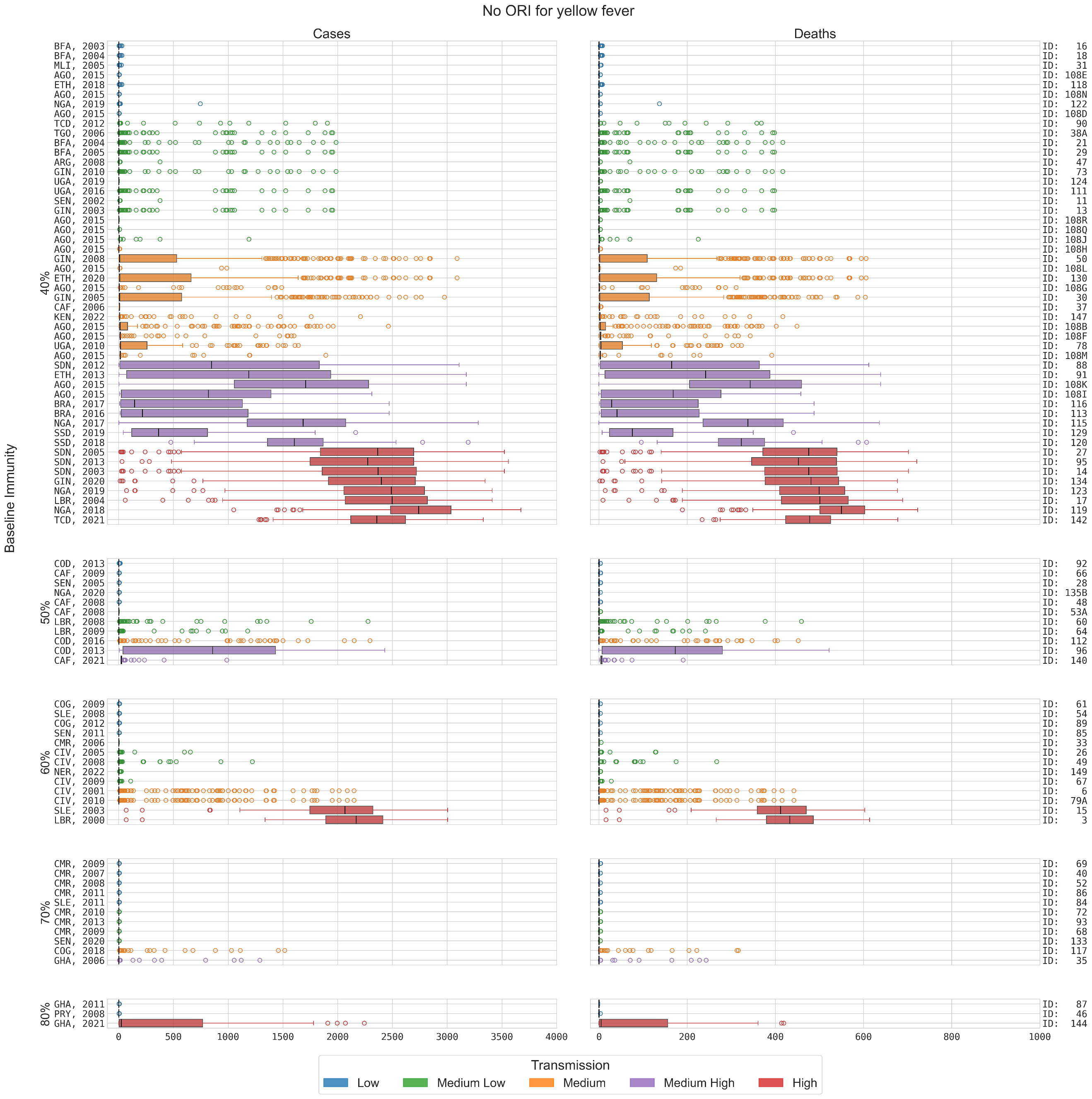

**Figure S18: Distribution of cases and deaths for each outbreak for counterfactual simulations with no ORI implemented.** Counterfactual simulations use equivalent transmission parameters and infection seeding to the matched baseline simulations.

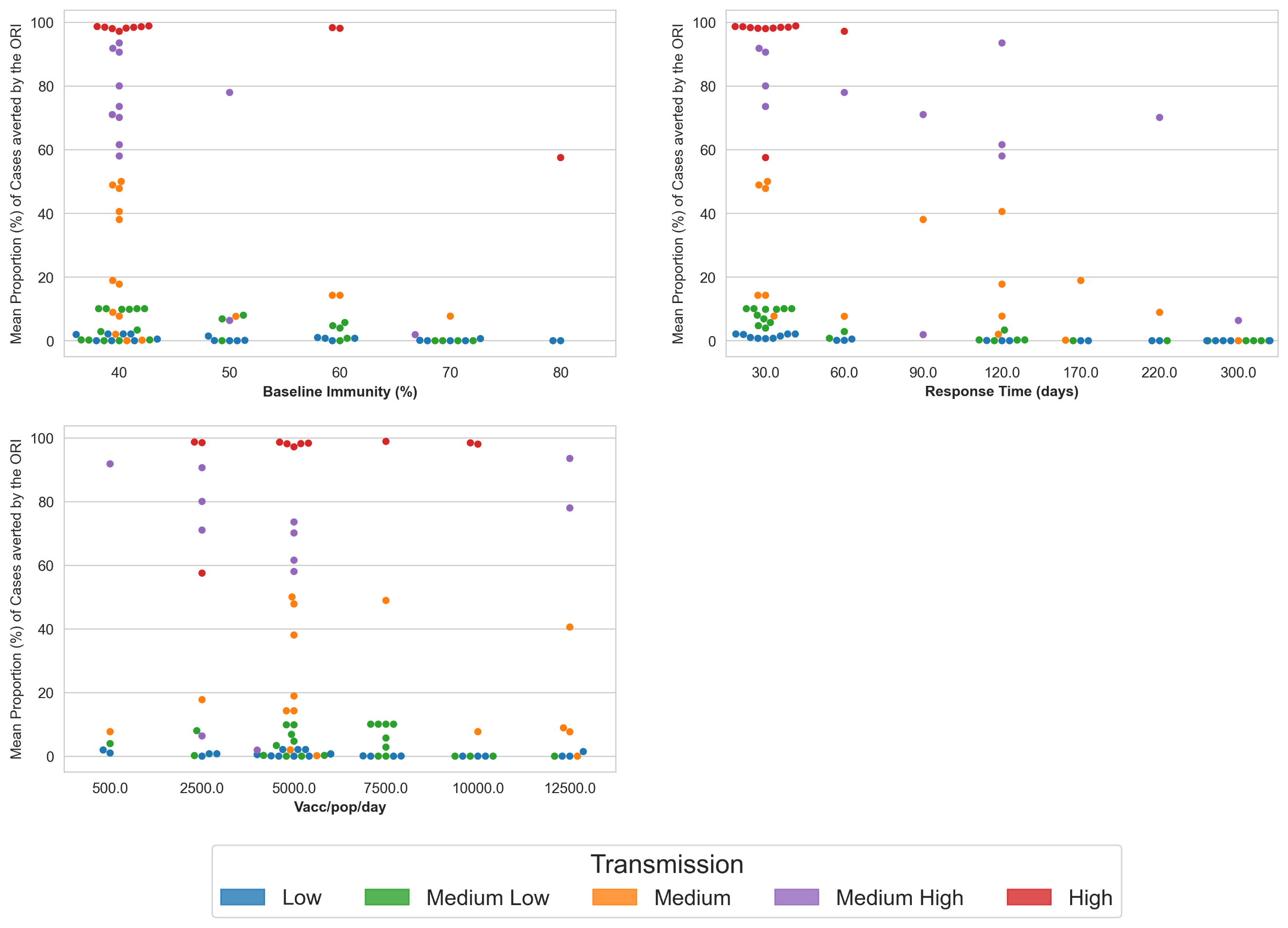

**Figure S19: ORI Impact estimation by lattice dimension**: Scatterplots showing the mean proportion of cases averted by the ORI, coloured by the assigned transmission level for each (fixed) lattice dimension.

**Table S9:** **Summary of impact estimation in 5-year increments.** Please note 35% of outbreaks are missing the number of observed cumulative deaths, which is therefore estimated using the calibrated mortality rate. Estimated Cases/ Deaths/ DALYs/ societal costs averted are summarized as mean (95% uncertainty interval).

|  | **# outbreaks** | **Observed cases** | **Observed/ Estimated deaths** | **Cases averted** | **Deaths averted** | **DALYs averted** | **Costs averted** |
| --- | --- | --- | --- | --- | --- | --- | --- |
| 2000 | 1 | 104 | 21 | 6428 (3871 – 8986) | 1296 (774 – 1818) | 54,483 (32,551 - 76,416) | $15·6M ($9·35M – 21·9M) |
| 2001-2005 | 15 | 1864 | 493 | 167K (150K – 189K) | 34,064 (30,205 - 37,924) | 1·44M (1·28M – 1·60M | $902M ($792M – 1011B) |
| 2006-2010 | 24 | 488 | 118 | 7269 (3092 – 11,445) | 1449 (615 – 2282) | 56,627 (24,258 - 88,995) | $38·4M ($13·8M – 62·9M) |
| 2011-2015 | 26 | 4907 | 1017 | 381K (320K – 443K) | 76,509 (64,155 - 88,864) | 3·36M (2·82M – 3·91M) | $5·87B ($4·90B – 6·85B) |
| 2016-2020 | 17 | 18,648 | 1227 | 830K (665K – 995K) | 166K (133K – 199K) | 7.27M (5.62M – 8.92M) | $23·5B ($14·0B – $32·9B) |
| 2021+ | 5 | 3804 | 111 | 102K (84K – 121K) | 20,554 (16,875 - 24,233) | 791K (645K – 936K) | $420M ($289M – 550M) |
| **Total** | 88 | 29,815 | 2988 | 1·50M (1·42M – 1·58M) | 300K (284K – 316K) | 13·0M (12·2M – 13·8M) | $30·7B ($26·5B – 34·9B) |

ORI also reduced the risk of large outbreaks. There was a wide variation in outbreak size within the data, with 42% of outbreaks having fewer than 25 cumulative cases, 30% having 25 – 100 cumulative cases, and 7% having more than 1,000 cases. However, with no ORI, the percentage of outbreaks with more than 1000 cumulative cases increased from 7% to 20%.

#### **Key messages**

Of the 88 yellow fever outbreaks over 2000-2023 where an ORI occurred and sufficient data was available for analysis, we estimate that the ORI averted a cumulative:

- 1·42M – 1·58M cases, compared to ~29,815 observed
- 284K – 316K deaths compared to ~2988 observed/estimated^[2]^
- 12·2M – 13·8M DALYs and
- USD 26·5B – 34·9B societal economic costs

The ORI was found to have higher impact in settings with high transmission and with lower baseline immunity, highlighting the importance of routine vaccination campaigns.

The presence of ORI was also estimated to have:

- Reduced the percentage of outbreaks with more than 1000 cumulative cases from 20% to 7% (i.e. from eighteen to six)
- Increased the percentage of outbreaks with fewer than 100 cases from 65% to 72% (i.e. from 57 to 63)

#### **Limitations**

- Uncertain case fatality rate: The model relies on 20% based on the observed mortality rate in the dataset. However, literature reports a mortality rate of 39%^46^. If the case fatality rate is greater than 20%, then the model may have underestimated deaths and vaccine impact.
- Uncertain baseline vaccine coverage: WUENIC data used for estimating the baseline immunity describes the percentage of surviving infants who received one dose of yellow fever vaccine in countries where yellow fever is part of the immunization schedule for children or is recommended in at-risk areas^43^. Additionally, there is uncertainty around the vaccine coverage due to over-estimation of yellow-fever vaccination reporting, which is accounted for in the model. However, there is high uncertainty around sensitivity and specificity of reporting.
- Uncertainty around re-vaccination: Re-vaccination of a proportion of the target population is expected as prior determination of the vaccination status is challenging in LMICs. The model incorporates re-vaccination of agents; however, there is no change in immunity. The results might therefore underestimate the impact of the ORI.
- Climate covariates: Mosquito population is controlled by rainfall and temperature (e.g. dry and wet seasons, hot and cold periods). The model assumes urban outbreaks without any initial spillover force of infection.
- Modelling mild vs. severe: The model is not disaggregating asymptomatic vs. symptomatic because little data is available to inform differences in disease duration and infectiousness, and because testing is driven by severe vs. non-severe.

### **Supplement 5: Meningococcal Meningitis**

This supplementary section describes the context and more detailed methods for our meningitis model and the ORI impact analysis

#### **Background and motivation**

Meningitis is an often-fatal inflammation around the brain and spinal cord, caused most frequently by infection with bacteria, however it can also be caused by viral, fungal, or parasitic infection^47^. Multiple bacteria are known to cause meningitis, but for this analysis only *Neisseria meningitidis*, which causes meningococcal meningitis is considered^48^. While people of all ages can develop meningitis, its burden is highest in very young children^48-50^. It is estimated that there are around 1·2 million cases and 135 thousand deaths due to meningitis globally per year ^48, 49^, with most of the burden occurring in the ‘meningitis belt’ in sub-Saharan Africa. Transmission of bacterial meningitis is highly seasonal, with the dry season (December-June) in the meningitis belt increasing both the rate of transmission and the incidence of invasive disease^51^.

Most people infected with *Neisseria meningitidis* do not become symptomatic and will passively transmit to others as an asymptomatic carrier, with studies estimating that 1-35% of the population in the meningitis belt are colonized by the bacteria^52, 53^. The burden of disease is primarily in children and teens, with most invasive disease cases occurring in children under five years of age, and most asymptomatic cases occurring in young teens^48, 50, 53^. The case fatality rate for meningococcal disease is estimated to be around 5-15% in most settings^54^, but around 20% of survivors will develop long-term sequelae as a result of the infection, including blindness, loss of hearing, epilepsy, or other motor/cognitive impairments^54, 55^.

Due to the high cost of the vaccines, during outbreak responses vaccine delivery is highly age-targeted, and doses are typically delivered to people either 1-29 years or 2-29 years as they carry the highest burden of disease. Most historical responses have used multivalent polysaccharide vaccines which protect against invasive disease but do not impact asymptomatic infection, but since 2019 some outbreaks have been responded to with multivalent conjugate vaccines, which provide additional protection against asymptomatic carriage as well as invasive disease^56^. In order to estimate the historical impact of vaccines delivered in response to outbreaks of bacterial meningitis we have developed a dynamic transmission model which incorporates vaccination and recreates the age-specific burdens of disease and its notable seasonality.

#### **Model overview**

The *Starsim* framework was used to create an agent-based model of meningitis among humans^1^, with states for susceptible, exposed, infected (symptomatically and asymptomatically), and recovered agents (Figure S20).

Agents in the model represent humans, who begin as susceptible, and each day have a probability of becoming infected that depends on their immunity status and is proportional to the prevalence of infection among the population. They are assigned an age (which affects infection outcomes and vaccine targeting), household contacts and community contacts. Additionally, agents can receive vaccines with characteristics matching either a conjugate or polysaccharide multivalent vaccine, depending on which was used during the outbreak under consideration^56-59^. Both susceptible and vaccinated people can become infected at a rate that is proportional to dynamic prevalence, however vaccinated people have reduced risk due to vaccine protection. Infection is primarily asymptomatic, with only a proportion of cases developing invasive meningococcal disease (IMD), and vaccination will either protect only against IMD (polysaccharide vaccines) or against both IMD and asymptomatic infection (conjugate vaccines). Following infection, people have an incubation period before becoming infectious and can present either symptomatically or asymptomatically. Asymptomatic carriers will eventually recover and clear their carriage of the bacteria, and symptomatic cases can either die or recover. The model's progression and transmission pathways are based on structures used in other modelling studies^60, 61^. Symptomatic humans in the model are identified as a suspected case with an assumed 50% probability per day once they develop symptoms, so most cases will be found within one to two days.

As the transmissibility and invasion rate of bacterial meningitis is highly dependent on seasonality^51^, the model assumes a sinusoidal forcing function which increases the probability of transmission and the development of invasive disease for one half of the year (aligning with the ‘dry season’ in the African meningitis belt) and decreases the probability for the other half of the year:

$\beta\left\{ t \right\}=\beta(1+0.6\cos\frac{2\pi t}{365})$ ,

where $\beta$ is the transmissibility parameter and $t$ is the daily step in the model. The form and parameterisation of this function are based on the methods of Karachaliou et al.^60^ Additionally, as asymptomatic infection is known to be the main driver of transmission^62^, and that approximately between 1% and 35% of the population in the meningitis belt is known to be carrying *Neisseria meningitidis* on average^52, 53^, we assume that the background carriage rate at the start of the epidemic season is a significant driver of outbreak size (which is a theory which has been discussed before^63^), and use it as a lattice parameter to be sampled during calibration.

The duration of vaccine immunity was set to well beyond the scope of the model period as it is assumed that no waning of immunity effects would be relevant over the outbreak period. Table S10 details the parameters used within the model, and their source or justification.

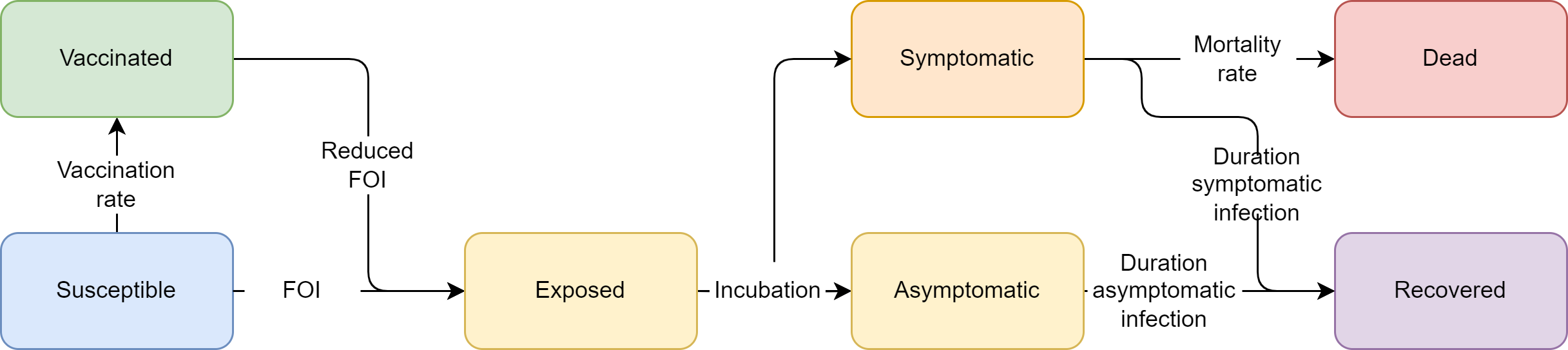

**Figure S20:** **Meningitis model schematic**. The Starsim framework was used to develop an agent-based model of meningitis among humans (S-E-I-R).

**Table S10: Meningitis model parameter values and sources.**

| Parameter | Value | Source |
| --- | --- | --- |
| Population parameters |  |  |
| Population age distribution | Empirical distribution | United Nations, Department of Economic and Social Affairs, Population Division^2^; averaged over countries with outbreaks used in this analysis. |
| Household size distribution | Empirical distribution | United Nations, Department of Economic and Social Affairs, Population Division^2^; averaged over countries with outbreaks used in this analysis. |
| Mean ‘community’ contacts per day | 11 | Prem et al.^7^; averaged over all non-household contact rates and all ages, for all countries with outbreaks used in this analysis. |
| Disease parameters |  |  |
| Average duration of exposed period | 2 days | Model uses a lognormal distribution fit to results of analysis by Bosis et al.^64^ |
| Average duration of asymptomatic infection | 90 days | Mean is an estimate from Blakebrough et al.^65^ and aligns with other recent estimates^66, 67^. Model uses a negative binomial distribution of to achieve a long tail as carriage durations vary significantly in the literature |
| Average duration of symptomatic infection (to recovery) | 7 days | Aligns with Karachaliou et al.^60^ MenA compartmental model. Model assumes a normal distribution with std=1 |
| Average duration of symptomatic infection (to death) | 8 days | Model uses a normal distribution fit to range of durations discussed by Sharew et al.^68^ |
| Probability of invasive disease | ~6% | The model uses age specific probabilities for invasive disease based on age-based risk estimates from Rivero-Calle et al.^50^ and estimates of IMD incidence by age from CDC.^69^ But the average probability of IMD is calibrated to match data |
| Probability of death given symptomatic disease | 7% | Calibrated value. The case fatality rate is 5-15% according to literature^48, 51, 54^. |
| Vaccine protection against IMD | 90% | Effectiveness across A,C,Y,W-135 estimated to be 85-100% according to Martinez et al.^57^, and 90% according to Daugla et al.^59^ |
| Vaccine protection against asymptomatic carriage | Polysaccharide: 0% Conjugate: 41% | No evidence that polysaccharide vaccines protect against carriage, for conjugate vaccines we aligned with value from Hadley et al.^56^ |
| Health economics parameters |  |  |
| Disability weights for meningitis infection | Acute disease: 0·133 Hearing loss: 0·074 Epilepsy: 0·263 Motor and cognitive impairment: 0·203 | Global Burden of Disease Disability Weight estimates^3^. Incidences of long-term sequelae estimated from Voss et al.^55^ |
| Average life expectancy | Specific to country and year of outbreak | United Nations, Department of Economic and Social Affairs, Population Division^2^. Used to estimate years of life lost. |
| Gross Domestic Product (GDP) per capita* | Specific to country and year of outbreak | World Bank, World Development Indicators^5^. |
| Discounting | 0% for DALYs; 3% for costs |  |

*Inflated to 2023 USD using average annual inflation rates since 2000^11^.

#### **Outbreak data**

Meningitis outbreaks have been recorded by the WHO in its *Weekly Epidemiological Record* and *Disease Outbreak News* since 2000, the MenAfriNet *Meningitis Weekly Bulletin*, and academic papers, with information on epidemiological and programmatic responses available online^12, 32, 70, 71^.

Between 2000 and 2022 there were 236 recorded outbreaks across countries in and around the African meningitis belt. Three outbreaks were not considered in this analysis because they occurred in a highly specific setting such as a prison, the reported case numbers and population size were inconsistent with the outbreak declaration threshold of 10 cases per 100k population, or because they are ongoing. Prior to 2013 most of these outbreaks were due to serogroup A, however due to the success of the MenAfriVac the incidence of meningitis A outbreaks in the region rapidly diminished. Since 2017 only one of the 65 outbreaks in our dataset is recorded as having cases with serogroup A. Due to the effective management of meningitis A, we have considered only outbreaks of other serogroups in this analysis, and 24 outbreaks were recorded as having received a vaccine response, with all having sufficient response data for inclusion. Data on cumulative cases, cumulative deaths, the target population size, and vaccines delivered per outbreak were the most complete, while information about the response time and duration of the outbreak was the most incomplete (Table S11). The recorded outbreaks varied widely in scale: 1-56 thousand cumulative cases, 73 thousand-2 million vaccines delivered, and 8-162 days to respond.

**Table S11: Summary of outbreak data used for meningitis analysis.**

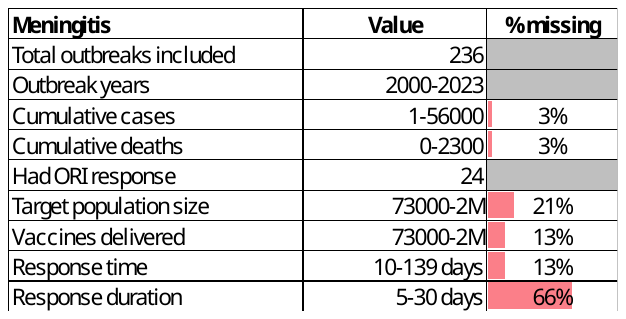

The following attributes were ascribed to each outbreak:

- Target population size: Directly from data, otherwise estimated based on number of vaccines (using linear relationship between number of vaccines and target population, fitted to all data with a target population and number of vaccines).
- Response time: Directly from data (time between outbreak declared and vaccine response started), which was then assigned to the closest of 24, 40, 50, or 65 days for simulation.
- Vaccine doses delivered: Directly from data where available, otherwise estimated based on target population size (using linear relationship between target population size and number of doses).
- Response duration: Directly from the data where available, otherwise based on average response duration from outbreaks in the same or nearby country.
- Response rate of vaccination: Directly from data where available, otherwise estimated based on target population size (using linear relationship between target population size and number of doses), which was then assigned to the closest of 900, 1700, 2200, or 3500 vaccines per day for simulation.
- Vaccine type used: Directly from data where available (stating either polysaccharide or conjugate vaccines), otherwise assumed to be polysaccharide as it is the standard type used.
- Target age group: Directly from data where available (stating the age ranges targeted during the response), otherwise assumed to be 1-29 years as it is the standard.

#### **Model population for simulated outbreaks**

The model population for each outbreak simulation represents the entire population of the specific geographic location where the outbreak occurred, based on the ‘target population size’ from the outbreak response data (i.e., people identified as being eligible for vaccination after the outbreak was declared). For computational reasons the model contains a maximum of 50,000 agents, and so for outbreaks with larger population sizes a scaling factor was used such that one agent in the model represents multiple people, based on the target population size and the proportion of the population eligible for the vaccine.

The model population was parametrized by age structure and household size distribution from the United Nations, Department of Economic and Social Affairs, Population Division^2^, and age-specific household contact rates from Prem et al.^7^. Each of these distributions and rates in the model were estimated using the average of the distributions and rates for each country with an outbreak included in the analysis. The age structure and household sizes are used to assign agents household contact networks, which are important for transmission in the model. The model also randomly generates ‘community’ contacts between agents, which have a much lower risk of transmission compared to household contacts^67^. These networks are randomly generated at each time step (representing a day in the model). Vaccine coverage, transmission risk and disease outcomes were modelled to vary by age, as the vaccines are highly age-targeted and both IMD and asymptomatic carriages incidences are well understood^48-50, 52, 53^.

#### **Diagnosis of cases, outbreak declaration and ORI**

Agents within the model are assumed to be detected as suspected cases of IMD (henceforth ‘cases’) within one day of developing symptoms, and we assume that all cases are detected. An outbreak is declared in the model after the detection of 5 symptomatic agents within a week, and once this occurs the ORI will begin after N days, where N is the response time for a given outbreak. This threshold aligns with the typical threshold of 10 cases per 100,000 population used for districts with a population greater than 30,000. Once the ORI begins, vaccines matching the characteristics of either multivalent conjugate or polysaccharide vaccines (depending on which was used for the given response in the data) will be targeted to the defined age group for the response, either 1-29, 2-29, or 1-49 years.

#### **Calibration**

Calibration involves estimating the transmissibility of meningitis in the model to produce outbreaks of a sufficient size, as well as the probability of death given symptomatic disease to capture the observed case fatality rate. Given well known variations in incidence of both invasive disease and asymptomatic carriage by age, the age-based susceptibility of humans to infection and developing symptoms was adjusted in the model to reproduce the typical age distributions observed in the literature^48-50, 52, 53^, seen in Figure S21. The model distributions from calibrated simulations with different vaccine response parameters can be seen in Figure S22. All other relevant model parameters were constrained by estimates from the literature. The strict seasonality of outbreaks was reproducible using a sinusoidal forcing function, using the same methods as work done by Karachaliou et al.^60^

Following this initial calibration process, for a given response time and vaccination rate, outbreak simulations could be run to produce a range of stochastic outcomes that could be compared to observed outbreaks. However, these simulations did not always align with the data, with differences potentially explained by climate, pre-existing immunity, and other factors that influence heterogeneity in transmission across settings. To account for this, an additional calibration variable was introduced to specify the rate of asymptomatic carriage in the model population before the increased seasonal transmission starts an outbreak. As transmission is known to be driven by asymptomatic carriers, and the prevalence is known to be highly variable during both endemic and epidemic periods, it is used as a free parameter to drive early epidemic growth rates in the model and produce the wide range of outbreak sizes observed in the well-defined epidemic season.

The range of initial prevalence values selected for the lattice to reflect the range of prevalences observed during endemic periods (0·25%, 1%, 2·5%, 5%, 7·5%, and 10%)^63^. Figure S23 demonstrates how the initial carriage prevalence drives the initial growth, time to peak, and final size of the simulated outbreaks, while still being constrained to a 4–6-month period of high IMD burden. Simulations initialised with an asymptomatic prevalence of 0.25% maintain a low level of asymptomatic disease and only produce a small increase in IMD cases due to the increased invasion rate during the epidemic season. However, simulations with higher initial prevalences tend to grow rapidly and produce noticeable peaks in both symptomatic and asymptomatic infections.

As the simulations are initialised with a non-zero carriage prevalence, we are in effect assuming that there was some level of endemic transmission occurring prior to when the model begins, which produced the assumed prevalence by the start of the epidemic season. This assumption means that we are ignoring the impact of population immunity which may have been developed prior to the epidemic season, and not accounting IMD cases accrued during the endemic period. These are significant simplifying assumptions, but as the rate of invasive disease outside of the epidemic season is known to be low and the model is calibrated against cases which occurred during a given epidemic season, we do not expect it to noticeably impact the calibration. The cumulative number of IMD cases and deaths produced across the range of initial prevalences allow us to reproduce the observed burdens in our outbreak data, and the estimates of increased asymptomatic prevalence during an outbreak align with estimates from the literature^52, 63^.

To produce the baseline simulations the model was initialized with a value for response time (24, 40, 50, or 65 days), vaccination rate (900, 1700, 2200, or 3500 vaccines per 50,000 population per day), initial carriage prevalence (0·25%, 1%, 2·5%, 5%, 7·5%, or 10%), vaccine type (polysaccharide or conjugate), and vaccine target ages (1-29 years, 2-29 years, or 1-49 years). Stochastic outbreak simulations were produced by running the model 1000 times for each outbreak in the dataset.

Observed outbreaks (i.e. with a given response time and vaccination rate) were then classified as having had different initial carriage prevalences based on a maximum-likelihood estimate of the total number of cumulative cases in the outbreak (see Kernel Density Estimation) (Figure S22; boxplots representing the range of simulated outcomes and crosses representing the observed outbreaks where they have occurred).

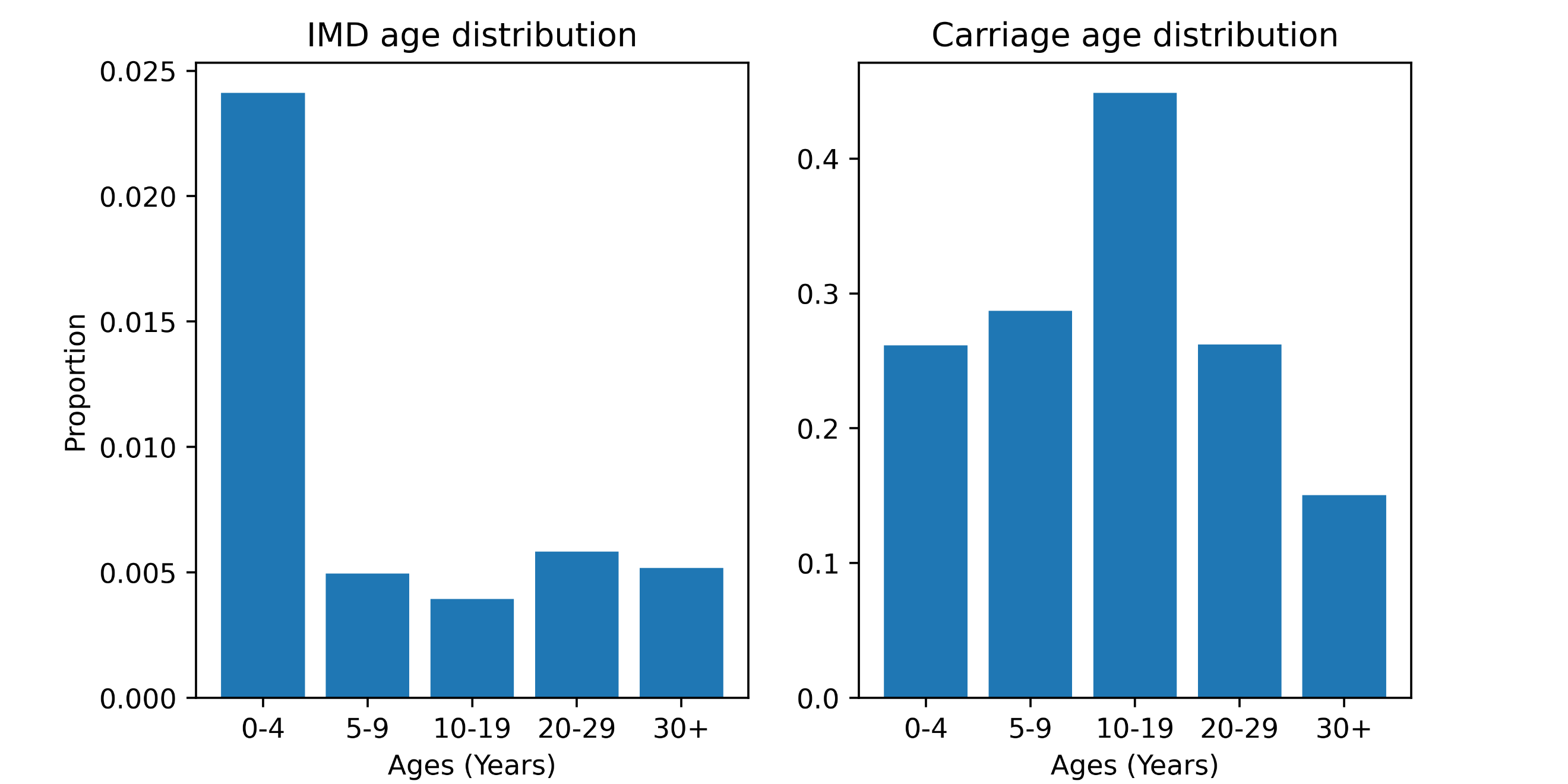

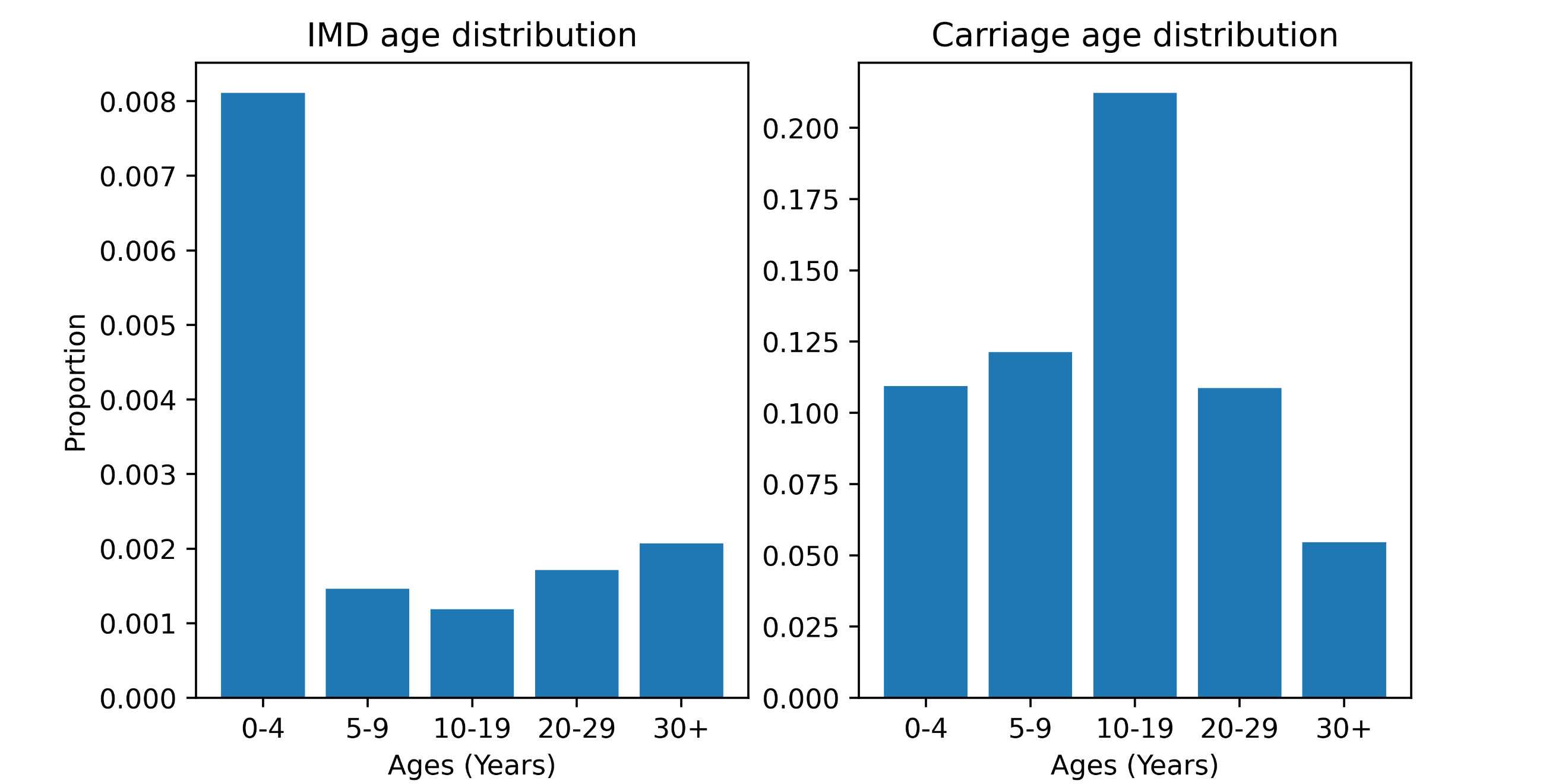

**Figure S21: Examples of IMD case and asymptomatic infection distributions from calibrated model simulations with a vaccine response.** IMD burden is highly focused in children less than one or two years of age (depending on how the target population for the response is defined), and people over 29 years of age because they typically do not receive vaccines as a part of the response. Rates of carriage in the model peak in older children and teenagers, which reflects what is observed in carriage studies from Africa.

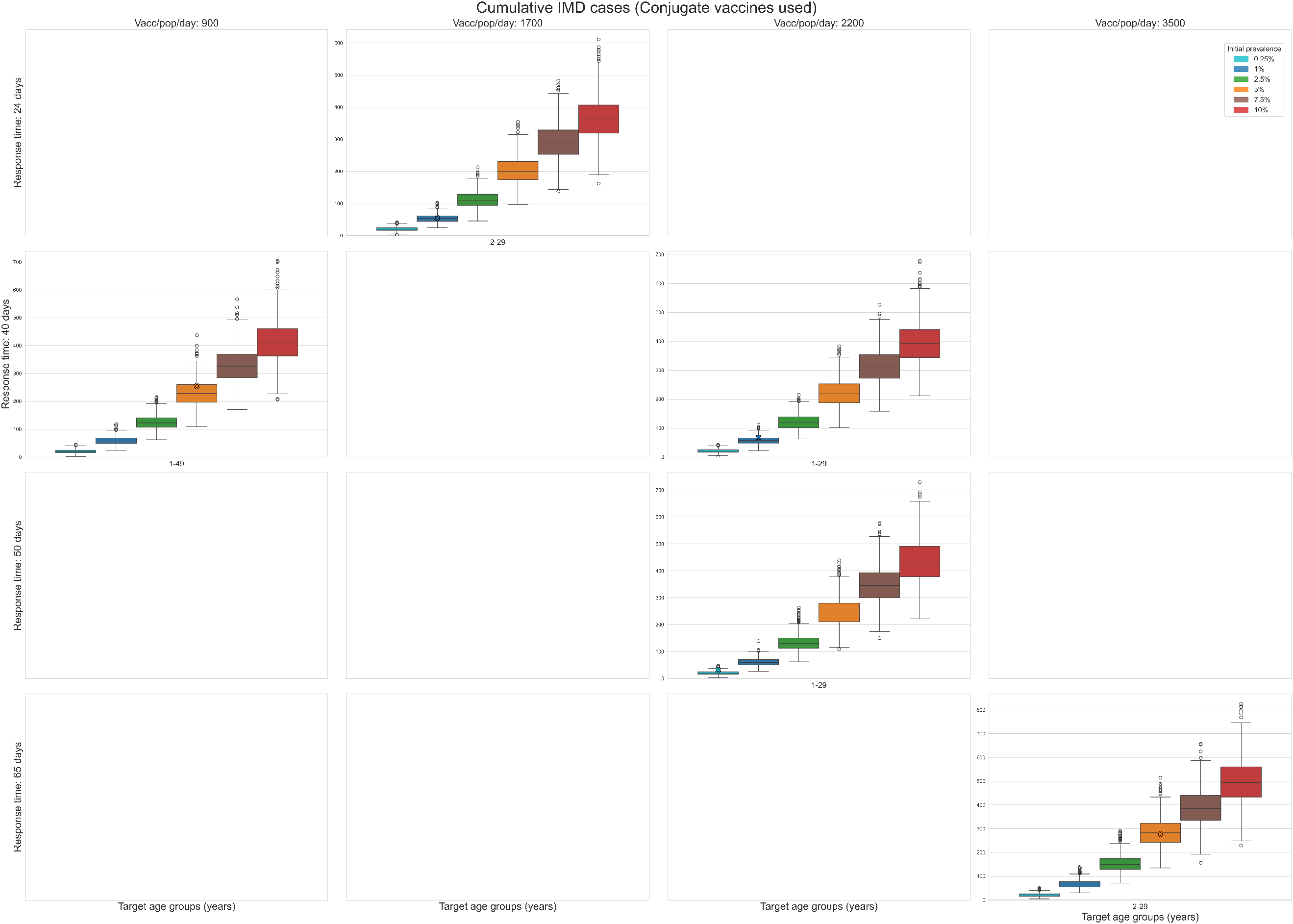

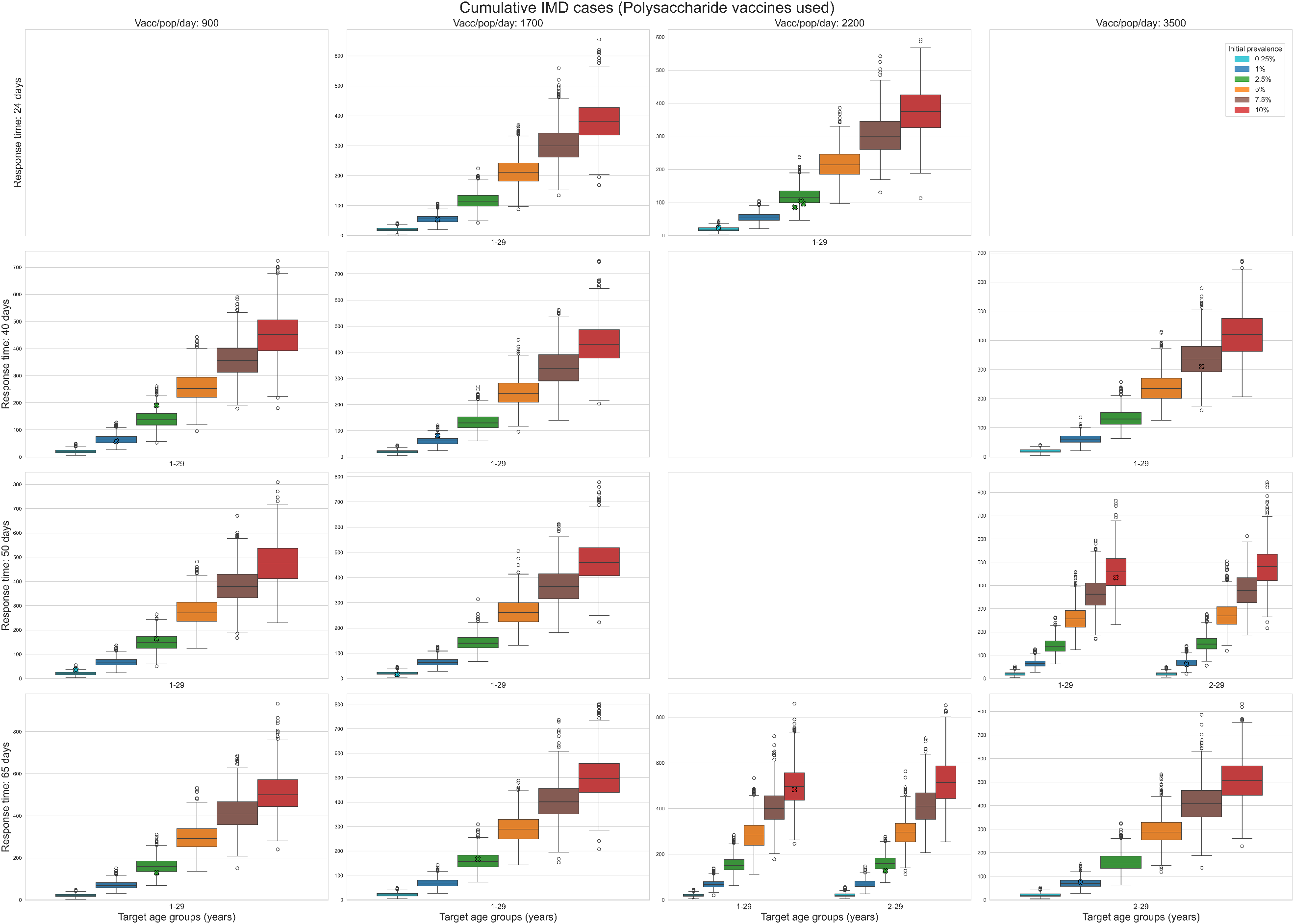

**Figure S22: Lattice of meningitis simulations before outbreak filtering was applied.** Each boxplot represents the distribution of total cases from a simulation run with response parameters defined by the lattice point. The crosses represent the cumulative suspected cases observed from an outbreak which has been assigned to the lattice point and scaled to a population of 50K. Subplot (a) shows the set of outbreaks which used polysaccharide vaccines, and subplot (b) shows the set of outbreaks which used conjugate vaccines.

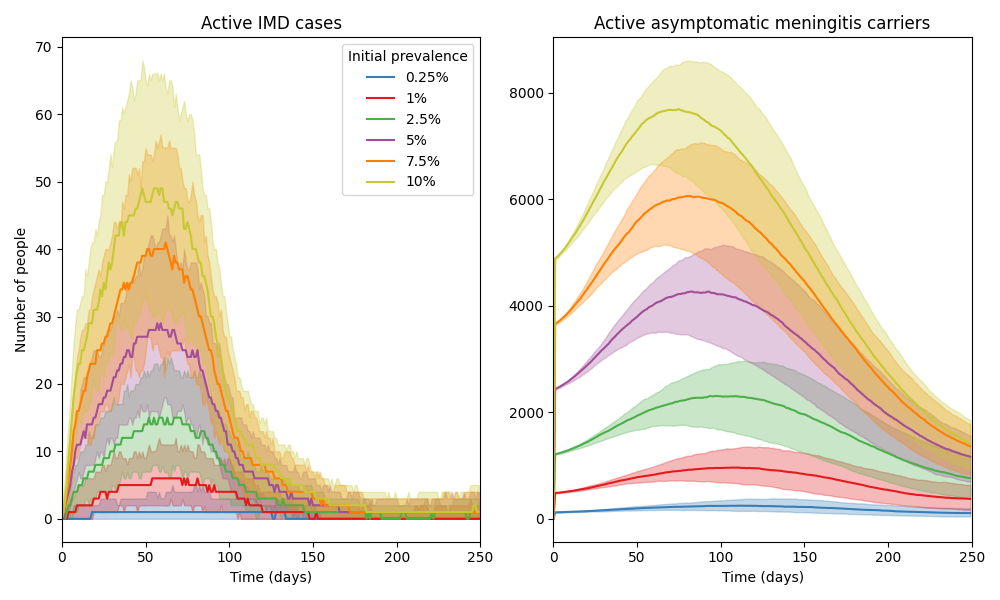

**Figure S23: An example set of model time series (using a population of 50K agents) after calibration, comparing the size and timing of outbreaks produced by different initial carriage prevalences.** The model is able to produce a wide range of outbreak sizes, all constrained to the epidemic season (assumed to be the first 6 months of model simulation), depending on the initial carriage prevalence. The initial prevalence drives the initial growth rate, time to peak, and final size of the simulated outbreaks. The colours in the figures represent the initial carriage prevalence used for the simulations.

#### **Scenarios**

For the purpose of running counterfactuals specific to the historical outbreaks, for each outbreak in the data, the 1000 simulations associated with the lattice point that the outbreak was assigned to were filtered to retain only those simulations where the cumulative cases were within +/- 25% of the reported cases (see Trajectory selection). Two scenarios were then examined:

- Baseline: ORI as occurred, using the filtered simulations from the lattice
- No ORI counterfactual: Simulations were run without any ORI intervention using the same filtered simulation seeds from the baseline scenario, such that everything was identical up until the date the ORI would have started.

#### **Outcomes**

For each outbreak and scenario, the distribution of cumulative cases and deaths across selected model simulations were recorded. The mean difference in outbreak size between scenarios and associated uncertainty in the mean were estimated from these collections of simulations using bootstrap resampling (i.e., for each outbreak to produce estimates of cases averted by ORI).

Total cases averted by ORI across all historic outbreaks was then estimated by aggregating the cases averted for each individual outbreak. As the outbreaks are independent, this was obtained by summing the cases averted per outbreak, with the variance estimated by summing the variances from each individual outbreak.

DALYs averted by ORI were estimated by multiplying cases averted by the disability weight per case and average duration of symptoms, plus years of life lost from deaths (for each outbreak, the average life expectancy in that country and year compared to the age of deaths in the model). Socio-economic costs averted by ORI were calculated by estimating productive years of life lost or lived with disability, and multiplying this by GDP per capita. Productive years of life lost were estimated as the difference between the average age of death in the model and a retirement age of 64, or the life expectancy in that country and year, whichever was lower. Costs were inflated to 2023 USD, with future costs discounted at 3% per annum as a standard method.

The impact of ORI on reducing the risk of large outbreaks was also estimated, by comparing the distribution of cumulative cases across outbreaks in the data to the distribution of cumulative cases across outbreaks in the no ORI scenario. For the purposes of this sub-analysis, cumulative cases in the no ORI scenario for each outbreak were approximated as the median from the no ORI counterfactual simulations.

#### **Results**

##### *Baseline*

The modelled cumulative cases and deaths for the baseline scenario is shown for each outbreak considered in the analysis in Figure S24. Outbreaks are grouped by the type of vaccine used in the response and the corresponding boxplots are coloured by the model-estimated initial asymptomatic carriage prevalence. The data points are scaled to the vaccine-eligible population in the model. The model indicates that bigger outbreaks are more likely to occur in settings with higher initial asymptomatic carriage at the start of the epidemic season. The fit of the simulations to the case data presented in Figure S24 differs from what is presented in Figure S22 (i.e., the median of the model simulations sits close to the data) as we are considering only the filtered simulations which align well with the observed cases. It should be noted that as the model applies a uniform probability of death given symptomatic disease (estimated from the average case fatality rate observed during calibration), if the fatality rate for a given outbreak differs then the model is unable to capture the cumulative deaths as well as the cumulative cases.

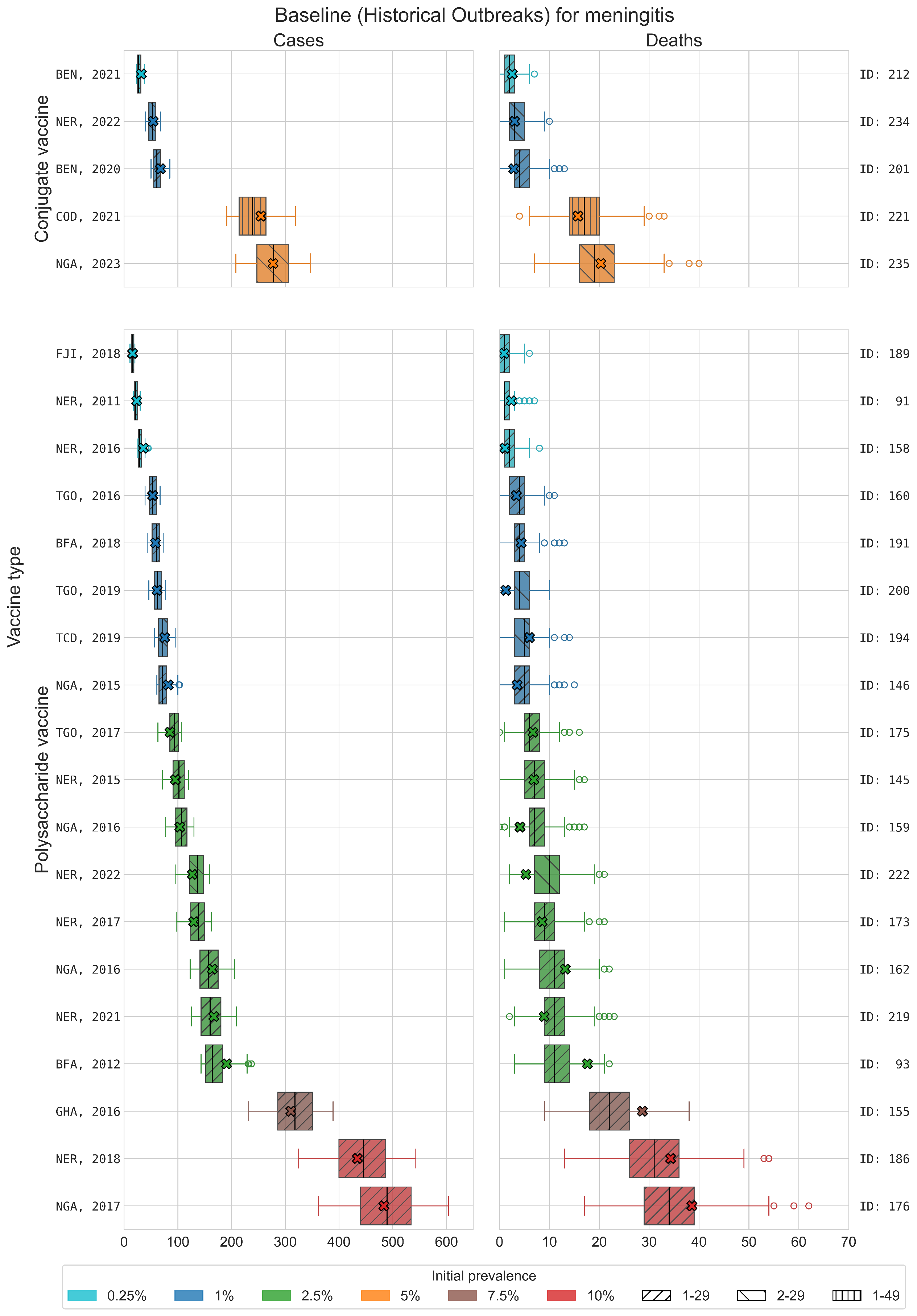

**Figure S24: Distribution of cases and deaths for each meningitis outbreak for model simulations retained after filtering.** Simulations which did not match within 25% of cumulative cases of historical outbreak were rejected. Crosses represent the data for each outbreak, scaled to a 50K population, and the colours represent the initial asymptomatic carriage prevalence.

##### *Baseline vs. No ORI*

The distribution of simulated outcomes under the No ORI scenario are much wider than the equivalent Baseline scenario, as they are not constrained by filtering against observed cases, and always larger (Figure S25).

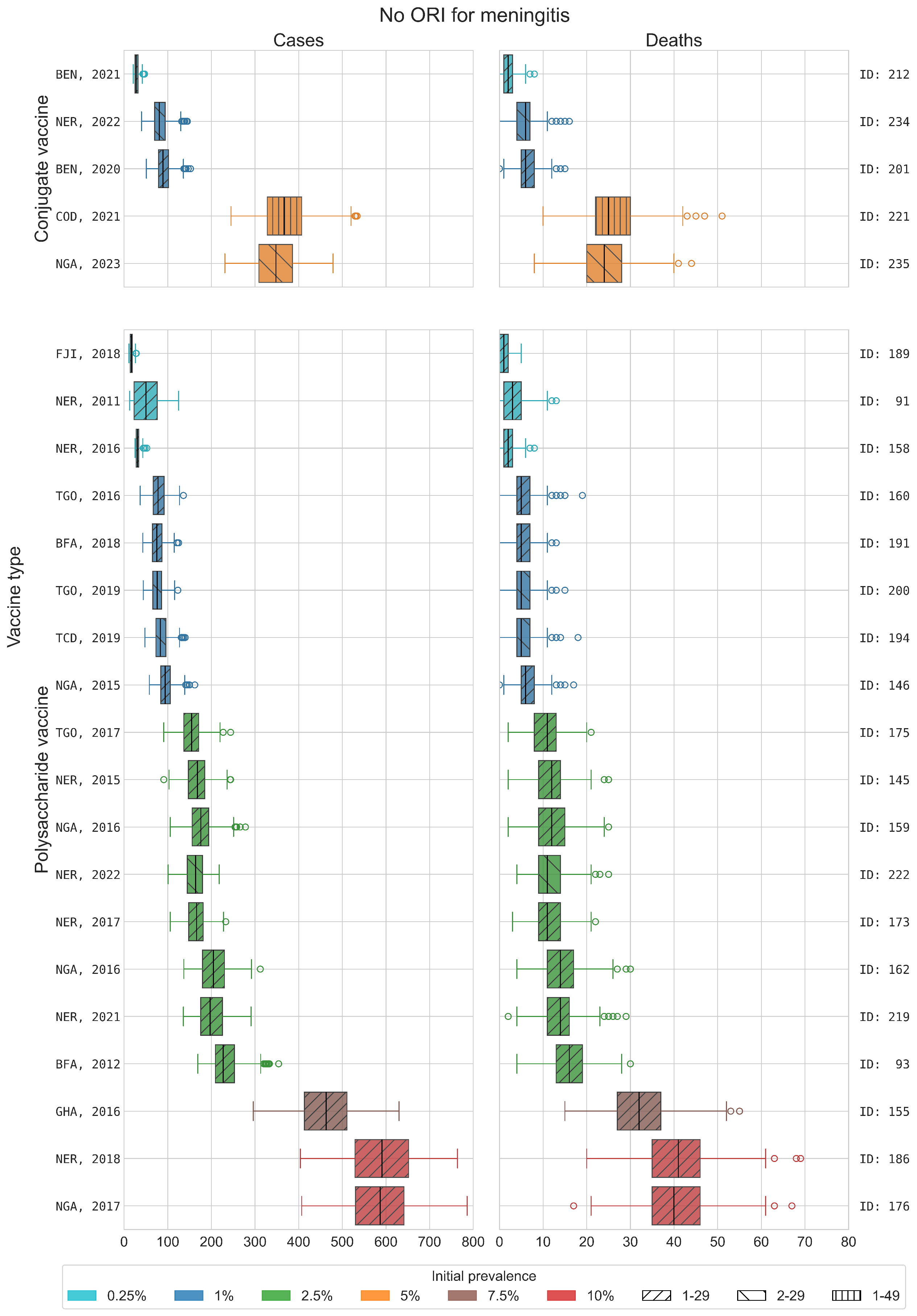

**Figure S25: Distribution of cases and deaths for each meningitis outbreak for counterfactual simulations with no ORI implemented.** Counterfactual simulations use equivalent transmission parameters and infection seeding to the matched baseline simulations. The colours represent the initial asymptomatic carriage prevalence.

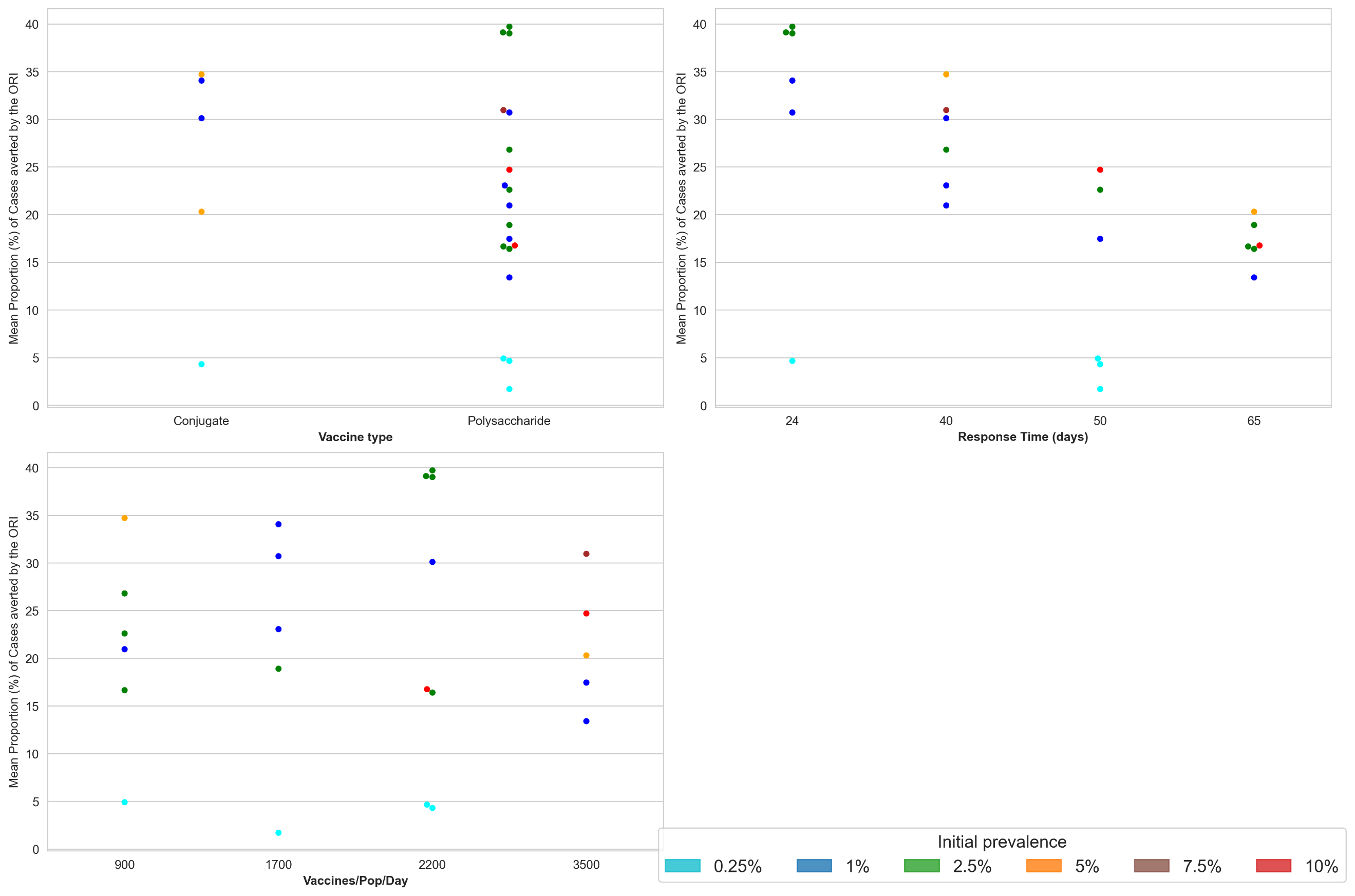

**Figure S26:** **Meningitis ORI Impact estimation by lattice dimension**: Scatterplots showing the mean proportion of cases averted by the ORI, coloured by the assigned transmission level for each (fixed) lattice dimension.

**Table S12: Summary of estimated ORI impacts across meningitis outbreaks in five-year periods.** Estimated Cases/Deaths/DALYs/societal costs averted are summarized as the mean and 95% uncertainty intervals.

| Years | # outbreaks | Observed cases | Observed deaths | Cases averted | Deaths averted | DALYs averted | Costs averted (2023 USD) |
| --- | --- | --- | --- | --- | --- | --- | --- |
| 2011-2015 | 4 | 11,130 | 801 | 6030 (4622 – 7437) | 454 (185 - 722) | 34,587 (21,313 – 47,862) | 17·8M (12·1M – 23·5M) |
| 2016-2020 | 14 | 33,804 | 2515 | 10,363 (9419 – 11,306) | 796 (601 – 991) | 55,542 (46,834 – 64,250) | 66·6M (55·4M – 77·8M) |
| 2021-2023 | 6 | 12,639 | 764 | 4867 (4127 – 5607) | 348 (223 – 473) | 22,740 (16,680 – 28,800) | 13·3M (10·5M – 16·0M) |
| Total | 24 | 57,573 | 4080 | 21,261 (20,268 – 22,254) | 1599 (1404 – 1794) | 113K (104K – 122K) | 96·6M (86·6M – 106·6M) |

ORI also reduced the risk of large outbreaks. There was a wide variation in outbreak size within the data, with 37.5% of outbreaks having fewer than 1000 cumulative cases, 54% having 1000 – 5000 cumulative cases, and one outbreak having more than 10,000 cases. However, with no ORI, the percentage of outbreaks with more than 2,500 cumulative cases increased from 29% to 42%.

#### **Key messages**

The vaccine responses delivered to 24 meningitis outbreaks from 2011-2023 averted significant burdens of disease, with more than half of the estimated impact being accrued between 2016-2020. Notably, responses are estimated to have been more effective when response times were faster.

We estimate that the ORI averted a cumulative:

- 20,268 – 22,254 cases, compared to 57,573 observed
- 1404 – 1794 deaths compared to 4080 observed
- 104K – 122K DALYs; and
- $86·6M – $106·6M societal economic costs

The presence of ORI was also estimated to have:

- Reduced the percentage of outbreaks with more than 2500 cumulative cases from 42% to 29% (i.e. from ten to seven)
- Increased the percentage of outbreaks with fewer than 1000 cases from 29% to 37·5% (i.e. from seven to nine)

#### **Limitations**

- Actual transmission might be heterogeneous and could be a factor that limits outbreak size: there are multiple sources of heterogeneity that the model cannot capture, across the populations and transmission networks. This includes
  - Highly connected contact networks (e.g., workplaces, schools, social groups) which could lead to additional transmission networks outside of households that the randomly generated community networks do not capture well. This may lead to the model underestimating the transmissibility of meningitis, particularly the lack of school and social networks which would increase the connectivity of the high carriage younger age groups and likely increase transmission.
  - Climactic heterogeneities which impact the timing, duration, and effect of the epidemic season in each outbreak. The model uses a simple period forcing function with fixed amplitude across outbreaks, which cannot capture these potential effects.
- High uncertainty around conjugate vaccine efficacy against carriage: only a small number of studies have investigated the efficacy of conjugate vaccines against carriage of *Neisseria meningitidis*. The source for the efficacy value used in the model aligns with the value used by others in the Vaccine Impact Modeling Consortium^60^, but if it is an inaccurate assumption then this would likely impact the results for the five outbreaks which used conjugate vaccines.
- Reporting of meningococcal cases: the model is calibrated to the cumulative suspected cases reported for each outbreak:
  - This may be inflated due to some suspected cases that are not meningococcal disease. If so, then the model may overestimate the transmissibility of meningitis which would produce overestimates of ORI impact.
  - On the other hand, reported cases are an underestimate of the true number of cases due to the limited availability of confirmatory testing. If so, then the model may underestimate the transmissibility of meningitis which would produce underestimates of ORI impact.
- Uncertainty around number of asymptomatic infections: there is high uncertainty around the proportion of infections which are asymptomatic as there is little data to inform this. The parameters in the model are constrained to the observed cases and deaths, and the carriage prevalence prior to and during outbreaks in the model align with ranges described in the literature, but these factors appear to be highly variable across outbreaks. It is known that the rate of invasive disease varies over time, but the degree of variation is unknown and it is unclear how to accurately parameterise it. As it is unclear whether the asymptomatic infection rate (and how it varies over time) is an under- or overestimate, it is unclear what impact this has on the results.
- Uncertainty around timing of infections over the year: the epidemic season in the meningitis belt is well-defined, and where possible the cumulative cases and deaths which were used for calibration across the set of outbreaks reflect the burden from the epidemic season. However, in some cases these data were not disaggregated by whether they occurred during the epidemic or endemic period and so the calibrations for some outbreaks may be overestimates. However, due to the typically low incidence of IMD during the wet season this impact is not expected to be high.

### **Supplement 6: Ebola**

This supplementary section describes the context and more detailed methods for our Ebola model and the ORI impact analysis

#### **Background and motivation**

Ebola is a deadly hemorrhagic fever virus which causes severe disease with a greater than 50% case fatality rate, and produces long-lasting sequelae in approximately 70% of cases even if they survive^72, 73^. Most outbreaks of Ebola are relatively small, but the exceedingly high fatality rate and frequent, debilitating effects in survivors mean that any detected cases of Ebola prompt a rapid response from health authorities. The large outbreaks which occurred in Western Africa in 2014 and the Democratic Republic of Congo in 2018 also indicate that Ebola can achieve significant levels of transmission if public health responses are not effective. Until recently, there was no vaccine available for Ebola and so outbreak responses relied on contact tracing, isolation, and quarantine of cases and their contacts. Since development of the Ervebo vaccine, and its first use during the 2014-2016 West African outbreak^74^, contact tracing is still a primary tool used during an outbreak response, and allows for targeted ring-based vaccination strategies to be implemented and protect the people at highest risk of infection.

Ebola is transmitted via contact with blood and other bodily fluids, and the virus survives long after the death of the host so contact with a corpse can also be a transmission pathway, and the use of safe burial practices is an important facet of outbreak response^75^. In order to estimate the historical impact of vaccines delivered in response to outbreaks of Ebola we have developed a dynamic transmission model which incorporates both contact tracing and vaccination, while also tracking the risk of infection from bodies which are not safely buried.

#### **Model overview**

The *Starsim* framework was used to create an agent-based model of Ebola among humans^1^, with states for susceptible, exposed, infected, recovered, and dead agents (not safely buried) (FigureS 27).

Agents in the model represent humans, and are assigned an age (which affects infection outcomes), household contacts and community contacts. Additionally, agents can receive vaccines with characteristics matching the Ervebo vaccine^76, 77^, which was used during the outbreaks under consideration. Both susceptible and vaccinated people can become infected at a rate that is proportional to dynamic prevalence, however vaccinated people have reduced risk due to vaccine protection. Following infection, people have an incubation period before becoming infectious, and after a period some infected agents will progress to more severe disease. Those who do not advance to a more severe disease state will recover, whereas those with severe disease can either die or recover. As Ebola can still be transmitted via contact with a corpse, the model includes a transmission pathway until burial occurs, however for people identified as Ebola cases a proportion of burials are assumed to be handled safely, which removes this risk of transmission. The model's progression and transmission pathways are based on structures used in other modelling studies^78-81^.

The duration of vaccine immunity was set to well beyond the scope of the model period as it is assumed that no waning of immunity effects would be relevant over the outbreak period. Table S13 details the parameters used within the model, and their source or justification.

In the model, contact tracing captures 95% of household contacts within 1 day, and 25% of community contacts in 1-2 days. These values are assumptions, calibrated to capture the numbers of contacts which are reported to have been successfully traced over outbreaks being considered^82-86^. Traced contacts are required to quarantine for 21 days, with PCR tests on day 1 and day 21.

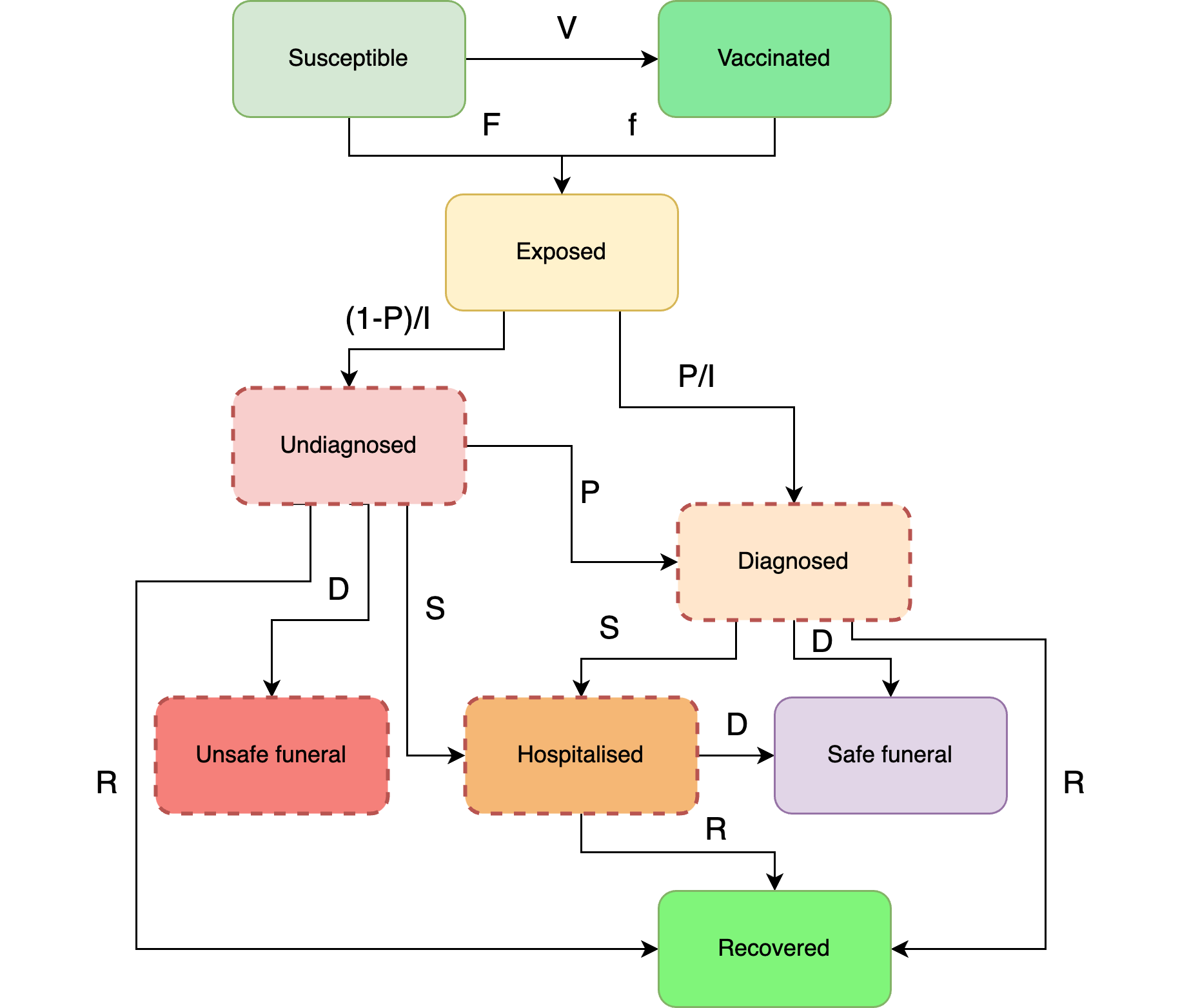

**FigureS 27: Ebola model schematic**. The Starsim framework was used to develop an agent-based model of Ebola among humans (S-E-I-R-D).

**Table S13: Ebola model parameter values and sources**

| Parameter | Value | Source |
| --- | --- | --- |
| Population parameters |  |  |
| Population age distribution | Empirical distribution | United Nations, Department of Economic and Social Affairs, Population Division^2^; using DRC data. |
| Household size distribution | Empirical distribution | United Nations, Department of Economic and Social Affairs, Population Division^2^; using DRC data. |
| Mean ‘community’ contacts per day | 14 | Prem et al.^7^; averaged over all non-household contact rates and all ages, using Congo as a proxy for DRC. |
| Disease parameters |  |  |
| Probability of developing severe disease | 70% | Proportion of cases which progressed to Stage 2 or 3 disease from Kangbai et al.^87^ |
| Probability of death, given severe disease | 55% | Calibrated value. Ebola case fatality rate is typically ~50%, but can range from 25%-90%^72, 73, 87^ |
| Average duration of exposed period | 12·7 days | Haas CN^88^. |
| Average duration of mild infection | 10·0 days | Assumption based on ranges and reported means in Kadanali and Karagoz^72^, Singh and Ruzek^89^, Legrand et al.^90^, and Simpson^91^. |
| Average time to develop severe disease | 6·0 days | Assumption based on reported 5–7-day ranges for time to develop fever and rash in Kadanali and Karagoz^72^ and Goeijenbier et al.^92^ |
| Average duration of severe infection (survived) | 10·4 days | Mean time in hospital for survivors from Hartley et al^93^. Aligns with convalescence time for survivors from Simpson^91^. |
| Average duration of severe infection (died) | 1·5 days | Mean time to death after symptom onset is 7·5 days from Centers for Disease Control and Prevention^94^. Subtract 6 days for average time to develop severe disease. |
| Average time to burial (unsafe funeral) | 2·0 days | Assumption based on parameter used in Legrand et al.^90^ |
| Vaccine protection against infection | 97·5% | Based on reported value by World Health Organization^77^. |
| Ebola PCR test sensitivity | 99% | Assumption, as the GeneXpert PCR tests seem to be considered the gold standard test. |
| Ebola PCR test delay | 1 day | Assumption. |
| Probability of successful contact tracing | Household: 95%  Community: 25% | Calibrated assumptions. |
| Time to trace contacts | Household: 1 day  Community: 2 days | Assumptions, 1 day is the minimum time given model time steps and community tracing is assumed to take longer. |
| Quarantine period | 21 days | Based on standard control methods for Ebola outbreaks^88^. |
| Contact tracing capacity | 25 people per day | Calibrated assumption, used to reduce the efficacy of contact tracing and allow larger outbreaks to occur, assuming contact tracers are overwhelmed during larger outbreaks. |
| Health economics parameters |  |  |
| Disability weights for cholera infection | Acute: 0·133  Chronic:  0·219 | Global Burden of Disease Disability Weight estimates^3^. Chronic infection is assumed to persist for one year. |
| Average life expectancy | Specific to year of outbreak | United Nations, Department of Economic and Social Affairs, Population Division^2^; using DRC data. |
| Gross Domestic Product (GDP) per capita* | Specific to year of outbreak | World Bank, World Development Indicators; using DRC data^5^. Used to estimate years of life lost. |
| Discounting | 0% for DALYs; 3% for costs |  |

*Inflated to 2023 USD using average annual inflation rates since 2000^11^.

#### **Outbreak data**

Ebola outbreaks have been recorded by the WHO in its *Disease Outbreak News* since 2000, with information on epidemiological and programmatic responses available online^32^. Between 2000 and 2022 there were 17 recorded outbreaks of *Zaire ebolavirus*, 53% of which were in the Democratic Republic of Congo. Two outbreaks were not considered in this analysis because they were much larger than any other recorded outbreak of Ebola and occurred in highly conflict-affected settings, which are known to have impacted health systems and outbreak response^95, 96^. As such, the model implementation would not have been able to capture their transmission and response features. 7 of the considered outbreaks were recorded as having received a vaccine response, with all having sufficient data for inclusion in the analysis. Data on cumulative cases, cumulative deaths, the time taken to start a response, vaccines delivered per outbreak, and the duration of the outbreak response were complete (Table S14). The considered outbreaks did not vary widely in scale: 1-264 cumulative cases, 550-40·9K vaccines delivered, and 4-14 days to respond.

**Table S14: Summary of outbreak data used for Ebola analysis.**

   
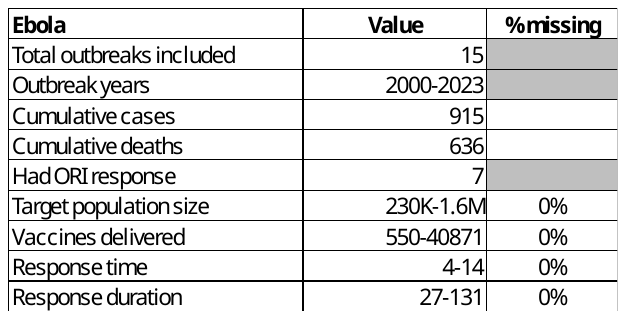

The following attributes were ascribed to each outbreak:

- Response time: Directly from data (time between outbreak declared and vaccine response started), which was then assigned to the closest of 4, 7, 10, or 14 days for simulation.
- Vaccine doses delivered: Directly from data.
- Response duration: Directly from the data.
- Response rate of vaccination: Directly from data, which was then assigned to the closest of 5, 10, 20, or 35 vaccines per 100,000 population per day for simulation.

#### **Model population for simulated outbreaks**

The model population for each outbreak simulation represents a subset of the population of the specific geographic location where the outbreak occurred. For computational reasons the model contains a maximum of 100,000 agents, each representing a single human in the subset population under consideration for each outbreak. As Ebola outbreaks are typically relatively small and contained by outbreak response even in the absence of vaccines, and because contact tracing plays such a key role in containment and vaccine targeting for a ring-based response, no scaling was applied to the agents in this model. 100,000 agents was considered large enough to allow for significant transmission, while not limiting growth in the counterfactual scenarios.

The model population was parametrized by age structure and household size distribution from the United Nations, Department of Economic and Social Affairs, Population Division^2^, and age-specific household contact rates from Prem et al.^7^.The age structure and household sizes are used to assign agents household contact networks, which are important for human-human transmission in the model. The model also randomly generates ‘community’ contacts between agents, which have a much lower risk of transmission compared to household contacts^97^. These networks are randomly generated at each time step (representing a day in the model). Transmission risk and disease outcomes were not modelled to vary by age, but vaccines were delivered to only people over the age of 18. The household size distribution and age distribution of humans within the model are informed by data from the Democratic Republic of Congo, as this is where most Ebola outbreaks occurred.

#### **Diagnosis of cases, outbreak declaration and ORI**

Symptomatic agents within the model are identified with a probability of 2% per day that they are symptomatic prior to outbreak detection, assuming low availability and access to diagnostic tests, and access to healthcare. Due to increased awareness after the outbreak is declared, this probability increases to 20% per day, assuming additional resources are directed to the affected population. An outbreak is declared in the model after the detection of a single symptomatic agent (henceforth a ‘case’), and once this occurs the ORI will begin after N days, where N is the response time for a given outbreak. All cases within the model will have their contacts traced and quarantined, and the case will undergo isolation to reduce risk of onward transmission. Once the ORI begins, a ring-based vaccination strategy will be implemented which will prioritise delivering vaccines to eligible contacts of known cases in the model. If the daily vaccination rate exceeds the number of contacts to be vaccinated then the excess doses will be delivered to eligible humans without additional targeting.

#### **Calibration**

For the calibration the model was initialised with a population of 100,000 agents. The overall transmission risk per contact (in household and community settings), the per-day probability of a symptomatic individual getting tested, the probability of contacts being successfully traced, and the number of seed cases were varied such that the distribution of model outcomes for cases, deaths, and total contacts traced was centered on the reported empirical data. All other relevant model parameters were constrained by estimates from the literature. The range of simulation outcomes is driven by the stochastic nature of the agent-based model, with the initialisation conditions randomising where infections are seeded and how contact networks are generated.

Once the transmission, mortality, testing, and contact tracing parameters had been calibrated, outbreaks were simulated by infecting three agents in the model at random, for a range of parameter lattice points representing vaccine response times (4, 7, 10, 14 days) and vaccination rates (5, 10, 20, 35 vaccines per 100,000 people per day). The set of seven outbreaks with a vaccine response and sufficient data were assigned to their nearest lattice point based on the observed response time and vaccination rate and compared to the distribution of model outbreak simulations (Figure S28). The calibration produces outcomes which align well with the seven outbreaks we considered, as all case counts fall within the range of cases produced by the model. For each lattice point, a variable number of simulations were run, depending on the number of outbreaks assigned to the lattice point and how frequently the model would simulate outbreaks of a similar size.

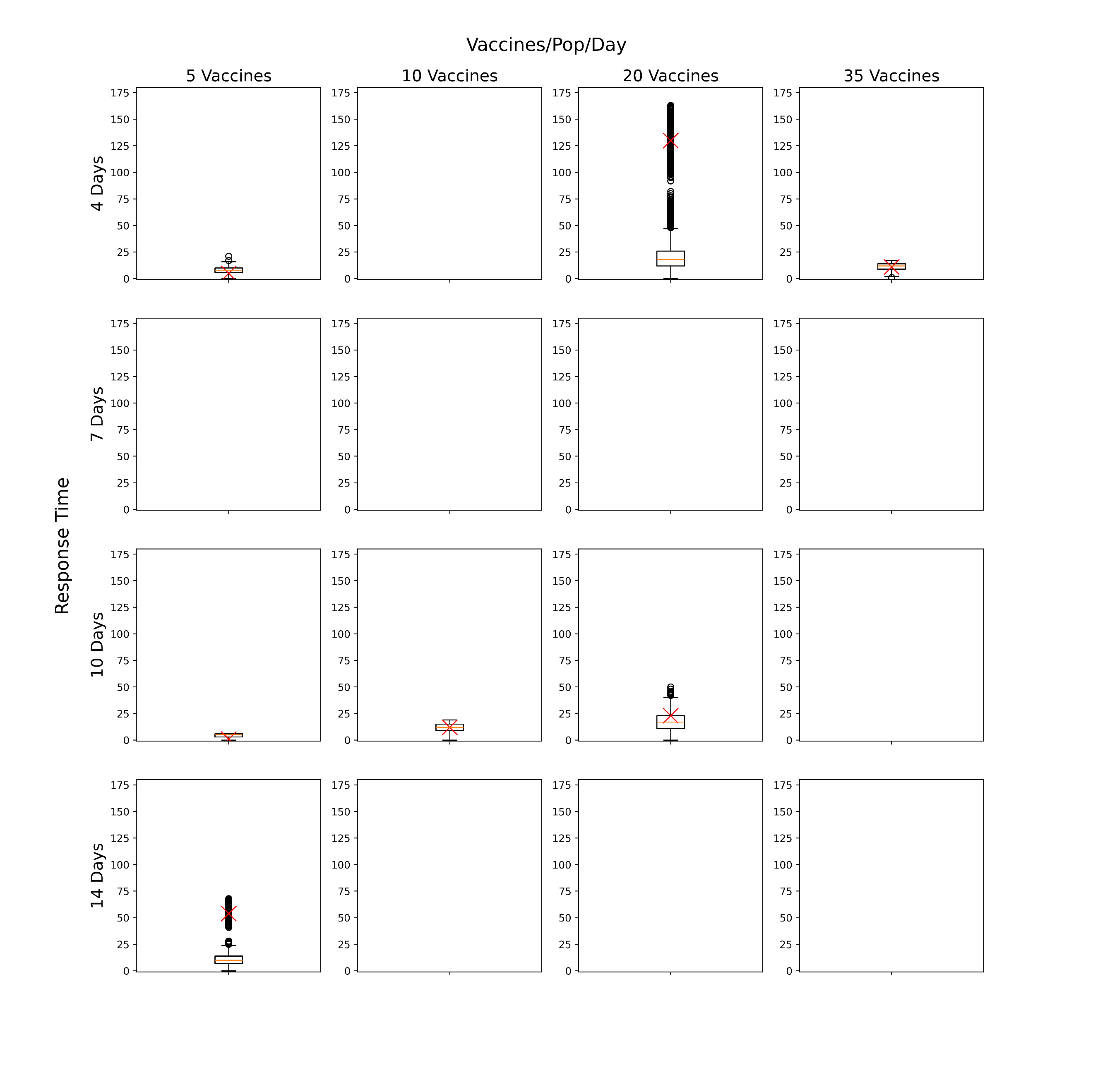

**Figure S28: Lattice of Ebola simulations before outbreak filtering was applied.** Each boxplot represents the distribution of total cases from simulations run with response parameters defined by the row/column lattice point. The red crosses represent the cumulative suspected cases from observed outbreaks which have been assigned to the lattice point. Lattice points are left blank if no outbreak was assigned.

#### **Scenarios**

For the purpose of running counterfactuals specific to the historical outbreaks, for each outbreak in the data, the set of simulations associated with the lattice point that the outbreak was assigned to were filtered to retain only those simulations where the cumulative cases were within +/- 25% of the reported cases (see Trajectory selection). Two scenarios were then examined:

- Baseline: ORI as occurred, using the filtered simulations from the lattice
- No ORI counterfactual: Simulations were run without any ORI intervention using the same filtered simulation seeds from the baseline scenario, such that everything was identical up until the date the ORI would have started.

#### **Outcomes**

For each outbreak and scenario, the distribution of cumulative cases and deaths across selected model simulations were recorded. The mean difference in outbreak size between scenarios and associated uncertainty in the mean were estimated from these collections of simulations using bootstrap resampling (i.e., for each outbreak to produce estimates of cases averted by ORI).

Total cases averted by ORI across all historic outbreaks was then estimated by aggregating the cases averted for each individual outbreak. As the outbreaks are independent, this was obtained by summing the cases averted per outbreak, with the variance estimated by summing the variances from each individual outbreak.

DALYs averted by ORI were estimated by multiplying cases averted by the disability weight per case and average duration of symptoms, plus years of life lost from deaths (for each outbreak, the average life expectancy in that country and year compared to the age of deaths in the model). Socio-economic costs averted by ORI were calculated by estimating productive years of life lost or lived with disability, and multiplying this by GDP per capita. Productive years of life lost were estimated as the difference between the average age of death in the model and a retirement age of 64, or the life expectancy in that country and year, whichever was lower. Costs were inflated to 2023 USD, with future costs discounted at 3% per annum as a standard method.

The impact of ORI on reducing the risk of large outbreaks was also estimated, by comparing the distribution of cumulative cases across outbreaks in the data to the distribution of cumulative cases across outbreaks in the no ORI scenario. For the purposes of this sub-analysis, cumulative cases in the no ORI scenario for each outbreak were approximated as the median from the no ORI counterfactual simulations.

#### **Results**

##### *Baseline*

The modelled cumulative cases and deaths for the baseline scenario is shown for each outbreak considered in the analysis in Figure S29. Outbreaks are coloured by the categorised response time or the ORI, and data points are scaled to the vaccine-eligible population in the model. The data points are scaled to the vaccine-eligible population in the model. The fit of the simulations to the case data presented in Figure S29 differs from what is presented in Figure S28 (i.e., the median of the model simulations sits close to the data) as we are considering only the filtered simulations which align well with the observed cases.

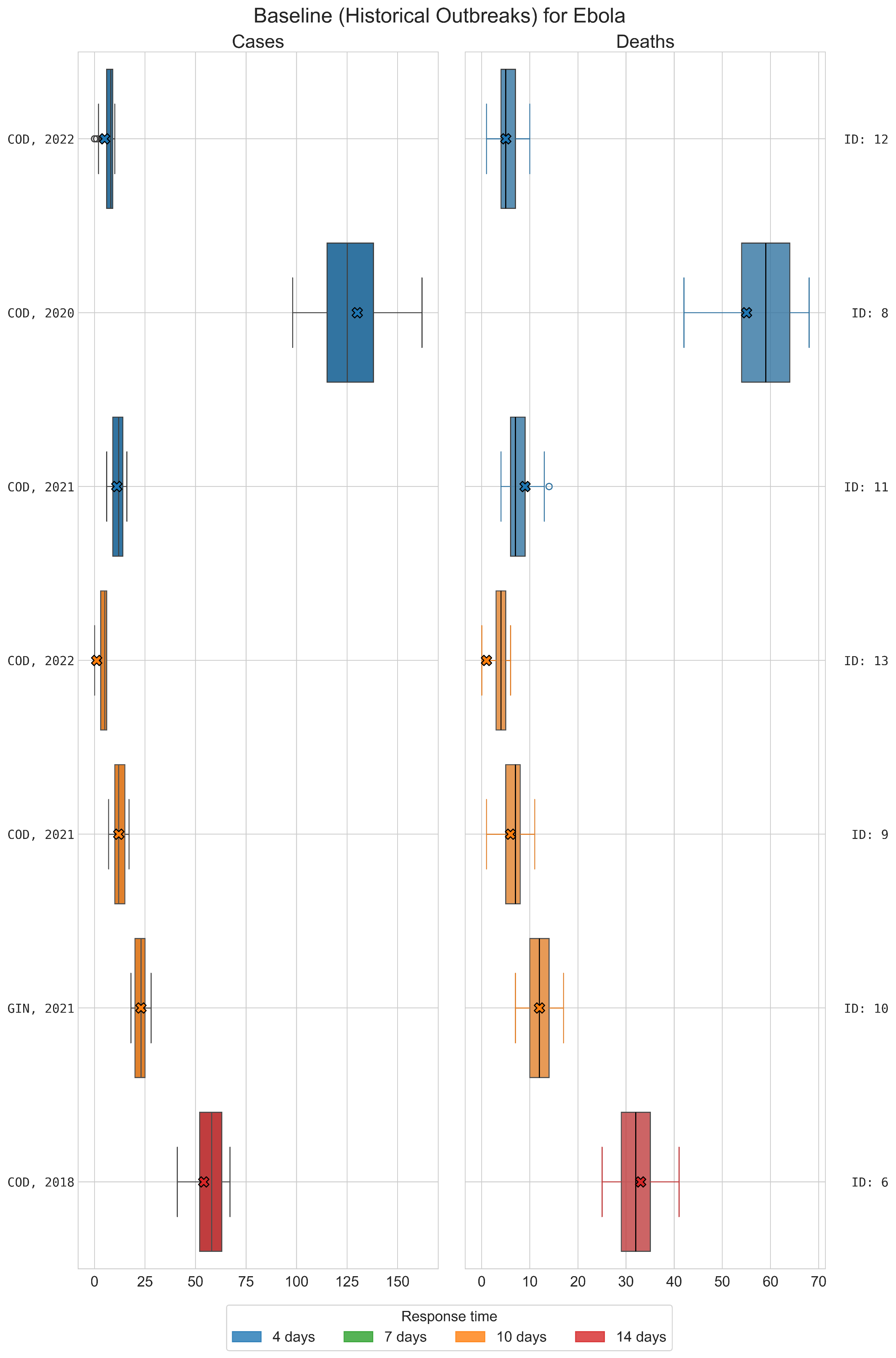

**Figure S29: Distribution of cases and deaths for each Ebola outbreak for model simulations retained after filtering.** Simulations which did not match within 25% of cumulative cases of historical outbreak were rejected. Crosses represent the data for each outbreak.

##### *Baseline vs. No ORI*

The distribution of simulated outcomes under the No ORI scenario are much wider than the equivalent Baseline scenario, as they are not constrained by filtering against observed cases, and always larger (Figure S30).

From Table S15 we can see that most of these impacts have been accrued across the five outbreaks since 2021, but that the impact across the two large outbreaks in 2018 and 2020 is almost equivalent.

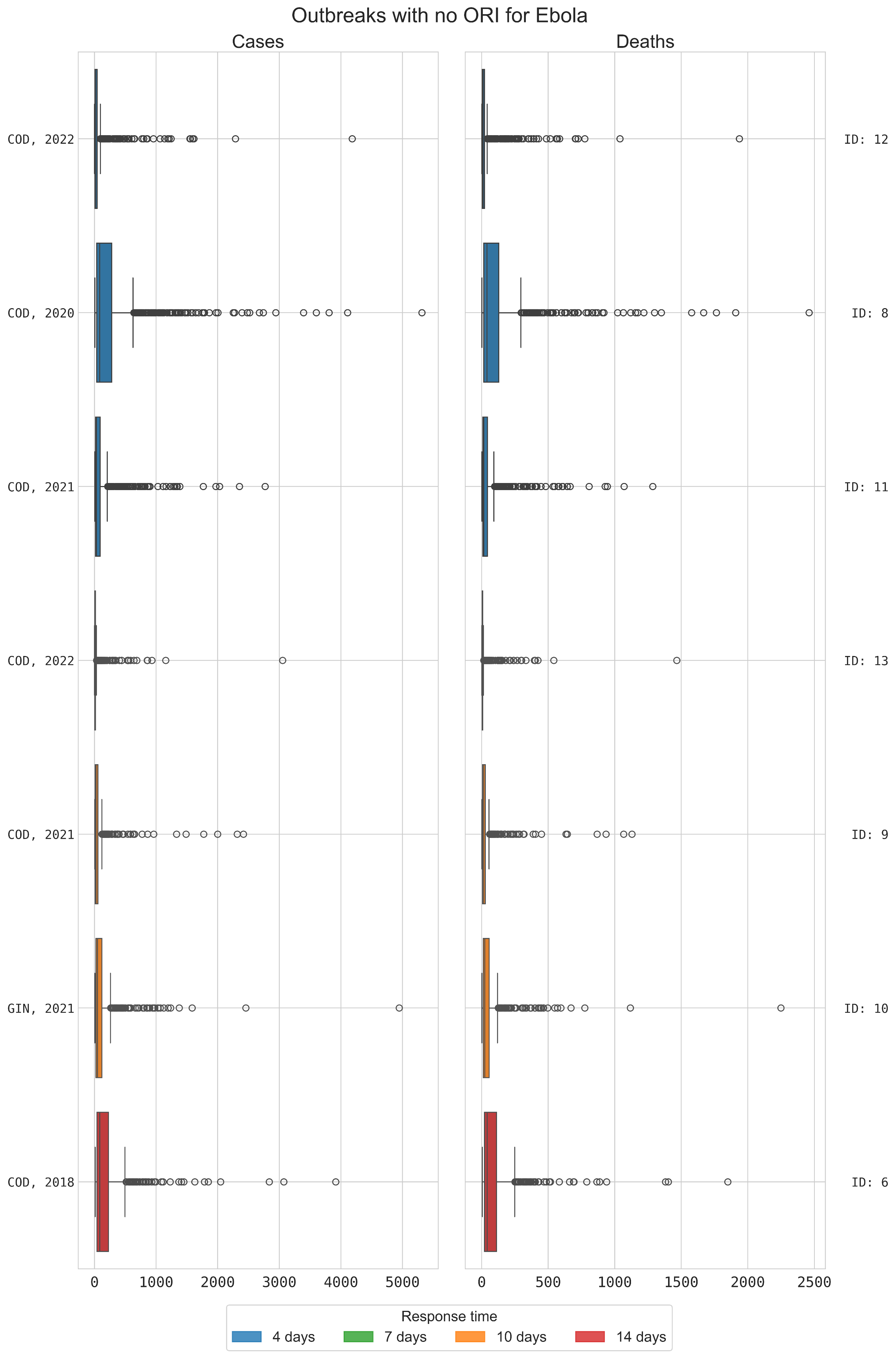

**Figure S30:** **Distribution of cases and deaths for each Ebola outbreak for counterfactual simulations with no ORI implemented.** Counterfactual simulations use equivalent transmission parameters and infection seeding to the matched baseline simulations.

**Table S15: Summary of estimated ORI impacts across Ebola outbreaks in five-year periods.** Estimated Cases/Deaths/DALYs/societal costs averted are summarized as the mean and 95% uncertainty intervals.

| Years | # outbreaks | Observed cases | Observed deaths | Cases averted | Deaths averted | DALYs averted (undiscounted) | Costs averted (discounted; 2023 USD) |
| --- | --- | --- | --- | --- | --- | --- | --- |
| 2016-2020 | 2 | 184 | 88 | 378 (305 – 452) | 175 (139– 212) | 7759 (6248– 9270) | 2·54M (2·04M – 3·04M) |
| 2021-2023 | 5 | 52 | 33 | 441 (328 – 553) | 204 (152 – 256) | 8856 (6575 – 11,137) | 4·17M (3·18M – 5·17M) |
| Total | 7 | 236 | 121 | 820 (633 – 1007) | 381 (292 – 469) | 16, 616 (12,824 – 20,409) | $6·72M ($5·23M – 8·21M) |

It is difficult to draw meaningful conclusions from the distribution of outbreak size thresholds for only seven outbreaks, but each of the outbreaks shifted to a higher threshold category without ORI, unless they already exceeded a 50-case threshold with ORI. With no ORI the percentage of outbreaks with more than 20 cumulative cases increased from 43% to 71%.

#### **Key messages**

The vaccine responses delivered to seven Ebola outbreaks from 2018-2022 averted significant burdens of disease, with more than half of the estimated impact being accrued since 2021.

We estimate that the ORI averted a cumulative:

- 633 – 1007 cases, compared to 236 observed
- 292 – 469 deaths compared to 121 observed
- 12,824 – 20,409 DALYs; and
- $5·23M – $8·21M societal economic costs

The presence of ORI was also estimated to have:

- Reduced the percentage of outbreaks with more than 20 cumulative cases from 71% to 43% (i.e. from five to three).

#### **Limitations**

- Actual transmission might be heterogeneous across age groups: there are likely age-based transmission effects beyond the household contact networks which the model implements which could have impacts on how transmission occurs, and age-based effects which impact how susceptible agents are to infection.
- There are limitations with estimating the at-risk population for the set of Ebola outbreaks considered: data were only available for three of the seven outbreaks, so we assumed that the model population represents a subset of the districts(s) where the outbreaks occurred and receives a proportion of the vaccines disbursed. This may produce an underestimate of the ORI impact, but we expect that the targeted ring-based response and the fact that most outbreaks are relatively small should minimise this effect.
- Contact tracing assumptions are uniform when they are likely variable across outbreaks: the effectiveness of the contact tracing performed during outbreaks likely varies based on the context, however we used a uniform set of assumptions for how effectively and quickly contacts are identified, followed-up, and quarantined. There is limited data to inform how this might vary across outbreak contexts, but it is expected to be reasonably accurate for smaller outbreaks. It may produce underestimates in the larger outbreaks in the dataset and in counterfactual simulations with no ORI as contact tracers would likely not be able to keep up as outbreaks grew too large, however the implementation in the model attempts to account for this by setting capacity limits on how many people can be traced each day.
- Uncertainty around vaccine efficacy: the efficacy value used in the model comes from a WHO report on the efficacy of the Ervebo vaccine. It has not been peer reviewed and the sample size used is relatively small. The efficacy reported is very high, and if the true efficacy is lower than the results presented here are likely to overestimate ORI impact.

**Supplement 7: Additional analyses**

Additionally, we have disaggregated results here to present only the estimated impacts accrued from outbreaks in settings which were eligible to access vaccines from a Gavi supported stockpile (199/210 total outbreaks). Table S16 presents the results disaggregated by disease and Table S17 presents the results disaggregated into five-year ranges which align with Gavi’s strategic funding periods.

**Table S16: Summary impacts of ORI (Gavi supported only) for the five diseases considered.** Estimated Cases/Deaths/DALYs/societal costs averted are summarized as mean (95% uncertainty interval).

| **Disease** | **Years** | **# outbreaks with ORI and sufficient data** | **Observed/  estimated cases** | **Observed / estimated deaths** | **Estimated cases averted** | **Estimated deaths averted** | **Estimated DALYs averted (undiscounted)** | **Estimated societal costs averted (discounted; 2023 US$)** |
| --- | --- | --- | --- | --- | --- | --- | --- | --- |
| Measles | 2003-2023 | 48 | 2·17M | 10,247 | 3·75M (3·70M – 3·81M) | 18,814 (18,451 – 19,176) | 1·18M (1·16M – 1·21M) | $622M ($609M – $635M) |
| Cholera | 2011-2023 | 38 | 806,592 | 9235 | 271K (263K – 280K) | 5017 (4682 – 5353) | 210K (195K – 225K) | $129M ($120M – $138M) |
| Yellow Fever | 2000-2023 | 83 | 19,455 | 2213 | 1·20M (1·16M – 1·25M) | 241K (232K – 250K) | 9·96M (9·58M – 10·34M) | $13·0B ($12·3B – $13·6B) |
| Meningitis | 2012-2023 | 23 | 57,488 | 4075 | 21,266 (20,676 – 21,856) | 1601 (1500 – 1702) | 113K (108K – 118K) | $110M ($105M – $114M) |
| Ebola | 2018-2022 | 7 | 246 | 121 | 820 (633 – 1007) | 381 (292 – 469) | 16, 616 (12,824 – 20,409) | $6·72M ($5·23M – $8·21M) |
| **Total** | | 199 | 3·05M | 25,891 | 5·25M (5·21M – 5·29M) | 267K (261K – 273K) | 11·5M (11·2M – 11·7M) | $13·8B ($13·4B – $14·2B) |

**Table S17: Summary impacts of ORI (Gavi supported only) across all five diseases in 5-year increments.** Estimated Cases/Deaths/DALYs/societal costs averted are summarized as mean (95% uncertainty interval).

| **Years** | **# outbreaks with ORI and sufficient data** | **Observed/  estimated cases** | **Observed / estimated deaths** | **Estimated cases averted** | **Estimated deaths averted** | **Estimated DALYs averted (undiscounted)** | **Estimated societal costs averted (discounted; 2023 US$)** |
| --- | --- | --- | --- | --- | --- | --- | --- |
| 2000 | 1 | 104 | 21 | 6428 (3871 - 8986) | 1296 (774 - 1818) | 54,483 (32,551 - 76,416) | $15·7M ($9·3M – $21·9M) |
| 2001-2005 | 16 | 4408 | 518 | 190K (163K – 218K) | 34,159 (30,421 – 37,898) | 1·44M (1·29M – 1·60M) | $903M ($979M – $1010M) |
| 2006-2010 | 22 | 424 | 105 | 7262 (2900 - 11,623) | 1447 (577 – 2318) | 56,563 (22,767 – 90,359) | $38·0M ($12·9M – $63·0M) |
| 2011-2015 | 48 | 1·16M | 2255 | 1·59M (1·53M – 1·65M) | 83,001 (73,904 – 92,098) | 3·78M (3·38M – 4·19M) | $6·16B ($5·44B – $6·87B) |
| 2016-2020 | 69 | 781,332 | 9945 | 2·79M (2·75M – 2·83M) | 121K (115K – 127K) | 5·07M (4·84M – 5·30M) | $6·17B ($5·87B – $6·48B) |
| 2021+ | 43 | 1·01M | 13,068 | 669K (653K – 685K) | 25,771 (24,486 – 27,055) | 1·07M (1·02M – 1·13M) | $540M ($495M – $585M) |
| **Total** | 199 | 3·05M | 25,891 | 5·25M (5·21M – 5·29M) | 267K (261K – 273K) | 11·5M (11·2M – 11·7M) | $13·8B ($13·4B – $14·2B) |

12.	Weekly Epidemiological Record (WER): World Health Organization; [Available from: <https://www.who.int/publications/journals/weekly-epidemiological-record>.

32.	Disease Outbreak News (DONs): World Health Organization; [Available from: <https://www.who.int/emergencies/disease-outbreak-news>.

33.	Cholera: International Coordinating Group (ICG) on Vaccine Provision; [Available from: <https://app.powerbi.com/view?r=eyJrIjoiYmFmZTBmM2EtYWM3Mi00NWYwLTg3YjgtN2Q0MjM5ZmE1ZjFkIiwidCI6ImY2MTBjMGI3LWJkMjQtNGIzOS04MTBiLTNkYzI4MGFmYjU5MCIsImMiOjh9>.

34.	Richterman A, Sainvilien DR, Eberly L, Ivers LC. Individual and Household Risk Factors for Symptomatic Cholera Infection: A Systematic Review and Meta-analysis. J Infect Dis. 2018;218(suppl_3):S154-S64.

42.	Assessing the impact of preventive mass vaccination campaigns on yellow fever outbreaks in Africa: A population-level self-controlled case series study [Internet]. PLOS Medicine. Available from: <https://doi.org/10.1371/journal.pmed.1003523>.

43.	WHO/UNICEF estimates of national immunization coverage [Available from: <https://immunizationdata.who.int/global/wiise-detail-page/yellow-fever-(yf)-reported-cases-and-incidence?CODE=Global&DISEASE=YFEVER>.

44.	Yellow Fever Outbreak Toolbox: World Health Organization; 2024 [Available from: <https://www.who.int/emergencies/outbreak-toolkit/disease-outbreak-toolboxes/yellow-fever-outbreak-toolbox>.

45.	Fraser K, Hamlet A, Jean K, Ramos DG, Romano A, Horton J, et al. Assessing yellow fever outbreak potential and implications for vaccine strategy. Research Square. 2024.

46.	Servadio JL, Muñoz-Zanzi C, Convertino M. Estimating case fatality risk of severe Yellow Fever cases: systematic literature review and meta-analysis. BMC Infectious Diseases. 2016;21(819).

47.	Hersi K, Gonzalez FJ, Kondamudi NP. Meningitis. StatPearls. Treasure Island (FL) ineligible companies. Disclosure: Francisco Gonzalez declares no relevant financial relationships with ineligible companies. Disclosure: Noah Kondamudi declares no relevant financial relationships with ineligible companies.2024.

70.	WHO Meningitis Bulletins: MenAfriNet; [Available from: <https://www.menafrinet.org/who-meningitis-bulletins>.

71.	Fernandez K, Lingani C, Aderinola OM, Goumbi K, Bicaba B, Edea ZA, et al. Meningococcal Meningitis Outbreaks in the African Meningitis Belt After Meningococcal Serogroup A Conjugate Vaccine Introduction, 2011-2017. J Infect Dis. 2019;220(220 Suppl 4):S225-S32.

72.	Kadanali A, Karagoz G. An overview of Ebola virus disease. North Clin Istanb. 2015;2(1):81-6.

73.	Patel PR, Shah SU. Ebola Virus. StatPearls. Treasure Island (FL) ineligible companies. Disclosure: Sumir Shah declares no relevant financial relationships with ineligible companies.2024.

74.	Wolf J, Jannat R, Dubey S, Troth S, Onorato MT, Coller BA, et al. Development of Pandemic Vaccines: ERVEBO Case Study. Vaccines (Basel). 2021;9(3).

75.	Prescott J, Bushmaker T, Fischer R, Miazgowicz K, Judson S, Munster VJ. Postmortem stability of Ebola virus. Emerg Infect Dis. 2015;21(5):856-9.

76.	Henao-Restrepo AM, Camacho A, Longini IM, Watson CH, Edmunds WJ, Egger M, et al. Efficacy and effectiveness of an rVSV-vectored vaccine in preventing Ebola virus disease: final results from the Guinea ring vaccination, open-label, cluster-randomised trial (Ebola Ca Suffit!). Lancet. 2017;389(10068):505-18.

77.	Preliminary results on the efficacy of rVSV-ZEBOV-GP Ebola vaccine using the ring vaccination strategy in the control of an Ebola outbreak in the Democratic Republic of the Congo: an example of integration of research into epidemic response.; 2019.

88.	Haas CN. On the quarantine period for ebola virus. PLoS Curr. 2014;6.

89.	Viral Hemorrhagic Fevers. Boca Raton, USA: CRC Press; 2013.

90.	Legrand J, Grais RF, Boelle PY, Valleron AJ, Flahault A. Understanding the dynamics of Ebola epidemics. Epidemiol Infect. 2007;135(4):610-21.

91.	Simpson DI. Marburg and Ebola virus infections: a guide for their diagnosis, management and control. Geneva, Switzerland: World Health Organization; 1977.

92.	Goeijenbier M, van Kampen JJ, Reusken CB, Koopmans MP, van Gorp EC. Ebola virus disease: a review on epidemiology, symptoms, treatment and pathogenesis. Neth J Med. 2014;72(9):442-8.

93.	Hartley MA, Young A, Tran AM, Okoni-Williams HH, Suma M, Mancuso B, et al. Predicting Ebola infection: A malaria-sensitive triage score for Ebola virus disease. PLoS Negl Trop Dis. 2017;11(2):e0005356.

94.	Ebola Disease Information for Clinicians in U.S. Healthcare Settings, U.S: Centers for Disease Control and Prevention; [Available from: <https://www.cdc.gov/vhf/ebola/clinicians/evd/clinicians.html>.

95.	McPake B, Witter S, Ssali S, Wurie H, Namakula J, Ssengooba F. Ebola in the context of conflict affected states and health systems: case studies of Northern Uganda and Sierra Leone. Confl Health. 2015;9:23.

96.	Kelly JD, Wannier SR, Sinai C, Moe CA, Hoff NA, Blumberg S, et al. The Impact of Different Types of Violence on Ebola Virus Transmission During the 2018-2020 Outbreak in the Democratic Republic of the Congo. J Infect Dis. 2020;222(12):2021-9.

97.	Dowell SF, Mukunu R, Ksiazek TG, Khan AS, Rollin PE, Peters CJ. Transmission of Ebola hemorrhagic fever: a study of risk factors in family members, Kikwit, Democratic Republic of the Congo, 1995. Commission de Lutte contre les Epidemies a Kikwit. J Infect Dis. 1999;179 Suppl 1:S87-91.

1. Including estimated values using the median model outcome and calibrated mortality rate. Please note 27% and 47% of outbreaks are missing the number of cumulative cases and deaths, respectively.
2. Please note 35% of outbreaks are missing the number of observed cumulative deaths, which is therefore estimated using the calibrated mortality rate.
